# Multiple Sclerosis: Exploring the Limits of Genetic and Environmental Susceptibility

**DOI:** 10.1101/2022.03.09.22272129

**Authors:** DS Goodin, P Khankhanian, PA Gourraud, N Vince

**Author notes:** Address for Correspondence: Douglas S. Goodin, MD, Department of Neurology, University of California, San Francisco, UCSF MS Center, 675 Nelson Rising Lane, Suite #221D, San Francisco, CA 94158, E mail. **Author Contributions** DSG: Conceptualized and led the project, wrote the analytic software, analyzed and interpreted the data, and wrote the original draft of the manuscript. PK: Assisted in data interpretation and review of the manuscript. PA: Assisted in data interpretation and review of the manuscript. NV: Assisted in data interpretation, and review of the manuscript.

## Abstract

**OBJECTIVE:** To explore the nature of genetic and environmental susceptibility to multiple sclerosis (MS) and to define the limits of this nature based on the statistical uncertainties regarding the various epidemiological observations that have been made.

**BACKGROUND:** Certain parameters of MS-epidemiology are directly observable (e.g., the risk of MS-recurrence in siblings and twins of an MS proband, the proportion of *women* among MS patients, the population-prevalence of MS, and the time-dependent changes in the female-to-male (*F:M*) *sex-ratio*. By contrast, other parameters can only be inferred from observed parameters (e.g., the proportion of the population that is genetically susceptible, the proportion of *women* among susceptible individuals, the probability that a susceptible individual will experience an environment sufficient to cause MS given their genotype, and if they do, the probability that they will develop the disease).

**DESIGN/METHODS:** The “genetically-susceptible” subset (*G*) of the population (*Z*) is defined to include everyone with <u>any</u> non-zero life-time chance of developing MS under *some* environmental conditions. For the observed parameters, acceptable ranges are assigned values such that they always include their 95% confidence intervals. By contrast, for the non-observed parameters, the acceptable ranges are assigned such that they cover the entire “plausible” range for each parameter. Using both a *Cross-sectional Model* and a *Longitudinal Model*, together with established parameter relationships, we explore, iteratively, trillions of potential parameter combinations and determine those combinations (i.e., solutions) that fall within the acceptable range for the observed and non-observed parameters.

**RESULTS:** Both *Models* and all analyses are consistent and converge to demonstrate that genetic-susceptibitly is limited to 52% or less of the population and to 30% or less of *women*. Consequently, most individuals (particularly *women*) have no chance whatsoever of developing MS, regardless of their environmental exposure. Also, currently, the penetrance of MS in susceptible *women* is greater than it is in *men*. Moreover, as expected, the probability that susceptible individuals will develop MS increases with an increased likelihood of these individuals experiencing an environment sufficient to cause MS, given their genotype. Nevertheless, although it is conceivable that these response-curves plateau at 100% for both *women* and *men*, this possibility requires extreme conditions and seems remote. Rather, at least *men*, seem to plateau well below this level and, if so, it is this difference, rather than any differences in the genetic and environmental determinants of disease, that primarily accounts both for the difference in penetrance between *women* and *men* and for the increasing proportion of *women* among of MS patients worldwide.

**CONCLUSIONS:** The development of MS (in an individual) requires both that they have an appropriate genotype (which is uncommon in the population) and that they have an environmental exposure sufficient to cause MS given their individual genotype. Nevertheless, even when the necessary genetic and environmental factors, sufficient for MS pathogenesis, co-occur for an individual, this still insufficient for that person to develop MS. Thus, disease pathogenesis, even in this circumstance, seems not to be deterministic but, rather, to involve an important element of chance.

**Author Summary:** Certain parameters of MS-epidemiology can be directly observed. These parameters include the risk of MS recurrence in siblings and twins of an MS proband, the proportion of *women* among MS patients, the population-prevalence of MS, and the time-dependent changes in the female-to-male (*F:M*) *sex-ratio*. By contrast, there are other parameters of MS-epidemiology, which can’t be observed, but which must be inferred based on the values of the observable parameters. These parameters include the proportion of the general population (*Z*) that is genetically susceptible to MS, the proportion of *women* among susceptible individuals, the probability that a susceptible individual will experience an environment sufficient to cause MS, and if they do, the likelihood that they will, in fact, develop the MS. We define the subset (*G*) – i.e., the genetically-susceptible subset – to include everyone in (*Z*) who has <u>any</u> non-zero chance of developing MS over their life-time, under some environmental circumstances. For the observed parameters, plausible ranges are assigned acceptable values such that they always include their 95% confidence interval. By contrast, for the non-observed parameters, the acceptable ranges are assigned such that they cover the entire “plausible” range for each parameter. Then, using both a *Cross-sectional Model* and a *Longitudinal Model*, together with established parameter relationships, we explore iteratively trillions of potential parameter combinations and determine those combinations (i.e., solutions) that are allowed by the observed and non-observed parameter ranges. The *Cross-sectional Model* makes two assumptions, commonly made in studies of monozygotic twins, to establish certain relationships between the observed and non-observed parameters. By contrast, the *Longitudinal Model* makes neither of these assumptions but, rather, this *Model* utilizes the observed changes in the female-to-male (*F:M*) *sex-ratio* and the disease prevalence, which have taken place over the past 4–5 decades, to determine the response curves for susceptible individuals, relating their probability of developing MS to their probability of experiencing an environment sufficient to cause MS. Both *Models* and all analyses are consistent with each other and converge to demonstrate that genetic-susceptibitly is limited to 52% or less of the population and 30% or less of women. Consequently, most individuals have no chance whatsoever of developing MS, regardless of their environmental experiences. Thus, MS is a genetic disease in the sense that, if an individual does not have the correct genetic makeup, they can’t develop the disease. However, the probability that susceptible individuals will develop MS increases with an increased likelihood of these individuals experiencing an environment sufficient to cause MS, given their genotype. Thus, MS is also and environmental disease in the sense that the development of MS (in an individual), in addition to their having an appropriate genotype, requires that they experience an environmental exposure sufficient to cause MS given their individual genotype. Nevertheless, there must be another factor involved in disease pathogenesis because, although it is conceivable that these response-curves plateau at 100% for both *women* and *men*, this possibility requires extreme conditions and seems remote. Rather, at least *men*, seem to plateau well below this and, if so, it is this difference, rather than differences in the genetic and environmental determinants of disease, that primarily accounts both for the difference in penetrance between *women* and *men* and for the increasing proportion of *women* among of MS patients worldwide. Consequently, even when the necessary genetic and environmental factors, sufficient for MS pathogenesis, co-occur for an individual, this still seems to be insufficient for that person to develop MS. Thus, disease pathogenesis, even in this circumstance, seems not to be deterministic but, rather, to involve an important element of chance.

## Introduction

Susceptibility to multiple sclerosis (MS) is known to be complex, involving the critical interplay between both environmental events and genetic factors [1-3]. Our previously published analysis regarding the nature of this susceptibility [3] was based on a few basic, well-established, epidemiological parameters of MS, which have been repeatedly observed in populations across Europe and North America. These parameters include the prevalence of MS in a population, the recurrence-risk for MS in siblings and twins of individuals with MS, the proportion of *women* among MS patients, and the time-dependent changes in both the female-to-male (*F:M*) *sex-ratio* and the disease prevalence, which have taken place over the last several decades [3]. For this analysis, we defined a “genetically susceptible” subset (*G*) of the general population (*Z*) to include everyone who has any non-zero chance of developing MS over the course their lifetime. We concluded that genetic susceptibility, so-defined, is limited to only a small proportion of these northern populations (<7.3%) and, thus, that most individuals in these populations have no chance whatsoever of developing MS, regardless of any environmental conditions that they may experience during their lifetimes [3]. Nevertheless, despite this critical dependence of susceptibility to MS upon the genotype of an individual, we also concluded that certain environmental events were also necessary for MS to develop and that, consequently, both essential genetic factors and essential environmental events are in the causal pathway leading to MS [3]. If either of these are missing, MS cannot develop. Finally, we concluded that, seemingly, even when the sufficient genetic and environmental determinants were present, the actual development of MS depended, in part, upon an element of chance [3].

What this analysis did not undertake, however, was to explicitly explore the limits of these conclusions based upon the statistical uncertainties, which surround each of the various epidemiological observations that have been made. It is the purpose of this study, therefore, to undertake such an exploration using both the confidence intervals (*CIs*) and “plausible” ranges for the different basic epidemiological parameters and by incorporating these uncertainties into the governing equations relating these parameters both to each other and to the underlying susceptibility to MS that exists within in a population.

For this analysis, we have used, primarily, the data reported from the Canadian Collaborative Project on Genetic Susceptibility to Multiple Sclerosis [4,5]. The reason for this choice is three-fold. First, this Canadian dataset is a *population-based* sample with an initial cohort of 29,478 MS patients who were born between the years 1891 and 1993 [4-7]. This cohort consists of all MS patients seen in 15 MS Centers scattered throughout the Canadian Provinces [5]. The cohort did not specifically include patients from the Northern Territories [5] although, likely, many of these patients were referred for 2^nd^ opinions to the provincial centers. This study endeavored to include most (or all) of the MS patients in Canada at the time and, indeed, the authors estimate (from their twin studies) that their ascertainment scheme captured 65-83% of all Canadian MS patients [7]. Importantly, also, this cohort ascertained cases from throughout Canada. Therefore, for the purposes of our analysis, this cohort is assumed to represent a large random sample of the symptomatic Canadian MS population at the time. Second, this dataset provides, from the same population, estimates for the recurrence-risk in monozygotic (*MZ*) twins, in dizygotic (*DZ*) twins, in non-twin siblings (*S*), and for changes in the (*F:M*) *sex-ratio* over time [4-7]. Consequently, this Canadian dataset is likely among the most complete and the most reliable in the world. And third, these data come from a geographic region of similar latitude, which is critical when considering a disease, for which disease prevalence has a marked latitudinal gradient in different parts of the world [8].

## Methods

### 1. General Methods

#### A. General Model Specifications and Definitions for Genetic Susceptibility to MS

We consider a general population (*Z*), which is composed of (*N*) individuals (*k* = 1,2, …, *N*) who are living under the prevailing environmental conditions during some specific *Time Period* (*T*) – conditions that are designated, generically, as (*E*_*T*_). In *Table 1*, we define the different parameters used in our analysis and, in *Table 2*, we provide a set of parameter abbreviations, which are used for the purposes of notational simplicity. The subset (*MS*) is defined to include *<u>all</u>* individuals within (*Z*) who either have or will develop MS over the course of their lifetime. The occurrence of (*MS*) represents the event that an individual, randomly selected from (*Z*), belongs to this (*MS*) subset and the term *P*(*MS*|*E*_*T*_) represents the probability of this event, given the prevailing environmental conditions of (*E*_*T*_) – i.e., *P*(*MS*|*E*_*T*_) = *P*(*MS*|*Z, E*_*T*_). This probability is referred to as the “*penetrance*” of MS for the population (*Z*) during the *Time Period* (*E*_*T*_). The occurrence of (*G*) is defined as the event that an individual, randomly selected from (*Z*), is a member of the (*G*) subset. The term *P*(*G*|*E*_*T*_) represents the probability of this event given the prevailing environmental conditions of (*E*_*T*_). In turn, the (*G*) subset is defined to include *<u>all</u>* individuals (genotypes) within (*Z*) who have *<u>any</u>* non-zero chance of developing MS under *<u>some</u>* (unspecified and not necessarily realized) environmental conditions, regardless of how small that chance might be. We assume that a person’s genotype is independent of the environmental conditions that prevail during (*E*_*T*_). Therefore:

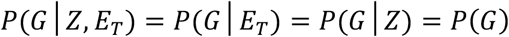

**Table 1.** Definitions for the Groups and Epidemiological Parameters used in the Analysis

| Parameter | Definition |
| --- | --- |
| $(Z)$ | Set of all $(N)$ individuals (i.e., unique genotypes) in the population |
| $(G), (G^c)$ | Subsets of individuals (genotypes) in $(Z)$ who have <u>any</u> non-zero chance $(G)$ or no chance $(G^c)$ of developing MS |
| $(G_{ws}), (G_{dws})$ | Subset of susceptible *women*: $(G_{ws}) = (F, G)$ ;<br>$(G_{dws}) =$ the $d^{th}$ susceptible *woman* in $(G)$ |
| $(G_i), (G_{is}), (G_{ia})$ | Genotypes or subsets either of the $i^{th}$, or an *i-type*, individual in $(G)$:<br>$(G_i)$ = full unique genotype; $(G_{is})$ = genotype of all *MS*-related factors<br>$(G_{ia})$ = genotype of only autosomal *MS*-related factors |
| $\{G_s\}, \{G_a\}$ | Families of sets: – $\{G_s\}$ includes all subsets $(G_{is})$ within $(G)$; and<br>$\{G_a\}$ includes all subsets $(G_{ia})$ within $(G)$ |
| $(M), (F)$ | Subsets of *men* $(M)$ and *women* $(F)$ in $(Z)$ -- $P(M) + P(F) = 1$ |
| $(G1), (G2)^\dagger$ | “*High*” $(G1)$ and “*low*” $(G2)$ penetrance” subsets in $(G)$ – *see Text* |
| $(G1'), (G2')^\dagger$ | “*High*” $(G1')$ and “*low*” $(G2')$ penetrance” subsets in $(F, G)$ or $(M, G)$ |
| $(MZ), (DZ), (S)$ | Subsets *MZ*-twins, *DZ*-twins, or non-twin siblings $(S)$ in $(Z)$ |
| $(E_T)^\dagger$ | The prevailing environmental conditions during the *Time Period* $(T)$ |
| $\{E_i\}^\dagger$ | Family of sets of environmental exposures, each of which is sufficient, by itself, to cause MS in the $i^{th}$ individual in $(G)$ – *see Text* |
| $\{E_{iw}\}^\dagger$ | Family of sets of more “*intense*” environmental exposures, within $\{E_i\}$, required by an *i-type woman* – $(E_{iw}) \subset (E_i)$ – *see Text* |
| $(E)^\dagger$ | Union of all disjoint events $(\{E_i\}, G_i)$ – *see Text* |
| $(E_w)^\dagger$ | Union of all disjoint events $(\{E_{iw}\}, G_i) - (E_w) \subset (E)$ – *see Text* |
| $(E_{pop}), (E_{sib}), (E_{twn})$ | Distinct parts of a sufficient environmental exposure, equally likely to be shared by anyone $(E_{pop})$, more or less likely to be shared by siblings $(E_{sib})$ and more or less likely to be shared by twins $(E_{twn})$ – *see Text* |
| $(MS)^\dagger$ | Subset of individuals in $(Z)$ who either have, or will develop, *MS* |
| $(MZ_{MS}), (DZ_{MS}), (S_{MS})$ | Subsets of *MZ* co-twins $(MZ_{MS})$, *DZ* co-twins $(DZ_{MS})$, or non-twin co-siblings $(S_{MS})$ who either have, or will develop, *MS* – *see Text* |
| $P(MS)^\dagger$ | Life-time probability of developing *MS* for a member of $(Z)$: |
| $P(MS \vert MZ_{MS})^\dagger$<br>$P(MS \vert DZ_{MS})^\dagger$<br>$P(MS \vert S_{MS})^\dagger$ | $P(MS)$ for a proband, randomly selected from $(Z)$, who has a co-twin or co-sibling in the $(MZ_{MS})$, $(DZ_{MS})$, or $(S_{MS})$ subsets – *see Text* |
| $P(MS \vert IG_{MS})^\dagger$ | $P(MS \vert MZ_{MS})$ adjusted for the shared environment of *MZ*-twins |
| $P(MZ_{MS})^\dagger$<br>$P(IG_{MS})^\dagger$ | $P(MS)$ for the co-twin from an *MZ* twin-pair, without considering the proband’s circumstances. $P(MZ_{MS}) = P(IG_{MS}) = P(MS)$ – *See Text* |

† Parameters that vary with environmental conditions $(E_T)$ , which is indicated in different manners in the *Text*, depending upon context. For example, during: $(E_T) = \text{Time Period \#2}$ :
The subscript “*MS*” indicates that the non-proband relative preceding the subscript has or will develop *MS*

**Table 2.** Principal Parameter Abbreviations^†^

| Parameter | Definition |
| --- | --- |
| $(F, w) \text{ \& } (M, m)$ | Alternate indicators for: female/women ($F, w$) and: male/men ($M, m$) |
| *Subscripts* (1,2) | Indicators for either *Time Period #1* or the “current” *Time Period #2*<br>-- except for the parameters: $x_1, x_2, x'_1, x'_2, y'_1$, and $y'_2$ -- see below |
| $x$ | Penetrance of MS for the ($G$) subset -- $x = P(MS \vert G)$ |
| $x_i$ | Penetrance of the ($i^{th}$) genotype in ($G$) -- $\forall G_i \in G: P(MS \vert G_i) = x_i$ |
| $X$ | Set of all individual penetrance values in ($G$) -- $X = \{x_i\}$ |
| $x'$ | Adjusted MZ-twin concordance for MS -- $x' = P(MS \vert IG_{MS})$ |
| $\sigma_X^2$ | Variance of the set ($X$) -- $Var(X)$ |
| $x_1$ | Penetrance of MS for ($G1$) subset -- $x_1 = P(MS \vert G1)$ |
| $x_2$ | Penetrance of MS for ($G2$) subset -- $x_2 = P(MS \vert G2)$ |
| $Zw$ | Failure probability in *women*: $Zw = P(MS, E_w \vert F, G) = x_1$ |
| $Zm$ | Failure probability in *men*: $Zm = P(MS, E \vert M, G) = x_2$ |
| $x'_1$ | Adjusted MZ-twin concordance for ($G1$) subset -- $x'_1 = P(MS \vert G1, IG_{MS})$ |
| $x'_2$ | Adjusted MZ-twin concordance for ($G2$) subset -- $x'_2 = P(MS \vert G2, IG_{MS})$ |
| $y'_1$ | MZ-twin concordance for the ($G1$) subset -- $y'_1 = P(MS \vert G1, MZ_{MS})$ |
| $y'_2$ | MZ-twin concordance for the ($G2$) subset -- $y'_2 = P(MS \vert G2, MZ_{MS})$ |
| $u$ | Odds of a sufficient environmental exposure -- $u = P(E) / [1 - P(E)]$ |
| $q_m^{min}, q_w^{min}$ | “Minimum” exposure level change in *men* ($q_m^{min}$) and *women* ($q_w^{min}$) |
| $q_m, q_w$ | “Actual” exposure level change in *men* ($q_m$) and *women* ($q_w$) |
| $h(u), k(u)$ | Hazard functions for *men* – $h(u)$ ; and for *women* – $k(u)$ |
| $R$ | “Actual” proportionality factor (if proportional) -- $k(u) = R * h(u)$ |
| $R_i, R_{dw^s}$ | “Actual” value of $R$ in *i-type* ($R_i$) and individual ($R_{dw^s}$) *women* |
| $R^{app}$ | “Apparent” value of $R$ -- $R^{app} = q_w^{min} / q_m^{min}$ |
| $\lambda_w, \lambda_m$ | Environmental threshold to develop MS in *women* ($\lambda_w$) and *men* ($\lambda_m$) |
| $\lambda, \lambda_i$ | Threshold differences, generally: $\lambda = (\lambda_w - \lambda_m)$; and for *i-types* ($\lambda_i$) |
| $C$ | Ratio of $P(MS)$ at *Time #1* to that at *Time #2* -- $C = P(MS)_1 / P(MS)_2$ |
| $C_F, C_M$ | The ratio ($C$) for *women* ($C_F$) ; and for *men* ($C_M$) |
| $p, p'$ | Proportion of *women* in subset ($G$) – $p$ ; and in subset ($MS, G$) – $p'$ |
| $r, s$ | Enrichment of genotypes – *women*: $r = (x'_1 / x_1)$ ; *men*: $s = (x'_2 / x_2)$ |
| $c, d$ | Limits for the exponential response curves in *men* ($c$) and *women* ($d$) |
| $s_a$ | Factor to adjust $P(MS \vert MZ_{MS})$ because of the shared twin environment<br>$P(MS \vert IG_{MS}) = P(MS \vert MZ_{MS}) / s_a$ ; $s_a \geq 1$ – see Text |

<sup>†</sup> Each of these parameters (except $h(u), k(u), R, R_i, R_w, \lambda, \lambda_w, \lambda_m, p, c, \text{ \& } d$ ) vary with different environmental conditions ( $E_T$ ), which is indicated in various ways in the Text– see Table 1.

In this circumstance, each of the (*m* ≤ *N*) individuals in the (*G*) subset (*i* = 1,2, …, *m*) has a unique genotype (*G*_*i*_). The occurrence of (*G*_*i*_) represents the event that an individual, randomly selected from (*Z*), belongs to the (*G*_*i*_) subset – a subset consisting of only a single individual (i.e., the so-called “*i*^*th*^ susceptible individual” or “*i*^*th*^ individual”) – and the term {*P*(*G*_*i*_) = 1/*N*} represents the probability of this event. Therefore, it follows from the definition of the (*G*) subset that, if *<u>every</u>* relevant environmental condition – *see below* – is possible during some *Time Period* (*E*_*T*_), then, during this *Time Period*:

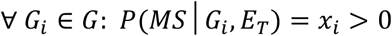

The conditional probabilities: {*x*_*i*_ = *P*(*MS*|*G*_*i*_, *E*_*T*_)} and: {*x* = *P*(*MS*|*G, E*_*T*_)}, are referred to as the “*penetrance*” of MS, during (*E*_*T*_), respectively, for *i*^*th*^ susceptible individual and for the (*G*) subset. Clearly, the *penetrance* of MS, both for the individual and for the group, will vary depending upon the likelihood of different environmental conditions during different *Time Periods*. If the environmental conditions during some *Time Periods* were such that certain members of the (*G*) subset have no possibility of ever developing MS then, for these individuals, during these *Time Periods*:

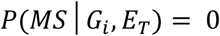

Because, currently, some individuals do develop MS, it must be that, during our “current” *Time Period*:

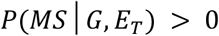

However, if, at some other time, the environmental conditions were such that no member of (*G*) could ever develop MS then, during these *Time Periods*:

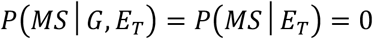

We also define a subset 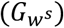, which consists of all *women* within the (*G*) subset {i.e., 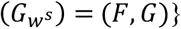}, and we define the proportion of *women* in (*G*) as: *p* = *P*(*F*|*G*). In this circumstance, each of the (*m* * *p*) *women* in the 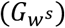 subset (*d* = 1,2, …, *mp*) has a unique genotype 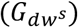. The occurrence of 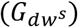 represents the event that an individual, randomly selected from (*Z*), belongs to the 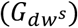 subset – a subset consisting of only the *d*^*th*^ susceptible *woman* – and the term 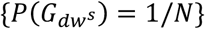 represents the probability of this event. Also, 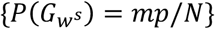 represents the probability of the event that an individual, randomly selected from (*Z*), belongs to the 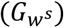 subset.

Individuals, who do not belong to the (*G*) subset, belong to the mutually exclusive (complimentary) subset (*G*^*c*^), which consists of *<u>all</u>* individuals who have *<u>no</u>* chance, whatsoever, of developing MS, regardless of any environmental experiences that they either have had or could have had. The occurrence of (*G*^*c*^) is defined as the event that an individual, randomly selected from the population (*Z*), is a member of the (*G*^*c*^) subset. The term *P*(*G*^*c*^|*E*_*T*_) represents the probability of this event, given the environmental conditions of (*E*_*T*_). Consequently, each of the (*m*^*c*^ = *N* − *m*) “non-susceptible” individuals in the (*G*^*c*^) subset (*j* = 1,2, …, *m*^*c*^) has a unique genotype (*G*_*j*_). As *above*, the occurrence of (*G*_*j*_) represents the event that an individual, randomly selected from (*Z*), belongs to the (*G*_*j*_) subset – a subset also consisting of a single individual. The term {*P*(*G*_*j*_) = 1/*N*} represents the probability of this event. Thus, under *<u>any</u>* environmental conditions, during *<u>any</u> Time Period*:

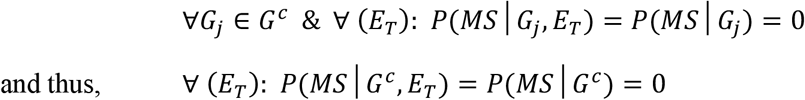

Notably, “identical” twins, despite having nearly “identical” genotypes (*IG*), nevertheless, still have subtle genetic differences from each other. Thus, even if these subtle differences are irrelevant to MS susceptibility (as seems likely, and which we assume to be true), these differences still exist. Consequently, every individual – i.e., each complete genotype (*G*_*i*_) and (*G*_*j*_) – in the population is unique. Despite this uniqueness, however, we can also define a so-called “susceptibility-genotype” for the *i*^*th*^ susceptible individual such that this genotype consists of all (and only) those genetic factors, which are related to MS susceptibility. Because the specification of such a susceptibility-genotype necessarily includes many fewer genetic factors than the *i*^*th*^ individual’s complete genotype, it is possible that one or more other individuals in the population share the same susceptibility-genotype with the *i*^*th*^ individual. For example, in this conceptualization, *MZ-*twins would necessarily belong to the same susceptibility-genotype. We refer to the group of individuals, who belong to the *i*^*th*^ susceptibility-genotype, either as the (*G*_*is*_) subset within (*Z*) or as the “*i-type*” group. Similarly, we refer to individuals who are members of this subset, as “*i-type*” individuals. In this case, the occurrence of (*G*_*is*_) represents the event that a person, randomly selected from (*Z*), belongs to the (*G*_*is*_) subset, which consists of only a single *i-type*. The term *P*(*G*_*is*_) represents probability of this event. Clearly, because some members of (*G*) are *MZ*-twins, the total number of such *i-type* groups in the population (*m*_*it*_) will be less than (*m*). In addition, we define the family {*G*_*s*_} to include all the (*G*_*is*_) subsets (or *i-type* groups) within (*Z*) and define the event {*G*_*s*_} as representing the union of the disjoint (*G*_*is*_) events such that:

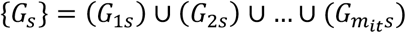

Because every susceptible person belongs to one and only one of these *i-type* groups, the probability of this event is expressed as:

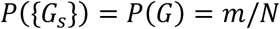

We also define the set (*X*) to be the set of penetrance values for members of the (*G*) subset during some *Time Period*. Provided that the variance of (*X*) is not equal to zero {i.e., 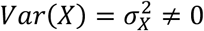}, the subset (*G*) can be further partitioned into two mutually-exclusive subsets, (*G*1) and (*G*2), suitably defined, such that the penetrance of MS for the subset (*G*1) during a certain *Time Period* is greater than that for (*G*2). The terms *P*(*G*1) and *P*(*G*2), represent the probabilities of the events that an individual, randomly selected from (*Z*), is a member of the subsets (*G*1) and (*G*2), respectively. Although many such partitions are possible, for the purposes of the present manuscript, (*G*1) is generally considered interchangeable with the subset of susceptible *women* – i.e., (*G*1) = (*F* ∩ *G*) = (*F, G*) – and (*G*2) is generally considered interchangeable with the subset of susceptible *men* – i.e., (*G*2) = (*M* ∩ *G*) = (*M, G*).

When considering the enrichment of more penetrant genotypes (*see Methods #2C*), the subsets (*F, G*) and (*M, G*) will each be further partitioned into high- and low-penetrance sub-subsets – i.e., (*G*1^′^) and (*G*2^′^), respectively – where the basis for this further partition into (*G*1^′^) and (*G*2^′^) sub-subsets is unspecified. The definitions of, and the probabilities for, these events mirrors that *above* for (*G*1) and (*G*2). Moreover, although the basis for this further partition must be something other than gender, it can be anything else that creates a partition, and it doesn’t need to be the same basis for both genders.

{*NB: A note on terminology. When a claim refers to any partition of the* (*G*) *subset, the probabilities of developing MS (i*.*e*., *the penetrance of MS) for members of the* (*G*1) *and* (*G*2) *subsets are designated, respectively, such that: x*_1_ = *P*(*MS*|*G*1) *and: x*_2_ = *P*(*MS*|*G*2). *When the partition is based specifically on gender, and to avoid any confusion with our Time Period designations, the group of females/women are indicated, alternatively, either by an upper-case (F) or by a lower-case (w). Similarly, in these circumstances, males/men are indicated, alternatively, either by an upper-case (M) or by a lower-case (m) – see Table 2. In some circumstances (when the meaning is clear), for purposes of notational simplicity, the designations of* (*x*_1_) *and* (*x*_2_) *continue to be used to designate the penetrance of MS for the subsets of susceptible women* (*x*_1_) *and men* (*x*_2_). *In other circumstances, however, greater clarity is provided by using the letter designations. For example, considering the partition of* (*G*) *into the subsets* (*F, G*) *and* (*M, G*), *the penetrance of MS for susceptible women and men are designated, respectively, as* (*Zw*) *and* (*Zm*) *such that: x*_1_ = *Zw* = *P*(*MS*|*F, G*) *and: x*_2_ = *Zm* = *P*(*MS*|*M, G*) – *see Table 2*.

*Moreover, although this manuscript focuses on the gender partition for the disease MS, the Models developed pertain to any partition for any disease, which has data analogous to that found in Canada for the gender partition of MS [6,7]*.}

#### B. General Model Specifications and Definitions for Environmental Susceptibility to MS

The term {*E*_*i*_} represents the family of specific sets of environmental exposures, each of which, by itself, is sufficient to cause MS to develop in the *i*^*th*^ susceptible individual. Each set within the {*E*_*i*_} family must be distinct (in some respect) from every other set within this family but, otherwise, there can be any degree of overlap between the factors or events that comprise these sets. Also, there can be any number of sets within the {*E*_*i*_} family although, because (∀ *G*_*i*_ ∈ *G*: *x*_*i*_ > 0) under *<u>some</u>* environmental conditions, the family cannot be empty. If we assign (*v*_*i*_) to the number of sets of sufficient exposures for the *i*^*th*^ susceptible individual, then {*E*_*i*_} represents the family of sets: 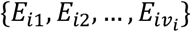; and *P*({*E*_*i*_}|*E*_*T*_) represents the probability of the event that, at least, one of these sets of sufficient exposures occurs, given the prevailing environmental conditions of the time (*E*_*T*_). Moreover, if more than one individual belongs to a particular *i-type* group, each group-member will share the same {*E*_*i*_} family of sufficient exposures as the *i*^*th*^ individual.

Notably, also, the probability, *P*({*E*_*i*_}), depends entirely upon the actual environmental conditions that prevail during any *Time Period* – i.e., conditions that are fixed for any specific (*E*_*T*_). Thus:

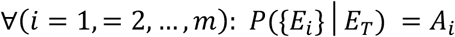

where (*A*_*i*_) represents an unknown constant. This constant (*A*_*i*_) may be different for each {*E*_*i*_} and, also, it may be different during different *Time Periods*. Consequently, during any (*E*_*T*_), each {*E*_*i*_} represents a *population-wide* exposure – i.e., an exposure that is “*available*” to everyone. However, whether anyone, in particular the *i*^*th*^ susceptible individual, experiences that exposure, is a different matter (*see below*).

Also, for MS to develop in the *i*^*th*^ susceptible individual, the events {*E*_*i*_} and (*G*_*i*_) must occur jointly – i.e., the individual (*G*_*i*_) must experience at least one of the {*E*_*i*_} environments. This joint occurrence is represented by the subset ({*E*_*i*_}, *G*_*i*_). The occurrence of ({*E*_*i*_}, *G*_*i*_) represents the event that an individual, randomly selected from (*Z*), is both in the (*G*_*i*_) subset (*described above*) and that they experience an environment sufficient to cause MS in them. The probability of this event, given that this person is a member of (*G*) subset and given that they are living during (*E*_*T*_), is represented as *P*({*E*_*i*_}, *G*_*i*_|*G, E*_*T*_). If the event (*G*_*i*_) occurs without {*E*_*i*_}, then whatever exposure does occur, it is insufficient, and the *i*^*th*^ individual cannot develop MS. However, the relationship between one individual’s family of sufficient exposures to that of others may be complex. For example, every individual (or *i-type*) may have a family with sets unique to them or, alternatively, the families for any two or more individuals (not in the same *i-type* group) may overlap to any degree, even to the point where their families are identical. If *<u>every</u>* susceptible individual has an *<u>identical</u>* family of sufficient environmental exposures, then: ∀(*i*): *P*({*E*_*i*_}, *G*_*i*_|*G, E*_*T*_) = *P*({*E*_*i*_}|*G, E*_*T*_). If some individuals can develop MS under *<u>any</u>* environmental condition, then, for these individuals: *P*({*E*_*i*_}, *G*_*i*_|*G, E*_*T*_) = *P*(*G*_*i*_|*G, E*_*T*_). And, finally, if there are (*s*_*e*_) specific sets of environmental exposure (*e* = 1,2, …, *s*_*e*_) that are sufficient to cause MS in *<u>any</u>* susceptible individual, then, for the family {*E*_*e*_} of these sets of environmental exposure:

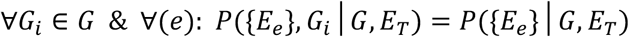

##### Definition of the Exposure (E)

Although an individual (genotype) may experience more than one set of exposures, which may be part of one or more than one {*E*_*i*_} family, the individual’s total environmental experience is still unique to them. Therefore, we will represent the exposure event of interest (*E*) as the union of the disjoint events, which exhibit the pairing of susceptible individuals with sufficient environments, such that:

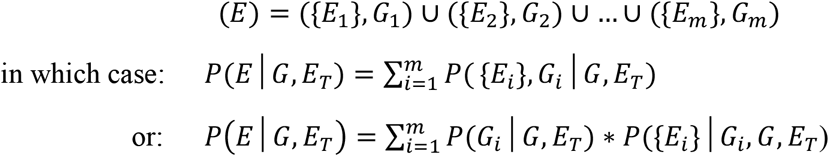

Because genotype is assumed to be independent of the prevailing environmental conditions (*E*_*T*_):

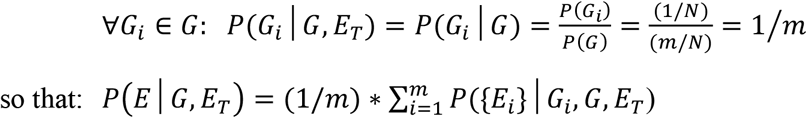

Thus, the term *P*(*E*|*G, E*_*T*_) represents the probability of the event that a member of the (*G*) subset, selected at random, will experience an environmental exposure sufficient to cause MS, given their unique genotype and given the prevailing environmental conditions of the time (*E*_*T*_). Furthermore, from the definition of (*E*), this event can only occur when the event (*G*) also occurs, so that:

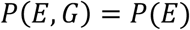

Notably, many environmental factors or events, which are part of a set within the {*E*_*i*_} family, may (and likely do) represent a range of environmental experiences. For example, suppose that Vitamin D deficiency is a factor in one such a set of sufficient exposures. Suppose further that, for the *i*^*th*^ susceptible individual to develop MS in response to this set of exposures, they need to experience a vitamin D deficiency of some minimum severity, lasting for some minimum amount of time, and occurring during some critical age-window. In this case, the definition for the environmental event of “vitamin D deficiency” would also include deficiencies of the same (or greater) severity, lasting the same (or a longer) amount of time, and occurring during the same (or narrower) age-window.

Importantly, as noted previously, each set of sufficient environmental exposures is unspecified as to: 1) how many events or factors are involved; 2) when, during the life of an individual, these events or factors need to occur; and 3) what these events or factors are. Each of these sets, of whatever they consist, simply needs to be sufficient, by themselves, to cause MS to develop in the *i*^*th*^ (or an *i-type*) susceptible individual.

##### Partitioning the Environmental Exposure

In addition, any set of environmental exposures, for any individual, can be partitioned conceptually into three mutually exclusive subsets, which we term: (*E*_*pop*_, *E*_*sib*_, and *E*_*twn*_). The subset (*E*_*pop*_) includes all those environmental experiences or events equally likely to be shared by the population generally (including siblings and twins). The occurrence of (*E*_*pop*_) represents the event that a specific environmental event or factor, which an individual experiences, is a member of the (*E*_*pop*_) subbset. The subset (*E*_*sib*_) includes all those environmental experiences or events either more or less likely to be shared by siblings (including twins) compared to the general population. The occurrence of (*E*_*sib*_) represents the event that a specific environmental event or factor, which an individual experiences, is a member of the (*E*_*sib*_) subset. Presumably, the (*E*_*sib*_) environmental experiences occur mostly (but not necessarily exclusively) during childhood. The subset (*E*_*twn*_) includes all those environmental experiences or events more or less likely to be shared by *MZ*- and *DZ-*twins compared both to non-twin co-siblings and to the general population. The the occurrence of (*E*_*twn*_) represents the event that a specific environmental event or factor, which an individual experiences, is a member of the (*E*_*twn*_) subbset. Presumably, the (*E*_*twn*_) environmental events occur mostly (but not necessarily exclusively) during the intrauterine and early post-natal periods. Importantly, creating this partition does not imply that any of these experiences are unique to twins or siblings – *everyone* experiences each environmental component. The difference is that twins and siblings are more or less likely to *share* certain experiences.

For example, each of the (*v*_*i*_) sets of sufficient environmental exposures within the {*E*_*i*_} family can be partitioned into these three distinct events such that the event (*E*_*ij*_) represents the union of these disjoint events. In this circumstance, therefore:

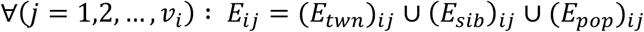

where the probability of each event (*E*_*ij*_) is defined as the the joint probability of these three independent component events so that:

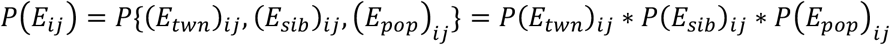

and the event {*E*_*i*_} is represented as:

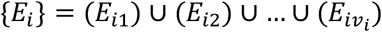

The same is true within every {*E*_*i*_} family – i.e., ∀(*i*): (*i* = 1,2, …, *m*).

{*NB: Many, perhaps most, environmental exposures are “population-wide” in the sense that the risk of these events is shared by everyone. For example, the amount of sunlight reaching the Earth’s surface in a particular region can be considered a “population-wide” exposure in the sense that the same amount of sunlight is “available” to everyone in that region. Despite this, however, there may be certain individuals or certain subgroups within the population who experience less sun-exposure than others (e*.*g*., *if they disproportionately use sun-screen, if they disproportionatley avoid the sun, or if they are otherwise disproportionatley protected from sun-exposure). Conversely, there may also be certain individuals or groups who experience more sun-exposurre than others. However, given the fact that a co-twin (or a non-twin co-sibling) experiences such an imbalance, unless their proband twin (or proband sibling) is either more or less likely to to experience a similar imbalance compared to others, then these exposures would still be considered part of the* (*E*_*pop*_) *environment. Also, the* (*E*_*sib*_) *environment may include experiences outside the childhood micro-environment if, for example, sharing the same biological mother made the intra-uterine environment more similar for siblings than that for the general population. In addition, if twins disproportionately shared certain childhood or adult experriences more so than other siblings or the general population, then these experiences woud be part of the* (*E*_*twn*_) *environment*.

*Although it is unspecified as to what experiences consitiute each subset, nevertheless, these three subsets of environmental exposure* (*E*_*twn*_, *E*_*sib*_, *and E*_*pop*_) *are envisioned to be mutually exclusive and that, together, they comprise any idividual’s unique environmental experience. Thus, as noted above, every individual experiences each of these components of enviornmental exposure, regardless of whether they are twins or non-twin siblings and regardless of whether they are members of the* (*G*) *subset. For example, even though the <u>same</u> intrauterine environment is shared by twins, everyone experiences <u>some</u> intrauterine environment. Similarly, although both twins and non-twin co-siblings experience a <u>similar</u> childhood environment, everyone experiences <u>some</u> childhood environment. Nevertheless, in considering these components of environmental exposure as they relate to the sufficent sets as described above – i*.*e. for* (*i* = 1,2, …, *m*) *and for* (*j* = 1,2, …, *v*_*i*_), *we are here focused on the events* (*E*_*ij*_), *for which, during any Time Period, it will be the case that:*

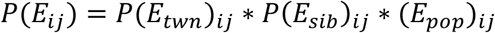

*Notably, during any specific Time Period* (*E*_*T*_), *both P*(*E*_*ij*_) *and its component parts are constants*.}

##### Impact of the (E_sib_) Environment

Despite this conceptual framework, however, the observations from Canada in adopted individuals, in siblings and half-siblings raised together or apart, in conjugal couples, and in brothers and sisters of different birth order, have indicated that MS-risk is not affected by the familial micro-environment but suggest, rather, that the important environmental risks (not considering twins) result from exposures that are experienced *population-wide* [9-15]. Thus, these studies, collectively, provide compelling evidence for the absence of any (*E*_*sib*_) environmental impact on MS.

##### Relationships between (MS), (E), and (G)

From the definitions of environmental and genetic susceptibility (*above*), for the event of (*MS*) to occur, both the event (*G*) and the event (*E*) must also occur. If either of these events does not occur, the event (*MS*) cannot occur. Therefore, it is clear that:

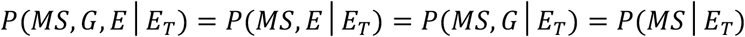

#### C. Circumstances Relating to Twins and Siblings of Individuals with MS

The terms (*MZ*), (*DZ*), and (*S*) represent, respectively, the subsets of *MZ-*twins, *DZ-*twins, and non-twin sib-ships within (*Z*). The occurrence of (*MZ*), (*DZ*), or (*S*) represents the event that an individual, selected at random from (*Z*), belongs, respectively, to each of these subsets and the terms *P*(*MZ*), *P*(*DZ*), *P*(*S*) represent the respective probabilities of these events. For clarity, the randomly selected individual is always referred to as the “proband twin” or the “proband sibling” depending upon the subset to which they belong. The other member (or members) of the twinship or sibship are always referred to as the “co-twin(s)” or “co-sibling(s)”.

##### Circumstances for Twins and Siblings of Selected Probands

Initially, we will consider two events for an *MZ* twin-pair. The first is the event that the randomly selected proband is a member of the (*MS, MZ*) subset and their co-twin is a member of the (*MZ*) subset; the second is the event that the proband is a member of the (*MZ*) subset and their co-twin is a member of the (*MS, MZ*) subset. Clearly, the probability of these two events is the same. Therefore, to distinguish the circumstances of the proband from those of the co-twin, we will use the term the term (*MZ*_*MS*_) to indicate, specifically, the status of the co-twin. Thus, during the *Time Period* (*E*_*T*_):

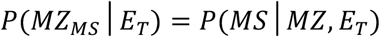

where {*P*(*MS*|*MZ, E*_*T*_)} represents the probability of the event (*MS*) in the proband twin during (*E*_*T*_) and {*P*(*MZ*_*MS*_|*E*_*T*_)} represents the same probability for the event (*MS*) in the co-twin during (*E*_*T*_). Also, because any two *MZ-*twins have “identical” genotypes, therefore:

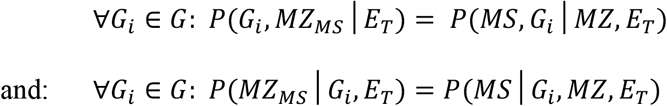

In which case:

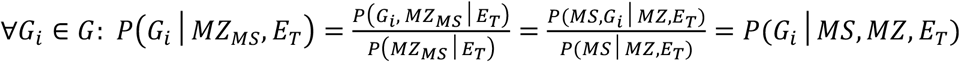

Every proband who has an *MZ* co-twin in the (*MS, MZ*) = (*MS, MZ, G*) subset and who, thus, shares an “identical” genotype with their co-twin, will also be a member of the (*G*) subset. Therefore, summing over all susceptible individuals:

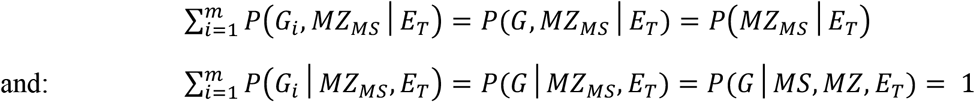

Similarly, the term *P*(*MZ*_*E*_) represents the probability of the event that the co-twin of an *MZ-* proband, randomly selected from (*Z*), is a member of the (*MZ, E*) subsets, respectively. Thus

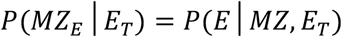

In an analogous manner, for *DZ-*twinships, the status of the co-twin is indicated by the subsets and the events of: (*DZ*_*MS*_) and (*DZ*_*E*_). And for non-twin sibships, the status of the co-sibling is indicated by the subsets and events of: (*S*_*MS*_) and (*S*_*E*_).

Thus, the two terms, *P*(*MS*|*MZ*_*MS*_, *E*_*T*_) and *P*(*MS*|*DZ*_*MS*_, *E*_*T*_) represent the conditional life-time probability of the event that an individual (the proband), randomly selected from (*Z*), is a member of the either the (*MS, MZ*) or the (*MS, DZ*) subset, given the fact that their co-twin also belongs, respectively, to the (*MS, MZ*) or the (*MS, DZ*) subset, and given the prevailing environmental conditions of the time (*E*_*T*_). These probabilities are estimated by the proband-wise concordance rate for either *MZ*- or *DZ*-twins [16]. This rate is estimated based on the number of concordant twin-pairs (*C*_*T*1_) compared to the number of discordant twin-pairs (*D*_*T*1_) and adjusted based upon the degree to which twins are “doubly ascertained”. The term “doubly ascertained”, in this context, represents the proportion of twin-pairs, for whom both twins were independently identified by the initial ascertainment scheme [16]. If all twin-pairs are “doubly ascertained” by this scheme, and if the sample from (*Z*), so ascertained, is random, then the formula for calculating the proband-wise concordance rate is:

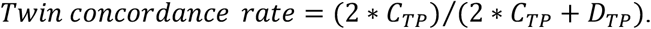

However, if the probability of “double ascertainment” is less than unity, then this formula requires some modification [16]. In the Canadian data [6] the double-ascertainment rate for concordant *MZ-*twins was 48.5% (16/33).

In a similar manner, the term *P*(*MS*|*S*_*MS*_, *E*_*T*_) represents the conditional life-time probability of the event that an individual (the proband), randomly selected from (*Z*), is a member of the (*MS, S*) subset, given the fact that one or more of their non-twin co-siblings is a member of the (*MS, S*) subset and given prevailing environmental conditions of (*E*_*T*_).

#### D. Adjustments for the Shared Environment of Twins

Lastly, the term *P*(*MS*|*IG*_*MS*_, *E*_*T*_), represents the proband-wise concordance rate for *MZ*-twins during (*E*_*T*_) – i.e., *P*(*MS*|*MZ*_*MS*_, *E*_*T*_) – which has been adjusted for the fact that concordant *MZ*-twins, in addition to sharing their “identical” genotypes (*IG*), also disproportionately share their (*E*_*twn*_) and (*E*_*sib*_) environments with each other. Such an adjustment may be necessary because, if these disproportionately shared environmental experiences contribute to causing MS in the co-twin, they could also increase the likelihood of MS developing in the proband twin and such a circumstance could, potentially, alter our conclusions regarding the nature of genetic susceptibility in the population.

Because, by definition (*see above*):

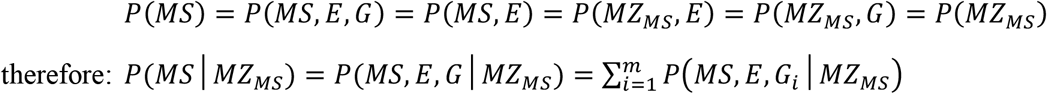

where: ∀(*i*): (*i* = 1,2, …, *m*):

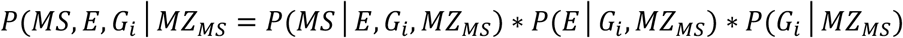

In this way, the probability of the proband being a member of the (*MS, E, G*_*i*_) subset, given the fact that their co-twin is a member of the (*MS, MZ*) subset, is expressed as: *P*(*MS, E, G*_*i*_|*MZ*_*MS*_). This probability can then be re-expressed as the product of three component probabilities – the probability of MS developing in an *MZ-*proband (*G*_*i*_) who experiences a sufficient environment, the probability that this *MZ-*proband experiences an environment sufficient to cause MS, and the probability of this *MZ-*proband being a member of the (*G*_*i*_) subset – where each probability is conditioned on the proband having an *MZ* co-twin, who is a member of the (*MS, MZ*) subset within (*Z*).

For susceptible probands who are members of (*G*), but who may or may not be from twinships or sibships where the co-twin or a co-sibling is a member of the (*MS*) subset, the analogous probabilities can be written:

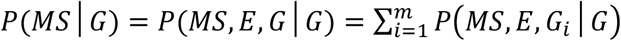

where: ∀(*i*): (*i* = 1,2, …, *m*):

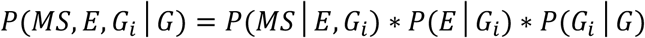

Therefore, to estimate the desired “adjusted” probability, *P*(*MS*|*IG*_*MS*_), we need to remove the impact of the shared environment of *MZ-*twins, while leaving the genetic impact of being *MZ-*twins unchanged. Thus, we define:

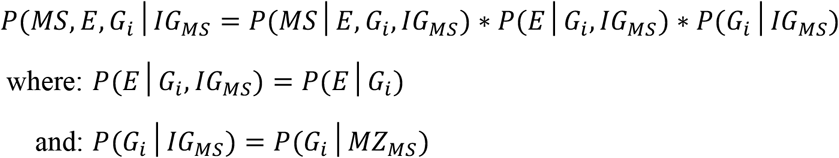

Moreover, we note that the conditioning events (*E, G*_*i*_) and (*E, G*_*i*_, *MZ*_*MS*_) both represent the same underlying event for the proband – i.e., the event that the *i*^*th*^ susceptible individual (the *MZ-*proband) experiences an environment sufficient to cause MS in them. In this circumstance, therefore:

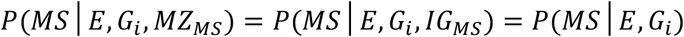

Incorporating this equivalence, into the *above* definitions, yields:

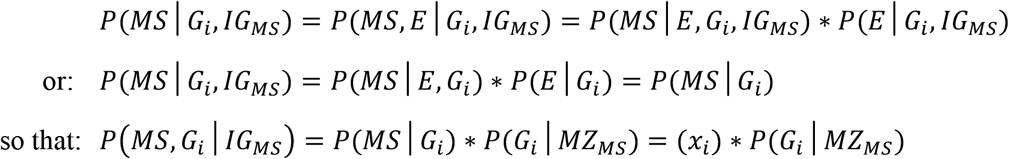

Therefore, summarizing the *above* arguments, the term (*IG*_*MS*_) is defined such that:

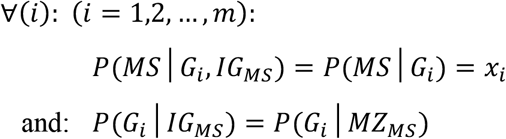

And, therefore, the desired “adjusted” probability, *P*(*MS*|*IG*_*MS*_), can be expressed such that:

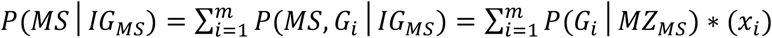

Effectively, this adjustment equates to a thought-experiment, in which susceptible *MZ-*twins are separated at conception, and in which the proband twin experiences random (*E*_*twn*_), (*E*_*sib*_), and (*E*_*pop*_) environments, just as would any other member of the (*G*) subset, given the environmental conditions of the time (*E*_*T*_).

In addition, as discussed previously (*Methods #1B*), the available evidence suggests that the (*E*_*sib*_) environment has no impact on the likelihood of a susceptible individual subsequently developing MS [9-15]. If this evidence is correct, it will also be the case that:

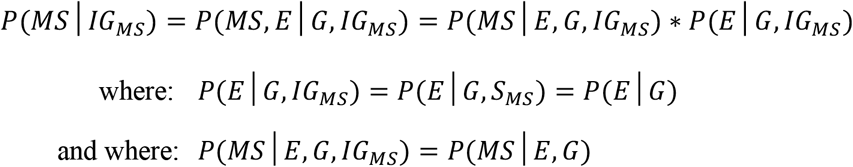

which represents the probability that the event (*MS*) occurs for an individual, randomly selected from (*Z*), given that they are both a member of the (*G*) subset and that they have an environmental experience sufficient to cause MS in them.

{*NB: This definition represents the intended meaning of the “adjusted” proband-wise recurrence rate for MZ-twins – i*.*e*., *P*(*MS*|*IG*_*MS*_). *The next section (which can be skipped) is devoted to estimating an adjustment that matches this definition and that is derived from directly observed epidemiological data*.}

##### Estimating the Necessary Adjustment from Observed Data

Here the event of interest is the development of MS in a randomly selected proband *MZ-*twin, whose co-twin already has or will develop MS – i.e., the co-twin of this proband is a member of the (*MZ*_*MS*_) subset. Consequently, in this conceptualization, the event (*IG*_*MS*_), which is independent of whatever happens to the proband, is envisioned to be identical to the event (*MZ*_*MS*_), defined *above*. Therefore, during any specific *Time Period*, (*E*_*T*_):

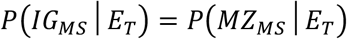

The difference, as discussed *above*, is that the probability of the event – i.e., *P*(*MS*|*IG*_*MS*_) – may be different from; *P*(*MS*|*MZ*_*MS*_) due to the shared environmental experiences of *MZ-*twins.

Including the circumstances of both twins and siblings, the occurrences of: (*MZ*_*E*_), (*DZ*_*E*_), and (*S*_*E*_) represent, respectively, the events that a co-*MZ-*twin, a co-*DZ-*twin, or a non-twin co-sibling, experience the exposure (*E*). The respective probabilities that the probands of these co-twins or co-siblings are members of the (*G*) subset and that they experience {*E*_*i*_} are expressed as: *P*({*E*_*i*_}|*MZ*_*E*_); *P*({*E*_*i*_}|*G, DZ*_*E*_); and: *P*({*E*_*i*_}|*G, S*_*E*_). Moreover, because any two siblings and any set of *DZ-*twins have the same genetic relationship to each other, and because, by definition, they both disproportionately share the same (*E*_*sib*_) environment, the only difference between them is that *DZ-*twins share the same (*E*_*twn*_) environment whereas siblings don’t.

These probabilities are then contrasted to the probability of the event {*E*_*i*_} for a randomly selected member of the (*G*) subset – i.e., *P*({*E*_*i*_}|*G*). As *above*, we can partition each of these exposure events into three independent component events (*E*_*twn*_, *E*_*sib*_, & *E*_*pop*_), during any *Time Period*, such that:

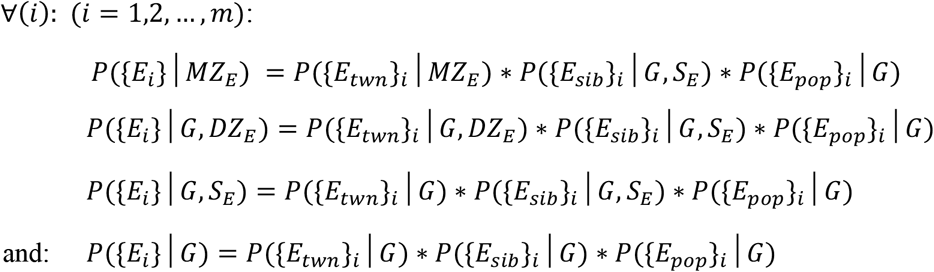

Probands both from an *MZ-*twinship and those from a *DZ-*twinship experience the same (*E*_*twn*_) environment as their co-twin who either has or will develop MS. Therefore, we assume that:

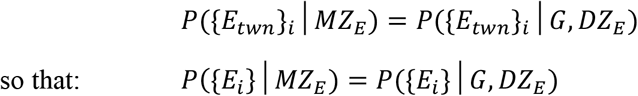

In this circumstance:

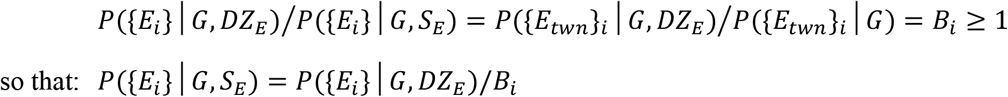

where (*B*_*i*_) is a constant for each (*i*) during any specific *Time Period* (*E*_*T*_).

The terms *P*(*MS, G*_*i*_, {*E*_*i*_}|*G, DZ*_*E*_) and *P*(*MS, G*_*i*_, {*E*_*i*_}|*G, S*_*E*_) represent, respectively, the probabilities of the events that the *i*^*th*^ individual, randomly selected from (*Z*), is a member of either the (*MS, DZ*) or the (*MS, S*) subsets, given that they are a member of the (*G*) subset and also given that their co-twin or co-sibling is a member of either the (*DZ, E*) or the (*S, E*) subsets. However, by definition (*see Methods #1B*), if both events (*G*) and {*E*_*i*_} occur together, then the event (*MS, G*_*i*_) will either occur or not occur, independently of the status of a co-twin or co-sibling.

Therefore, ∀(*i*): (*i* = 1,2, …, *m*):

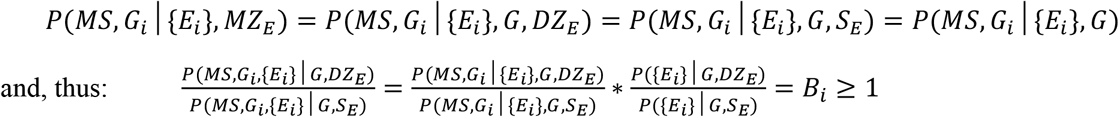

In this manner, we can use the adjustment factor (*B*_*i*_), to remove, mathematically, the impact of the shared {*E*_*twn*_} environment for the *i*^*th*^ individual, during any *Time Period*, such that:

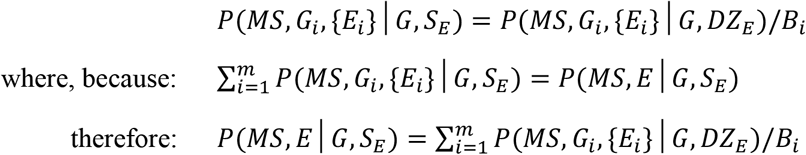

In this case, we can assign variables (*y*) and (*z*) such that:

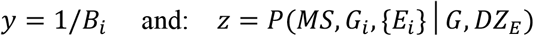

In which case, the covariance (*σ*_*yz*_), using its standard definition [39], can be expressed such that:

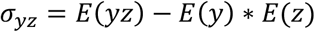

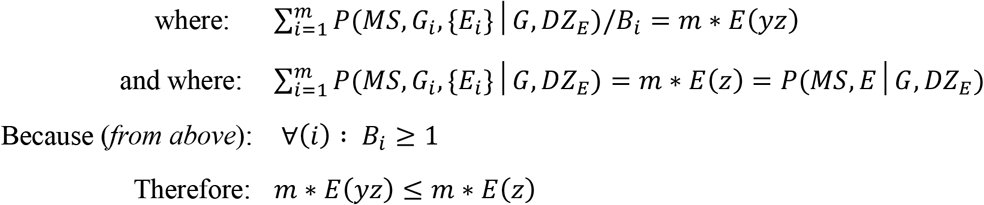

In this circumstance, we can define an adjustment factor (*s*_*a*_) such that:

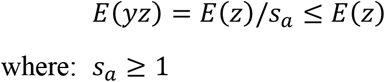

Substituting this back into the definition (*above*) for the covariance (*σ*_*yz*_), yields:

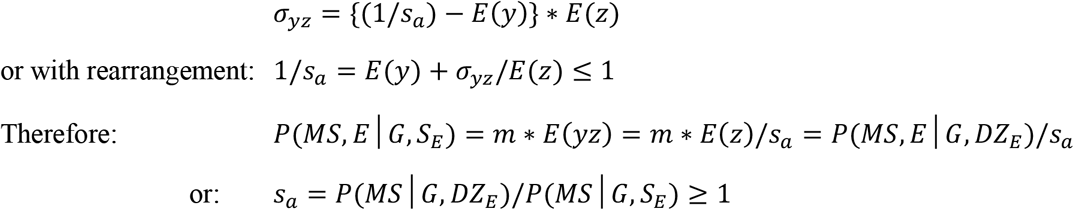

We define the term (*IG*_*E*_) in an analogous manner to (*MZ*_*E*_) – *see above*. Moreover, because everyone who develops MS must experience the event (*E*), because both *MZ-* and *DZ-*twins disproportionately share the (*E*_*twn*_) and (*E*_*sib*_) environments with their co-twin, and because siblings (including twins) disproportionately share their (*E*_*sib*_) environment with their co-sibling(s), and because everyone shares their (*E*_*pop*_) environment, therefore, with respect to the environmental experiences of the proband, it will be the case that:

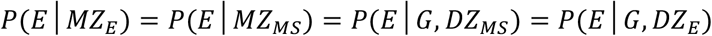

Moreover, from the definition of (*IG*_*MS*_) and because the (*E*_*sib*_) environment doesn’t seem to contribute to the event (*MS, E*) and because genotype is independent of (*E*_*T*_), then, in this circumstance:

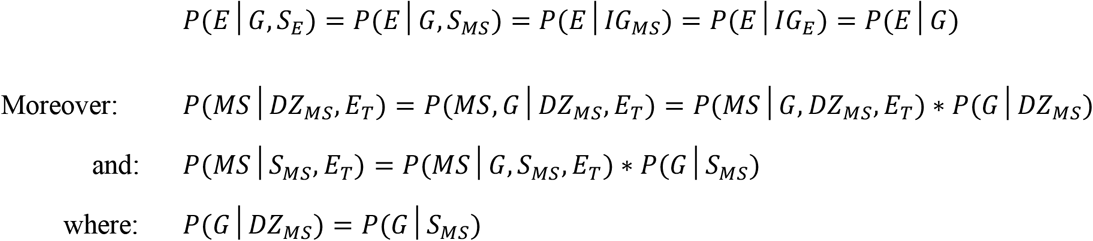

Therefore, to convert *P*(*MS*|*MZ*_*MS*_, *E*_*T*_) into *P*(*MS*|*IG*_*MS*_, *E*_*T*_), we can remove the impact of the shared (*E*_*twn*_) environment of *MZ-*twins, during (*E*_*T*_), using observable population parameters such that:

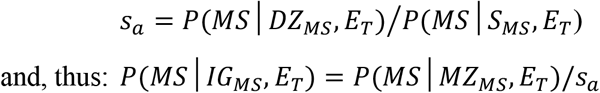

{*NB: In this development of the adjustment factor* (*s*_*a*_), *we have assumed that the* (*E*_*twn*_) *environment is the same for MZ- and DZ-twins. We have also assumed, based on evidence [9-15], that the* (*E*_*sib*_) *environment doesn’t impact the likelihood of a person subsequently developing MS. However, either one (or both) of these assumptions may be incorrect. For example, certain* (*E*_*twn*_) *exposures may be more or less likely to occur in MZ-twins compared to DZ-twins, in which case our estimate for* (*s*_*a*_) *would be either an under or an overestimate. Nevertheless, if either the* (*E*_*twn*_) *or the* (*E*_*sib*_) *environments are important to MS pathogenesis, then some adjustment is necessary. It is for this reason that, for our analysis, we consider a wide range of possible values for* (*s*_*a*_*), including those circumstances for which no adjustment is necessary – see below*.}

#### E. Characterizing Genetic Susceptibility to MS in a Population

From these *Model* specifications and definitions, we can use estimated values for observable population parameters to deduce the value of the non-observable parameter *P*(*MS*|*G, E*_*T*_), which represents the probability of the event that an individual, randomly selected from (*Z*), will develop MS over the course of their lifetime, given that they are a member of the (*G*) subset and given the prevailing environmental conditions of (*E*_*T*_). From the definition of (*G*), as noted earlier:

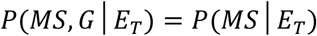

Therefore, because genotype is assumed to be independent of the prevailing environmental conditions of (*E*_*T*_):

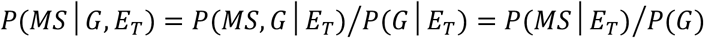

Rearrangement of this equation, yields:

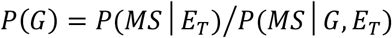

Consequently, the value of *P*(*G*) can be estimated using the data from *<u>any</u>* specific *Time Period* (*E*_*T*_) – including ours – during which: *P*(*MS*|*E*_*T*_) > 0. Therefore, considering only our “current” *Time Period*, this equation can be simplified to yield:

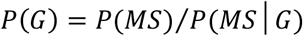

Moreover, once the value of *P*(*G*) is established, it can then be used to assess the nature of MS pathogenesis. For example, if: {*P*(*G*) = 1} – i.e., if the penetrance of MS for the population (*Z*) is the same as that for the subset (*G*) – then anyone can get MS under the appropriate environmental conditions. By contrast, if: {*P*(*G*) < 1} – i.e., if the penetrance of MS for the population (*Z*) is the less than that for the subset (*G*) – then only certain individuals in the general population (*Z*) have any possibility of getting MS. In this circumstance, MS must be considered a “genetic” disorder (i.e., unless a person has the appropriate genotype, they have no chance, whatsoever, of getting MS, regardless of their environmental exposure). Importantly, even if MS is “genetic” in this sense, this has no bearing upon whether disease pathogenesis also requires the co-occurrence of specific environmental events.

In this analysis, two basic *Models* are used to estimate the values of various unknown epidemiological parameters of MS. The first *Model* takes a cross-sectional approach, in which deductions are made from the epidemiological data obtained during a single *Time Period* (i.e., the “current” *Time Period*). This will be referred to as the *Cross-sectional Model*. The second *Model* takes a longitudinal approach, in which deductions are made both from the “current” epidemiological data and from the observed changes in MS epidemiology that have taken place over the past 4–5 decades [3,6]. This will be referred to as the *Longitudinal Model*. These two *Models* are independent of each other although both incorporate many of the same observed and non-observed epidemiological parameters, which are important for MS pathogenesis. The *Cross-sectional Model* derives theoretical relationships between different epidemiologic parameters, but it also makes two assumptions regarding *MZ*-twin data to establish these relationships (*see below; Methods #3*). These two assumptions are also commonly made by other studies, which analyze *MZ-*twin data, and each has observational data to support them [17-19]. Nevertheless, both for *Assertions A–D* and for *Equations 4a* and *4b* (*see below*), these conditions need to be assumed. By contrast, the *Longitudinal Model* does not make either of these assumptions to estimate possible ranges for the non-observed parameters and several possible conditions for this *Longitudinal Model* are depicted in *Figures 1–4* (*see below*).

**Figure 1.**
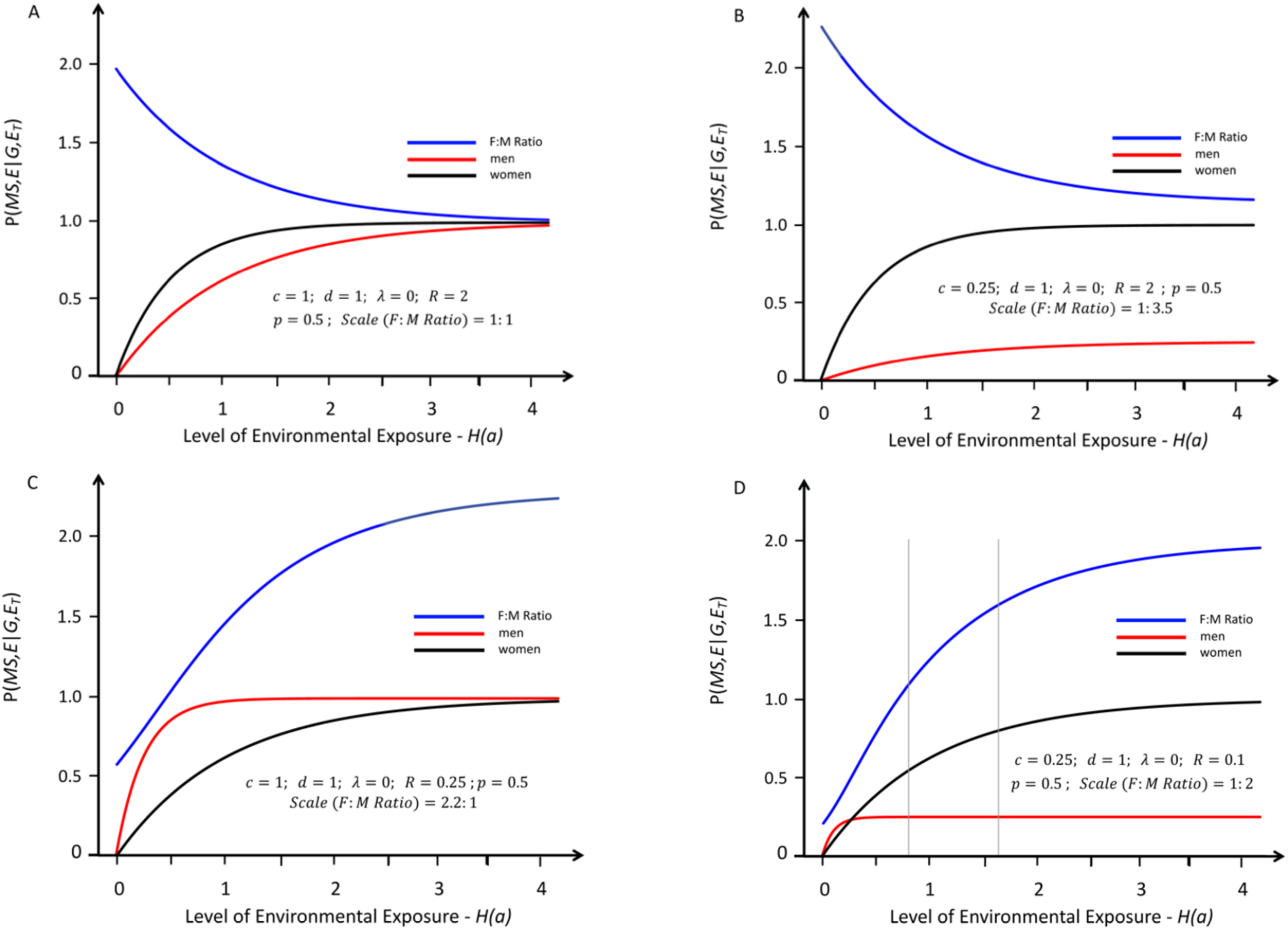
Response curves representing the likelihood of developing MS in genetically susceptible *women* (black lines) and *men* (red lines) with an increasing probability of a “sufficient” environmental exposure – *see Methods #1B*. The curves depicted are “strictly” proportional, meaning that the environmental threshold is the same for both *men* and *women* – i.e., under conditions in which: (*λ* = 0) – *see Text*. The blue lines represent the change in the (*F:M*) *sex ratio* (plotted at various scales) with increasing exposure. The thin grey vertical lines represent the portion of the response curve that covers the change in the (*F:M*) *sex ratio* from 2.2 to 3.2 (i.e., the actual change observed in Canada [6] between *Time Periods #1 & #2*). The grey lines are omitted under circumstances either where these observed (*F:M*) *sex ratios* are not possible or where both (*Zw* > *Zm*) and an increasing (*F:M*) *sex ratio* are not possible. Response curves *A* and *B* reflect conditions in which (*R* > 1); whereas curves *C* and *D* reflect conditions in which (*R* < 1). If (*R* = 1), the blue line would be flat. Response curves *A* and *C* reflect conditions in which (***c*** = ***d*** = 1); whereas curves *B* and *D* reflect those conditions in which (***c*** < ***d*** = 1). Under the conditions for curves *A* and *B* (*R* ≥ 1), there is no possibility that the (*F:M*) *sex ratio* will be observed to increase with increasing exposure. Under the conditions of curve *C* – i.e., (***c*** = ***d*** = 1) and (*R* < 1) – at no exposure level is it possible that:

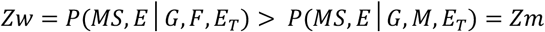

Thus, the only “strictly” proportional model that could possibly account for an increasing (*F:M*) *sex ratio*, and for the fact that: (*Zw*_2_ > *Zm*_2_), is a *Model* in which (***c*** < ***d*** ≤ 1) and (***R*** < 1) – i.e., curve *D*.

For both *Models*, the first step is to assign acceptable ranges for the value of certain “observed” parameters (e.g., twin and sibling concordance rates, the population prevalence of MS, or the proportion of *women* among MS patients). These ranges are assigned such that they always include their calculated 95% *CIs*. However, for certain parameters, the ranges considered plausible are expanded beyond the limits set by the *CIs*. The second step is to assign acceptable ranges for the “non-observed” parameters (e.g., the proportion of susceptible persons in the population or the proportion of *women* among susceptible individuals). These ranges are assigned such that they cover the entire “plausible” range for each such parameter. In both *Models*, a “substitution” analysis is undertaken to determine those parameter combinations (i.e., solutions), which fit within the acceptable ranges for both the observed and non-observed parameters. These solutions are then used to assess their implications about the basis of genetic and environmental susceptibility to MS in the population. For each *Model*, the total number of parameter combinations interrogated in this manner was ∼*10*^*11*^.

### 2. Establishing Plausible Ranges for Parameter Values

#### A. Observed Parameter Values

For notational simplicity, we sometimes use subscripts (*1*) and (*2*) to indicate the parameter values at *Time Period #1* and *Time Period #2* {e.g., *P*(*MS*)_2_ = *P*(*MS*|*E*_*T*_) at *Time Period #2*}. For the purposes of this analysis, those parameter-values observed for persons born between 1976 and 1980 (i.e., *Time Period #2*) are always taken to be the “current” values and, when only this *Time Period* is being considered, the terms (*E*_*T*_) and the subscript (*2*) are often omitted entirely to simplify the notation.

{*NB: In general, for individuals born during Time Period #2 (1976–1980), their MS status cannot be determined until 25-35 years later (i*.*e*., *2001–2015). The estimates of other epidemiological parameters are from reports in the Time Period of (2001–2015), which is also when the Time Period #2 (F:M) sex ratio is assessed [6,7,11-15]. For this reason, Time Period #2 is fixed as the “current” period. However, because the (F:M) sex ratio has increased between every previous 5-year epoch and Time Period #2 [6], the choice of any specific Time Period #1 is equivalent to any other. For our Time Period #1, we chose the 5-year epoch (1941–1945) because it was the earliest epoch with the narrowest CI [6]*.}

The 2010 Canadian census [20], reported that the proportion of *women* among the general Canadian population (*Z*) is 50.4%. Thus, *men* and *women* comprise essentially equal proportions of this population and, therefore, the probabilities of the events that an individual, randomly selected from (*Z*), is a *woman* or a *man* – *P*(*F*) and *P*(*M*) respectively – are each ∼50%. Therefore, by definition:

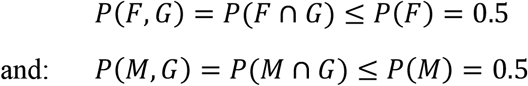

Currently, the proband-wise concordance rate [16] for MS in *MZ*-twins, observed in Canada, is:

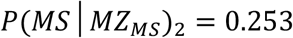

with (*n* = 146) twin-pairs included in this estimation [7]. Therefore, the 95% *CI* for this parameter, calculated from an exact binomial test [21], is:

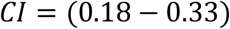

The estimates, from different studies, for the “current” proportion of *women* among the MS patients – i.e., *P*(*F*|*MS*)_2_ − in North America ranges between 66% and 76% [3]. For the *Cross-sectional Model*, we expanded the “plausible” range beyond the 95% *CI* calculated from “current” Canadian data presented *below* [6]. Thus, for this *Model*, we considered the range:

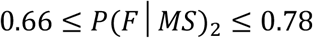

The reason for this is because the “current” estimated range from the Canadian study is quite narrow and some solutions, which fall within the range of different estimates from other locations in North America [3], might be excluded. This choice permits a wider range of possibilities to be considered as solutions for our *Cross-sectional Model*.

By contrast, for our *Longitudinal Model*, because we were interested specifically in how the parameter *P*(*F*|*MS*) has changed for the Canadian population over time [6], we used the 95% *CIs* (from this single study) to estimate the ranges for this parameter value during each *Time Period*. For example, the proportion of *women* among the MS patients in Canada was 69% for patients born during *Time Period #1* (1941–1945) and this proportion was significantly less (*p* < 10^−6^) than the 76% observed for patients born during the “current” *Time Period #2* (1976–1980) [6]. Although the authors of this study, do not report the actual numbers of individuals in each 5-year epoch, they do report that their 5-year samples averaged 2,400 individuals per epoch [6]. Also, the authors graphically present the 95% *CIs* for the (*F:M*) *sex ratio* during each of these 5-year epochs in the manuscript *Figure* [6]. Estimating that the number of individuals in both *Time Periods #1* and *#2* is ∼2,000, and using an exact binomial test [21], for our *Longitudinal Model*, we estimate that:

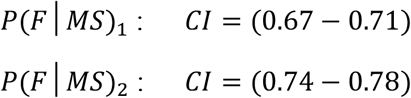

Both of these ranges exceed those (based on the 95% *CIs*) presented in the manuscript *Figure* [6].

The “current” proband-wise concordance rates for MS in *female* and *male MZ*-twins, observed in Canada, are 34% and 6.5%, with the total number of *female* and *male* twin-pairs included in these calculations being (*n*_1_ = 100) and (*n*_2_ = 46), respectively [7]. Using an exact binomial test [21], and using the definitions provided in *Table 2*, the *CIs* for these observations are:

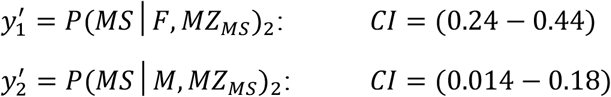

The 95% *CI* for the difference in *MZ-*twin concordance between *men* and *women* is calculated as:

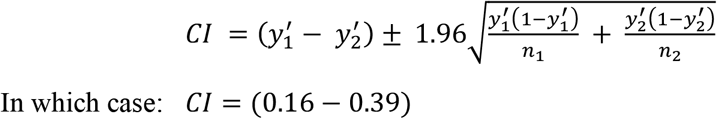

Previously, we used three independent methods (*based on observation*) to estimate the value of *P*(*MS*)_2_ [3]. The first method relied on measures of the population prevalence of MS in North America together with the observed age-distribution for MS-onset, the second method considered the age-specific prevalence of MS in the age-band of 45–54 years, and the third method considered a *population-based* multiple-cause-of-death study from British Columbia, which reported the proportion of death certificates that mention MS. The parameter-value range supported, collectively, by these different methods was: 0.0025 ≤ *P*(*MS*)_2_ ≤ 0.0046 [3]. Nevertheless, for the purposes of the present analysis, we expanded the “plausible” range for this parameter to include:

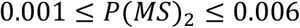

#### B. Initial Constraints on the Non-Observed Parameter Values

In addition to the observed parameter values (*above*), and using the definitions in (*Table 2*), we determined acceptable values for 12 additional parameters: *P*(*G*); *p* = *P*(*F*|G) ; *x* = *P*(*MS*|*G*)_2_ ;

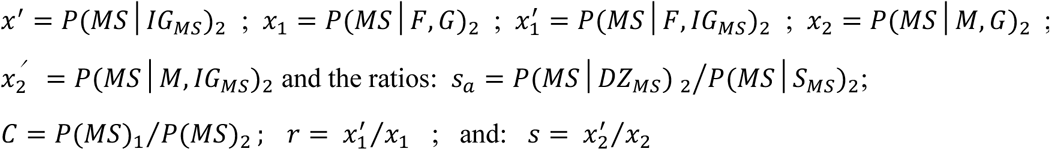

Most of these parameters vary depending upon the level of exposure – i.e., all except *P*(*G*) and *P*(*F*|G). Therefore, the acceptable ranges were estimated for the “current” *Time Period #2*. In several cases, there are constraints on the values that these non-observed parameters can take. For example, *P*(*MS*) has been observed to be increasing, especially (but not only) among *women*, in many parts of the world between the two *Time Periods* [6,22-30]. Therefore, the parameter (*C*) is constrained in three ways such that:

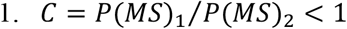

In this case, on theoretical grounds [3], (*C*) is also constrained such that:

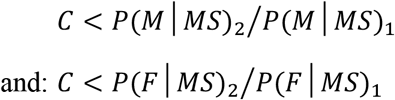

Using the limits, provided earlier, for the proportion of *women* among MS patients during different *Time Periods*, the ratio (*C*) is at its maximum possible value when:

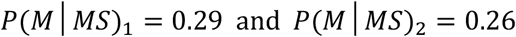

and, therefore, on these theoretical grounds (*see above*), the value of (*C*) is further constrained such that:

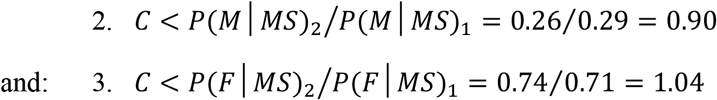

Only the second of these constraints satisfies all three, so that the maximum upper bound for (*C*) is 0.90. Nevertheless, the actual upper bound for (*C*) will depend upon the values that *P*(*M*|*MS*)_1_ and *P*(*M*|*MS*)_2_ take for in any specific solution. Moreover, if (*C* < 0.25) then there must have been a greater than 4-fold increase in *P*(*MS*) for Canada, which has taken place over a 35–40 year-interval. This seems to be an implausibly large increase based on the available data [6,22-30]. Therefore, we conclude that:

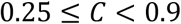

Because MS develops in some individuals, the parameter *P*(*G*) cannot be equal to 0. Also, because both *women* and *men* can develop MS, the parameter *P*(*F*|*G*) cannot be equal to either 0 or 1. Therefore, the plausible ranges for these parameters are:

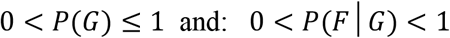

Furthermore, the penetrance of MS for the subsets (*F, G*) and (*M, G*) can be expressed (*see above*) such that:

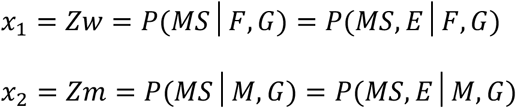

Because everyone who develops MS must be a member of the (*G*) subset, therefore, considering only this subset, the ratio of *women* to *men*, during any *Time Period* can be expressed as:

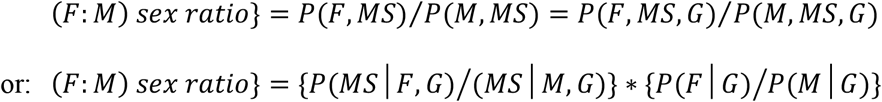

Consequently, using the definitions provided *above* and in *Table 2*, the parameters (*x*_1_), (*x*_2_), (*x*), (*p*) and (*p*^′^), during any *Time Period* for the gender partition, are related such that:

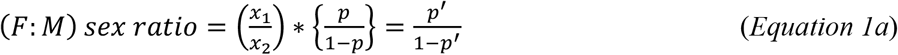

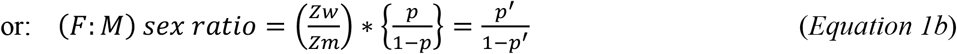

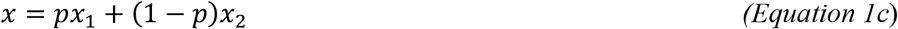

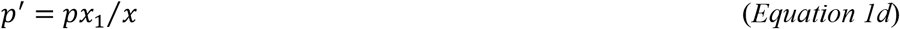

These relationships require no assumptions and (*x*_1_), (*x*_2_), (*x*), (*p*) and (*p*^′^) must <u>always</u> satisfy *Equations 1a–d* during any *Time Period*, regardless of which *Model* is employed in the analysis [3].

#### C. Enrichment of More Penetrant Genotypes

In addition, it can be demonstrated that more penetrant genotypes are “*enriched*” in the (*G, MS*), (*F, G, MS*) and (*M, G, MS*) subsets compared, respectively, to the subsets of susceptible individuals (*G*), (*F, G*) and (*M, G*). For example, during any *Time Period*, consider the two ratios, (*A*) and (*B*), such that:

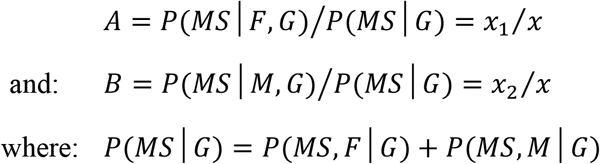

Therefore, during any *Time Period*, both of the following relationships must hold:

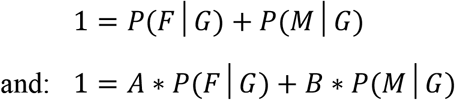

Consequently, during any *Time Period*, it is clear that:

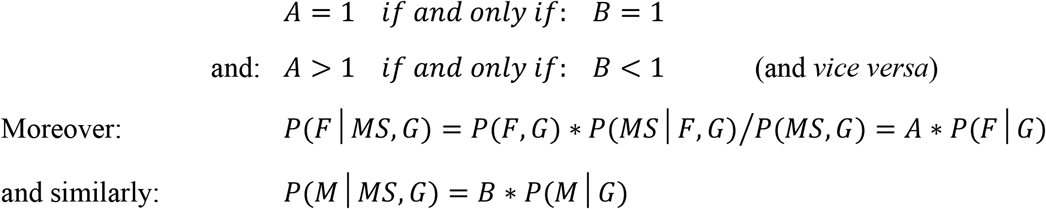

Therefore, if (*A* < 1 < *B*) – i.e., if *men* are more penetrant than *women* – then:

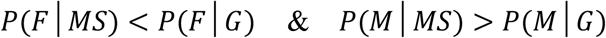

Conversely, if (*A* > 1 > *B*) – i.e., if *women* are more penetrant than *men* – then:

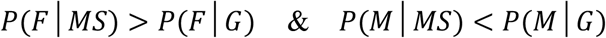

In either case, the more penetrant gender can be described as being “*enriched*” in the (*MS*) subset in comparison to the (*G*) subset.

In addition, considered separately, and partitioning each of the subsets (*F, G*) and (*F, G*) into their high- and low-penetrance sub-subsets, (*G*1^′^) and (*G*2^′^), respectively – *see Methods #1A* – an analogous argument demonstrates that an “*enrichment*” of more penetrant genotypes will also occur within each of these two sub-subsets such that:

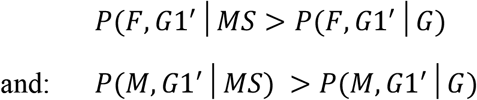

##### Penetrance of MS in Susceptible Men and Women

Importantly, the evidence overwhelmingly favors of the arrangement (*considered above*) in which: (*A* > 1) or, equivalently, in which: (*Zw*_2_ > *Zm*_2_). For example, the (*F:M*) *sex ratio* has increased by a similar amount (i.e., at a similar rate) between every two *Time Periods* in the Canadian data except one [6]. Thus, the observed increase in MS prevalence over time has disproportionately impacted the prevalence in *women* [6,22-30]. Indeed, from *Equation 1b*, during the “current” *Time Period*, it must be that:

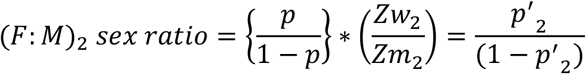

The value of (*p*) is unknown but fixed, regardless of *Time Period*, and the values of (*Zw*_2_) and (*Zm*_2_) are also unknown but fixed during the current *Time Period*. The current value of (*p*^′^) – i.e., (*p*^′^_2_) – by contrast, is both fixed and known (i.e., observed). From *Equations 1c–d*, and also as indicated *above*, the acceptable value-range for the parameter (*p*^′^_2_) is such that:

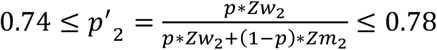

Therefore, considering the implications of *Equation 1b*, the only circumstances in which the (*F:M*) *sex ratio* can increase over time is, also, for the ratio (*Zw*/*Zm*) to increase over time. Certainly, the steady increase in the (*F:M*) *sex ratio* observed in Canada over the last several decades would be most easily explained by those circumstances in which (*p* < *p*^′^) during every *Time Period* represented by this data [6]. Moreover, there are two considerations that strongly support this view. First, the only circumstances both for which this ratio will increase and for which: (*Zm*_2_ ≥ *Zw*_2_), are also those where: (*p* ≥ *p*^′^_2_ ≥ 0.74) and also those where both (*p* ≥ *p*^′^) and (*Zm* > *Zw*) during every previous *Time Period* represented by the Canadian data [7]. In fact, this conclusion would pertain to every observation throughout the history of MS, including a time when the proportion of *men* among MS patients was reported to exceed that of *women* [40], and also including currently, where women account for a large (and increasing) majority of MS patients [3]. Such a conclusion would certainly be counter intuitive.

Second, and notably, other findings from the Canadian study [7] also suggest the possibility that (*Zm*_2_ ≥ *Zw*_2_) is very unlikely. Specifically, this study reported that, for *female* and *male* probands, who have a co-twin in the (*MZ*_*MS*_) subset, the observed penetrance of MS for *women* is (5.7)-fold greater than it is for *men*. Thus, this study [7] reported that:

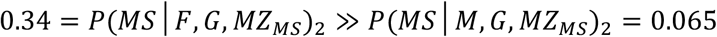

This difference in penetrance between genders is highly significant such that:

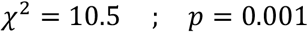

As a consequence of this penetrance difference, and considering *MZ-*twins who either are of unknown concordance or are known to be concordant – i.e., who, respectively, are members of either the (*MZ*_*MS*_) subset or the (*MS, MZ*_*MS*_) sub-subset – this study [7] indicates that *women* are being continuously *enriched* such that:

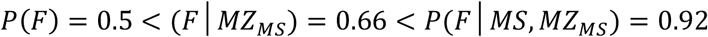

Because any proband *MZ*-twin is genetically “identical” to their (*MZ*_*MS*_) co-twin, the probands of such co-twins are already “*enriched*” for more penetrant genotypes compared to the general population (*see above*). Consequently, the fact that this “*enriched*” group of *women* has a considerably greater penetrance than similarly “*enriched*” *men* (*see below*), strongly suggests that, currently, susceptible *women* are more penetrant than susceptible *men*. Otherwise, for the circumstance of (*Zm*_2_ ≥ *Zw*_2_) to be true, the *enrichment* of more penetrant genotypes among *women*, when going from *P*(*F*|*G*) to *P*(*F*|*MS*), must be substantially greater than it is among *men*, when going from *P*(*M*|*G*) to *P*(*M*|*MS*). Such a circumstance, using the definitions of *Table 2*, requires that the variance of penetrance values within the subset of susceptible *women* considerably exceed that in and susceptible *men –* i.e., this requires that: 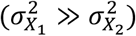 *– see Equations 3a–d –* or, equivalently, it requires that: (*r* ≫ *s*). Either would be hard to rationalize.

Therefore, based on both of these considerations, and without making any assertions regarding the circumstances that might pertain during earlier *Time Periods*, we assume that:

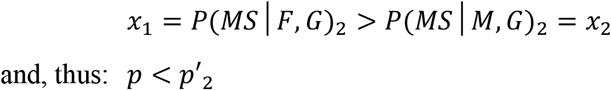

{*NB: If our assumption that, currently*, (*x*_1_ > *x*_2_), *is incorrect, then, as noted above, susceptible men would always have a greater penetrance than susceptible women. Moreover, this would also indicate that men comprise only a small proportion* (≤ 26 %) *of all susceptible individuals or, equivalently, that:* {*P*(*G*|*M*) ≤ 0.36)} *and that:* {*P*(*G*) ≤ 0.68)}. *Nevertheless, in such a case, conditions similar to those depicted in Figure* 1*C (although approaching a sex ratio greater than unity) would be possible. However, using the definitions from the Longitudinal Model (see below and Table 2), and using the substitution analysis (described above), for every solution, where this assumption was violated, we found that:*

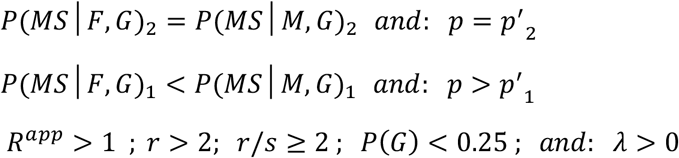

*Notably, if the hazards are proportional, any solution for which* (*p* = *p*^′^_2_) *implies that the response curves in both women and men have already reached their maximum values by Time Period #2 (see Methods #4C; Equation 11d). Such a circumstance further implies that no further increase in the (F:M) sex ratio is possible and, thus, as defined below (see Methods #4A), it also must be that:* (***c*** ≪ ***d***). *For our current Time Period to have precisely coincided with this point in time seems surprising, especially given the fact that the observed sex ratio has been steadily increasing, and at an approximately even rate, up to this point [6]*.

*Also, as it pertains to other aspects of either Model, it is worth noting that, because:* {*P*(*F*) = 0.5}, *<u>any</u> circumstance for which:* (*p* ≠ 0.5), *<u>requires</u> that:* [*P*(*G*) < 1] *and, therefore, would indicate that MS is a “genetic” disease in the sense discussed above in Methods #1E*.}

#### D. Other Constraints on the Non-Observed Parameter Values

Based on theoretical considerations (*see Methods #2C*), we concluded that: (*s*_*a*_ ≥ 1). Indeed, this relationship is confirmed observationally, where the recurrence risk of MS for a proband with a co-twin who is a member of the (*DZ*_*MS*_) subset, is consistently reported to be greater than the recurrence risk of MS for a proband sibling with a co-sibling, who is a member of the (*S*_*MS*_) subset [7,31-37]. Therefore, from the definitions of (*MZ*_*MS*_) and (*IG*_*MS*_) – *see Methods #1C & 1D* – it must be the case that, during our “current” *Time Period*:

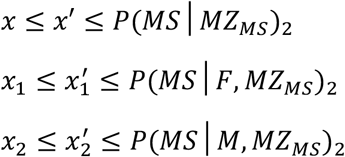

Using the previously described constraints (*above*) on *P*(*MS*|*MZ*_*MS*_)_2_, *P*(*MS*|*F, MZ*_*MS*_)_2_ and *P*(*MS*|*M, MZ*_*MS*_)_2_, therefore, the plausible ranges for (*x*), (*x*_*1*_) & (*x*_*2*_) during the *2*^*nd*^ *Time Period* are:

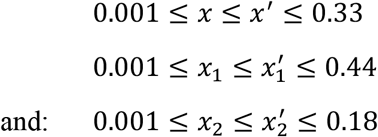

We noted earlier that *P*(*MS*|*G*) varies depending upon *P*(*E*). Also, as noted earlier in *Methods #1D*, the observed *MZ*-twin concordance rates {i.e., *P*(*MS*|*MZ*_*MS*_, *E*_*T*_)} may need to be converted into adjusted rates {i.e., *P*(*MS*|*IG*_*MS*_, *E*_*T*_)} because the observed *MZ-*twin concordance rate will reflect, in part, any increased likelihood that an *MZ-*twin proband will develop MS based on their disproportionately sharing the (*E*_*twn*_) and (*E*_*sib*_) environments with a co-twin who has (or will develop) MS. Because any such increase represents an environmental effect – i.e., it is caused by twins disproportionately sharing the (*E*_*twn*_) and (*E*_*sib*_) environments – the maximum probability of developing MS for susceptible individuals under optimal environmental conditions – i.e., *P*(*MS*|*G, E*) – must be greater (at least marginally, if not more so) than these observed *MZ-*twin concordance rates [3]. Consequently, using the *Table 2* notations, from the definitions of (***c***) and (***d***) – see *Methods #4A; below* – because, currently, (*x*_1_ > *x*_2_), and also because, currently, both *P*(*MS*) and *P*(*F*|*MS*) are increasing, each of the following relationships must hold simultaneously:

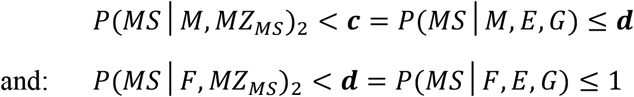

Notably, these relationships *include* the possibility that: ***c*** = ***d*** ≤ 1

Finally, as discussed previously (*see Methods #1D* and *above*), we estimate the impact of the disproportionately shared (*E*_*twn*_) and (*E*_*sib*_) environments for *MZ*-twins as:

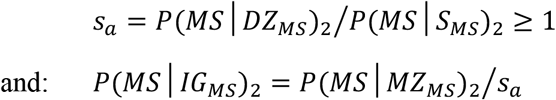

In the Canadian data [7], the life-time probability of developing MS for the proband of a co-*DZ-* twin with MS (5.4%) was found to be greater than that for the proband of a non-twin co-sibling with MS (2.9%). From these observations, the point-estimate for (*s*_*a*_) becomes:

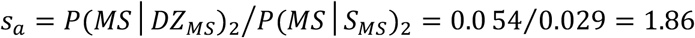

This point estimate is approximately the same for both *men* and *women* [3]. Thus, these observations from Canada suggest that sharing the (*E*_*twn*_) environment with a co-twin who develops MS markedly increases the likelihood of the proband twin developing MS for both *men* and *women* [3]. Nevertheless, it is possible that impact of these disproportionately shared environments may be over- or under-estimated by the Canadian data [7]. In any event, based on theoretical considerations (*see Methods #1D*), if we use the point-estimate from the Canadian data [7] that:

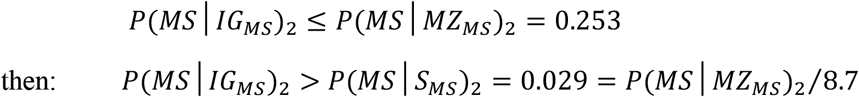

Therefore, for the purpose of both *Models*, we considered the plausible range for (*s*_*a*_) to be:

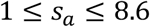

However, because, the point-estimate for (*s*_*a*_) from the Canadian data [7] is generally greater than that reported in other similar studies [31-37], we also considered, separately, the more restrictive circumstance in which:

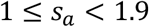

### 3. Cross-sectional Model

For the purposes of this section, because the *Cross-sectional Model* considers only the “current” *Time Period #2*, the environmental designations relating to the conditions of the time – i.e., both the designation of (*E*_*T*_) and the use of the subscript (*2*) – have been eliminated from those parameter definitions that vary with the environmental conditions of the time (*see Methods #1A; above; see also Table 2*). Also, for simplicity of notation, in the development of the *Cross-sectional Model*, we use the definitions from the previous section, which are also provided in *Table 2*.

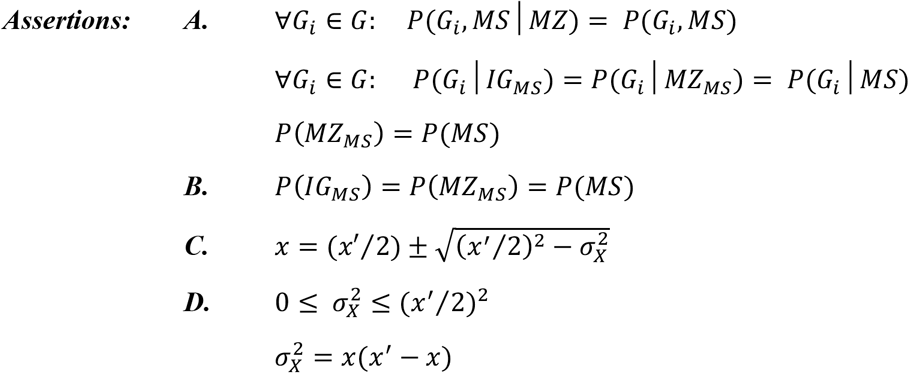

#### Definitions and Assumptions

The subsets (*G*) and (*G*^*c*^) have already been defined *(see Methods #1A*) and as noted there:

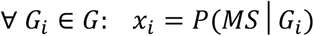

Thus, (*x*_*i*_) represents the MS-penetrance for the *i*^*th*^ individual in the (*G*) subset during <u>any</u> specific *Time Period* and it is unique to the *i*^*th*^ individual, regardless of whether it is identical (in value) to that of another individual.

Also, as discussed in *Methods #1A*, we defined the set (*X*) to include the penetrance value for each of the (*m*) members of the (*G*) subset. Thus: *X* = {*x*_*i*_}. Letting (*x*_*G*_) be a random variable representing any of the {*x*_*i*_} elements within the set (*X*), and because, as noted earlier, all genotypes in a population are unique, it follows that:

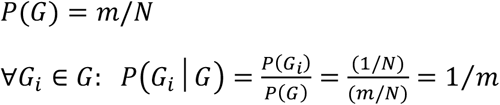

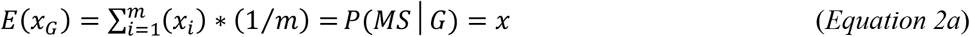

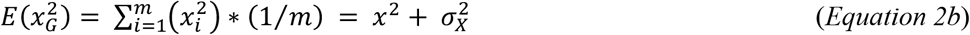

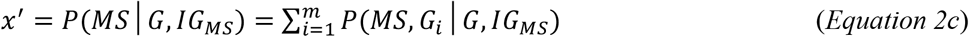

As noted earlier, if 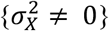, then the subset (*G*) can be partitioned into two mutually exclusive subsets, (*G*1) and (*G*2), suitably defined, such that:

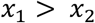

We now specify our two assumptions:

#### Assumption #1

Because *MZ*-twinning is generally thought to be non-hereditary [17-19], we assume that every person (i.e., genotype) in the general population (*Z*) has the same chance, *a priori*, of having an *MZ*-twin (i.e., *MZ*-status is independent of genotype). Thus, we assume that, for any *Time Period*:

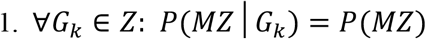

Because this relationship applies to every genotype in (*Z*), therefore, also:

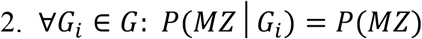

If, rarely, *MZ*-twinning were familial [19], it would still be the case that:

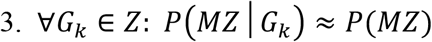

Finally, even if some cases of *MZ*-twinning were thought to be familial [19], the exact relationship (*#2, above*) still follows if those genetic factors, which are associated with *MZ-*twining, are independent of those genetic factors associated with MS. Here we assume that either this or #1 (*above*) pertains.

#### Assumption #2

The penetrance of MS for a proband *MZ*-twin, whose co-twin is of unknown status, is assumed to be the same as if that genotype had occurred without having an *MZ* co-twin (i.e., the penetrance of MS for each genotype is independent of *MZ*-status). This assumption translates to assuming that the impact of experiencing any particular (*E*_*twn*_) and (*E*_*sib*_) environments together with an *MZ* co-twin is the same as the impact of experiencing the same (*E*_*twn*_) and (*E*_*sib*_) environments alone. Alternatively, it translates to the testable hypothesis that the mere fact of having an *MZ* co*-*twin does not alter the (*E*_*twn*_) and (*E*_*sib*_) environments in such a way that MS becomes more or less likely in both twins. Thus, we are here assuming that, for any *Time Period*:

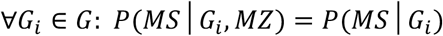

##### Proof of Assertion A

From *Assumption #1*, it follows that:

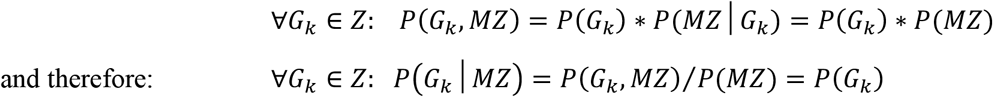

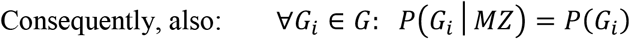

From this conclusion, from the definitions relating to (*MZ*_*MS*_) – *see Methods #1C* – and from *Assumption #2*, it follows that, for any *Time Period*:

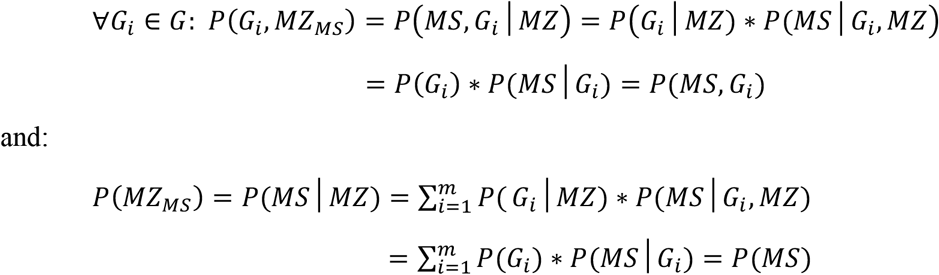

This last conclusion is also evident from the definition of (*MZ*_*MS*_) – see *Methods #1C*. From these two equivalences, and from the definition of (*IG*_*MS*_) – see *Methods #1D* – we conclude that, for any *Time Period*:

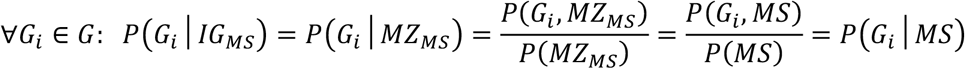

##### Proof of Assertion B

From the definition of (*IG*_*MS*_) – see *Methods #1D* – and from *Assertion A*, it directly follows that, for any *Time Period*:

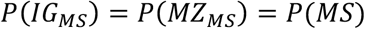

##### Proof of Assertion C

From the definitions of (*G*) & (*IG*_*MS*_) – *see Methods #1A & 1D* – it directly follows that:

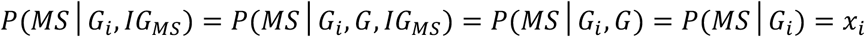

Consequently, during any *Time Period*, the probability *P*(*MS, G*_*i*_|*G, IG*_*MS*_) can be re-expressed as:

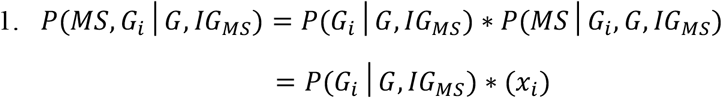

From *Assertion A* and from the definitions of (*G*) & (*IG*_*MS*_) – *see Methods #1A & 1D* – the term *P*(*G*_*i*_|*G, IG*_*MS*_) can be re-expressed as:

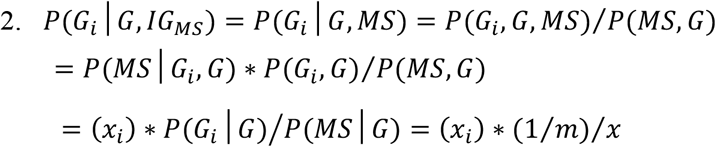

Combining *1 & 2 (above*) yields:

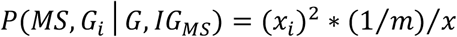

However, from *Equation 2c*, it is the case that:

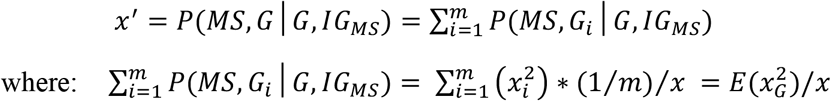

Therefore, from *Equation 2b*, it follows that:

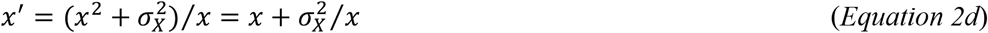

*Equation 2d* can be rearranged to yield a quadratic equation in (*x*) such that:

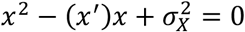

This quadratic can be solved to yield:

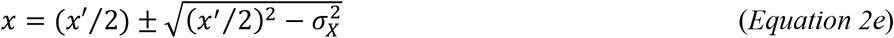

Defining the variance of penetrance values within the subsets of susceptible *women* and *men* as 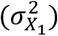 and 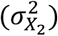, respectively, the same line of argument also leads to the conclusions that:

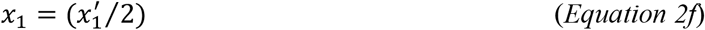

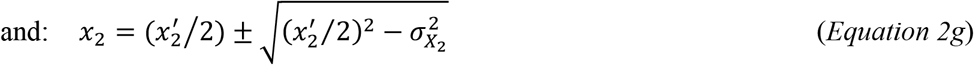

##### Proof of Assertion D

*Equation 2e* has real solutions only for the range of:

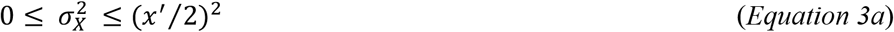

and, also, from *Equations 2f–g*:

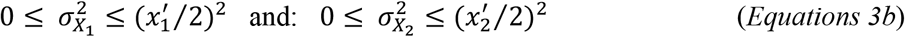

Notably, the maximum variance for any distribution [38] on the closed interval [*a, b*] is:

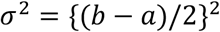

Therefore, irrespective of any *Assumptions* we have made (*see above*), the variances ranges provided in *Equations 3a–b* are the maximum possible variance ranges for *<u>any</u>* distribution on each of the respective closed intervals 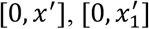 and: 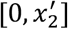.

Also, *Equation 2d* can be re-arranged to yield:

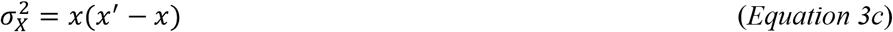

and, from *Equations 3b*:

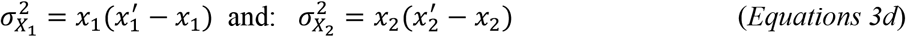

#### Assertion C Solutions

*Equation 2e* has two solutions – the so-called *<u>Upper Solution</u>* and the *<u>Lower Solution</u>*, depending upon the value of the (*±*) sign. The *<u>Upper Solution</u>* represents the gradual transition from a distribution, when 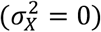, in which everyone has a penetrance of (*x*^′^) to a bimodal distribution, when 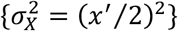, in which half of the (*G*) subset has a penetrance of (*x*^′^) and the other half has a penetrance of zero. Although, under *some* environmental conditions: (∀ *x*_*i*_ ∈ *X*: *x*_*i*_ > 0), as noted previously (*see Methods #1A*), there may be certain environmental conditions, in which, for some individuals in the (*G*) subset:

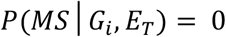

Therefore, the *<u>Upper Solution</u>*, during any particular *Time Period*, is constrained such that:

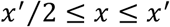

The *<u>Lower Solution</u>* represents the gradual transition from the bimodal distribution described above to increasingly extreme and asymmetric distributions [3]. The *<u>Lower Solution</u>*, however, is further constrained by the requirement of *Equation 2d* that when: 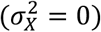 then: (*x* = *x*^′^). Therefore, the *<u>Lower Solution</u>* is constrained such that:

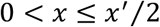

Moreover, regardless of whether the *<u>Upper</u>* or *<u>Lower Solution</u>* pertains, the values that (*x*_1_) and (*x*_2_) can take are further constrained [3]. Thus, using the notation presented *above* and in *Table 2*, for the gender partition, during any *Time Period*:

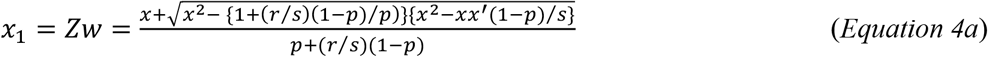

and:

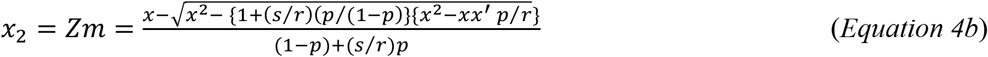

Although the derivation of *Equations 4a & 4b* does not depend directly upon *Assertions A–D* (*above*), nevertheless, one step in the derivation [3] requires the equivalence:

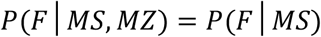

Consequently, *Equations 4a & 4b* make the same two assumptions.

For this analysis we also assume that either the set (*G*) by itself or, considered separately, the sets (*F, G*) and (*M, G*), conform to the *<u>Upper Solution</u>* [3]. In this circumstance, on theoretical grounds, from *Equations 2e–g & 3a–b*, it must be the case that either:

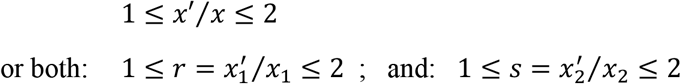

Using a “substitution” analysis, we wrote a computer program, which incorporated the above acceptable ranges (*see Methods #2*) for the parameters {*P*(*G*); *P*(*MS*|*MZ*_*MS*_); *p* = *P*(*F*|*G*); *r; s; P*(*MS*); *P*(*F*|*MS*); and *s*_*a*_}, into the *Summary Equations* (*below*) and determining those combinations (i.e., solutions) that fit within the acceptable ranges for both the observed and non-observed parameters (*see Methods #2*).

*<u>Summary Equations</u>* from *Definitions & Equations 1a–d* ; *2a–e, 3a–b & 4a–b*

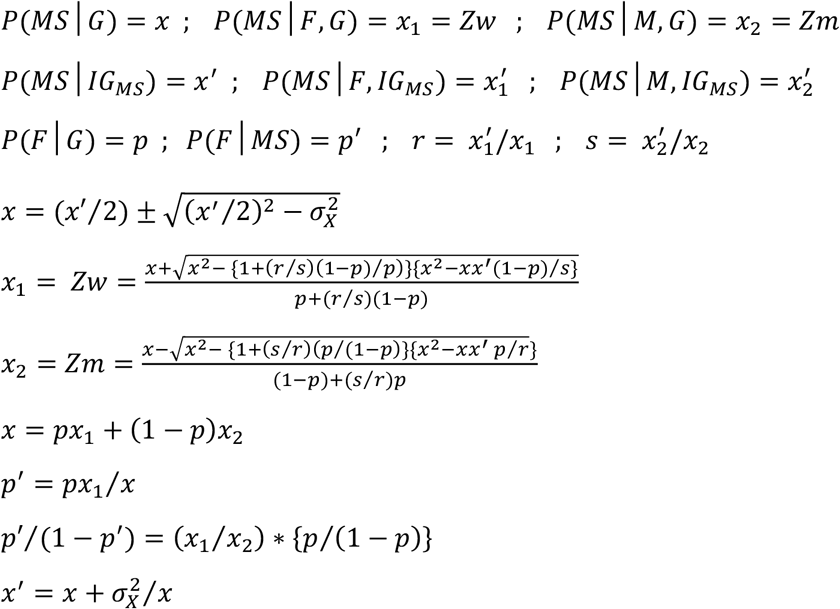

### 4. Longitudinal Model

#### A. General Considerations

Following standard survival analysis methods [39], we define the cumulative survival {*S*(*u*)} and failure {*F*(*u*)} functions where: *F*(*u*) = 1 − *S*(*u*). These functions are defined separately for *men* {*S*_*m*_(*u*) *and F*_*m*_(*u*)} and for *women* {*S*_*w*_(*u*) *and F*_*w*_(*u*)}. In addition, we define the hazard-rate functions for developing MS at different exposure-levels (*u*) in susceptible *men* and *women* {i.e., *h*(*u*) and *k*(*u*), respectively}. These hazard-rate functions for *women* and *men* may or may not be proportional to each other but, if they are proportional, then: *k*(*u*) = *R* * *h*(*u*), where (*R* > 0) represents the hazard proportionality factor. Furthermore, as defined previously (*see Methods #1B*), the term *P*(*E*|*G, E*_*T*_) represents the probability of the event that a member of the (*G*) subset, selected at random, will experience an environmental exposure sufficient to cause MS, given their unique genotype and given the prevailing environmental conditions of the time (*E*_*T*_). We define the exposure (*u*) as the odds of this event during the *Time Period* (*E*_*T*_) such that:

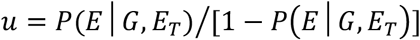

We define *H*(*a*) to be the cumulative hazard function (for *men*) at an exposure-level (*u* = *a*) such that:

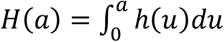

Similarly, we define *K*(*a*) to be the cumulative hazard function (for *women*) at an exposure-level (*u* = *a*) such that:

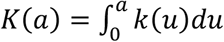

For *men*, following the usual definition of the hazard function [39] that:

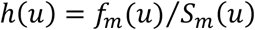

together with the fact that, by definition:

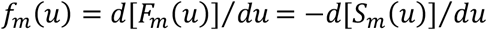

a standard derivation from survival analysis methods [39] demonstrates that, for *men*, because:

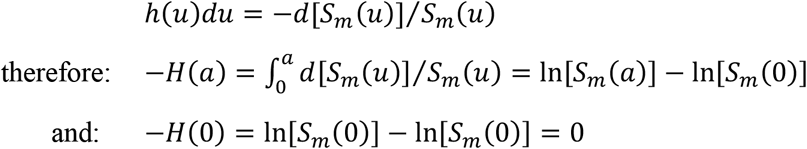

Because we are measuring exposure as the odds that a member of the (*G*) subset receives an environmental exposure sufficient to cause MS, given the environmental conditions at the time, by definition, when: {*P*(*E*|*G, E*_*T*_) = 0}, no member of (*G*) can develop MS {i.e., *S*_*m*_(0) = 1}. In this circumstance: {In[*S*_*m*_(0)] = In(1) = 0}; and therefore:

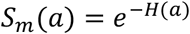

Consequently, the cumulative survival function is exponentially related to the integral of the underlying hazard function – i.e., the cumulative hazard function. Even in the unlikely circumstance that the hazard function is discontinuous at some points, the function will still be integrable in all realistic scenarios. In this circumstance, the failure function for susceptible *men* becomes:

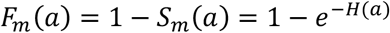

{*NB: In this circumstance, we are using the cumulative hazard function, H*(*a*), *as a measure of exposure for susceptible men, not as a measure of either survival or failure. By contrast, failure, as defined here, is the event that a person develops MS over the course their life-time. The term: Zm* = *P*(*MS, E*|*M, G, E*_*T*_) *represents the probability that, during the Time Period* (*E*_*T*_), *this failure event occurs for a randomly selected individual from the* (*M, G*) *subset of* (*Z*). *Notably, also, the exposure of H*(*a*) *is being used in preference to the, perhaps, more intuitive measure of exposure* (*u* = *a*) *provided above*. *Nevertheless, when* {*P*(*E*|*G, E*_*T*_) = 0}; *both exposure measures are zero – i*.*e*., {*a* = 0} *and* {*H*(*a*) = 0} − *also, as:* {*P*(*E*|*G, E*_*T*_) → 1}; *both exposure measures become infinite: i*.*e*., {*a* → ∞} *and* {*H*(*a*) → ∞} − *and, finally, both exposure measures increase monotonically with increasing P*(*E*|*G, E*_*T*_). *Therefore, the mapping of the* (*u* = *a*) *measure to the H*(*a*) *measure is both one-to-one and onto. Consequently, these two measures of exposure are equivalent and the use of either exposure scale is appropriate. Although the relationship of the H*(*a*) *scale to P*(*E*|*G, E*_*T*_) *is less obvious than it is for the* (*u* = *a*) *scale where: P*(*E*|*G, E*_*T*_) = *a*/(*a* + 1), *the H*(*a*) *scale, nonetheless, has the advantage that the probability of failure is an exponential function of exposure as measured by H*(*a*) *and, thus, it is more mathematically tractable*.}

##### Environmental Exposure Levels during Different Time Periods

If the environmental exposure level for susceptible *men* during the *1*^*st*^ *Time Period* is {*H*(*a*_1_)}, and if, as *above*, we define {*F*_*m*_(*a*) = *Zm*} as the failure probability for *men* (i.e., the probability of the event that a randomly selected susceptible *man* develops MS), during some *Time Period* (*E*_*T*_), and (***c***) as the maximum failure probability for susceptible *men* then:

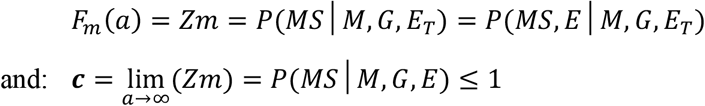

In this case, the failure probability for susceptible *men* during the *1*^*st*^ *Time Period* (*Zm*_1_), can be expressed as:

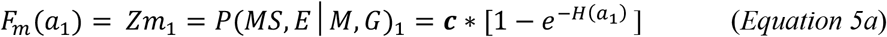

If the exposure level for susceptible *men* during the *2*^*nd*^ *Time Period* is {*H*(*a*_2_)}, then, because (*Zm*) is currently increasing with time, the difference in exposure for *men* between the *1*^*st*^ *and 2*^*nd*^ *Time Periods* can be represented as the difference in the environmental exposure level between these two *Time Periods* (*q*_*m*_) such that:

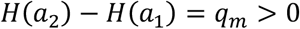

In this circumstance, the failure probability for susceptible *men* during the *2*^*nd*^ *Time Period* (*Zm*_2_), can be expressed as:

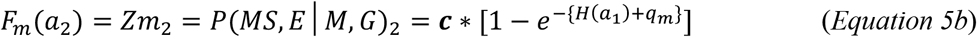

Rearrangement of *Equations 5a & 5b* yields:

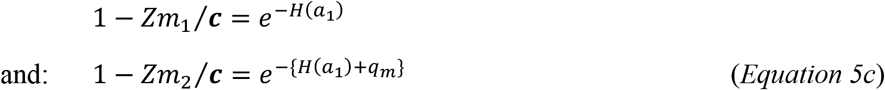

And combining (dividing) these two rearranged *Equations* yields:

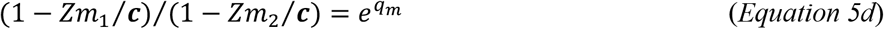

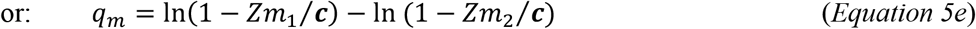

Previously, we assigned the value of these arbitrary units as (*q*_*m*_ = 1) in these *Equations* [3], although such an assignation may be inappropriate. Thus, this unit (whatever it is) still depends upon the actual (but unknown) level of environmental change that has taken place between the two chosen *Time Periods*. From *Equations 5d–e*, this level depends upon the value of (***c***), which can range over the interval of: (1 ≥ ***c*** > *Zm*_2_) – *see Methods #2D*. Because (*Zm*) increases with increasing exposure, the ratio on the *LHS* of *Equation 5d* is always greater than unity and it increases monotonically as (***c***) varies throughout its range. This ratio is at its minimum when: (***c*** = 1) and approaches infinity as: (***c*** → *Zm*_2_). Therefore, we can define 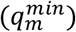 as the *“minimum”* possible exposure level change for *men* between these two *Time Periods*. In this case, from *Equations 5d & 5e*, this *minimum* exposure level change will occur under circumstances where:

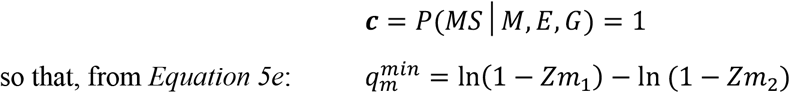

However, this *minimum* exposure level change 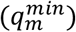 may not accurately characterize the actual (but unknown) level of environmental change, which has taken place for *men* between the two *Time Periods*. Therefore, we will refer to (*q*_*m*_) as the *“actual”* exposure-level change, which may be different from this *minimum* exposure-level change such that:

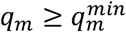

In an analogous manner, we also define {*F*_*w*_(*a*) = *Zw*} as the failure probability for susceptible *women* during any *Time Period* and (***d***) as the ultimate failure probability for susceptible *women* such that:

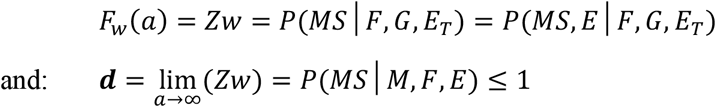

In this case, because (*Zw*) is also currently increasing with time, the *Equations* for the failure probability in susceptible *women* during the *1*^*st*^*& 2*^*nd*^ *Time Periods*, (*Zw*_1_ *and Zw*_2_), become:

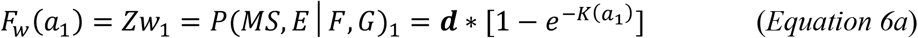

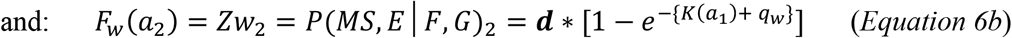

where {*K*(*a*_1_)} represents the exposure level in *women* at the *1*^*st*^ *Time Period* and (*q*_*w*_) represents the *“actual”* level of environmental change for *women*, which has taken place between the two *Time Periods* such that:

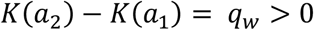

In a manner directly analogous to that presented *above* for the development of *Equation 5e*, it is also the case that:

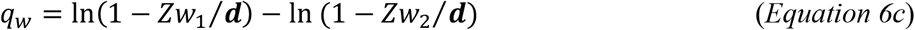

Thus, as *above* for susceptible *men*, the *“minimum”* value 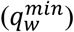 for the exposure level change in susceptible *women* will occur under those circumstances for which: (***d*** = 1), so that:

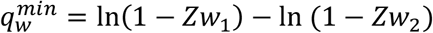

##### Relationship of Failure to True Survival

However, unlike true survival (where everyone dies given sufficient time), the probability of developing MS, either for the subset of susceptible *women* {*Zw* = *P*(*MS, E*|*F, G*)} or for the subset of susceptible *men* {*Zm* = *P*(*MS, E*|*M, G*)}, may not approach 100% as the probability of exposure {*P*(*E*|*G, E*_*T*_)} approaches unity. Moreover, the limiting value for the probability of developing MS in susceptible *men* (***c***) need not be the same as that in susceptible *women* (***d***). Also, even though the values of the (***c***) and (***d***) parameters are unknown, they are, nonetheless, constants for any disease process, which requires environmental factors as an essential component of disease pathogenesis, and they are independent of whether the hazards are proportional. Finally, the threshold environmental exposure (at which MS becomes possible) must occur at: *P*(*E*|*G, E*_*T*_) = 0; for one (or both) of these two subsets, provided that this exposure level is possible [3]. If the hazards are proportional, a difference in threshold (*λ*) can be defined as the difference between the threshold in *women* (*λ*_*w*_) and the threshold in *men* (*λ*_*m*_) – i.e., (*λ* = *λ*_*w*_ − *λ*_*m*_). Thus, if the threshold in *women* is greater than the threshold in *men*, (*λ*) will be positive and (*λ*_*m*_ = 0); if the threshold in *men* is greater than the threshold in *women*, (*λ*) will be negative and (*λ*_*w*_ = 0).

Also, in true survival, both the clock and the risk of death begin immediately at time-zero and continue indefinitely into the future, so that the cumulative probability of death always increases with time. By contrast, here, it may be that the prevailing environmental conditions during some *Time Period* (*E*_*T*_) are such that: *P*(*E*|*G, E*_*T*_) = 0 ; even for quite an extended period (e.g., centuries or millennia). In addition, unlike the cumulative probability of death, here, exposure can vary in any direction with time depending upon the specific environmental conditions during (*E*_*T*_). Therefore, although the cumulative probability of failure (i.e., developing MS) increases monotonically with increasing exposure, it can increase, decrease, or stay constant with time.

##### Relationship of the (F:M) Sex Ratio to Exposure

Finally, regardless of (*λ*), and regardless of any proportionality, during any *Time Period*, the failure probability for susceptible *women* (*Zw*) can be expressed as:

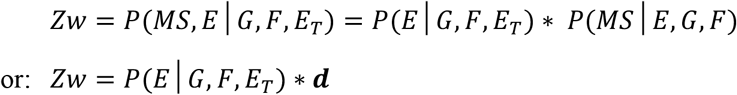

and, similarly, the failure probability for susceptible *men* (*Zm*) can be expressed as:

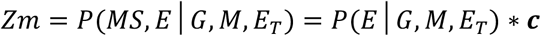

Dividing the *2*^*nd*^ of these two *Equations* by the *1*^*st*^, during any *Time Period*, yields:

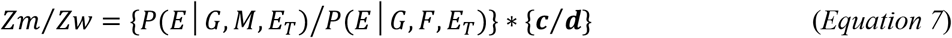

Consequently, any observed disparity between (*Zw*) and (*Zm*), during any *Time Period*, must be due to a difference between *men* and *women* in the likelihood of their experiencing a sufficient environmental exposure, to a difference between (***c***) and (***d***), or to a difference in both. It is not due to any difference between *women* and *men* in their responsiveness to a sufficient environmental input – at least insofar as this difference is due to something other than the limiting values of (***c***) and (***d***). Under conditions of: (***c*** = ***d*** ≤ 1), therefore, any difference between (*Zw*) and (*Zm*) must be due exclusively to a difference in exposure to sufficient environments between the two genders. Also, regardless of whether the hazards are proportional, and because proportion of *women* among susceptible individuals (*p*) is a constant (*see Table 2*), therefore, for any solution, the ratio (*Zw*/*Zm*), during any *Time Period* (*E*_*T*_), will be proportional to the observed (*F:M*) *sex ratio* during that period (*see Equation 1b*).

##### The Response Curves to Increasing Exposure

Notably, also, because the response curves for both *men* and *women* are exponential, any two points of observation on these curves will define the entire curve (e.g., the values of *Zw* and *Zm* during *Time Period #1 and Time Period #2 – see Equations 5a & 5b* and *6a & 6b*). Moreover, if these two curves can be plotted on the same *x-axis* (i.e., if *men* and *women* are responding to the same environmental events), the hazards are always proportional and the values of (*R* = *q*_*w*_/*q*_*m*_) and (*λ*) are determined by a combination of the observed values for (*Zw*_2_), (*Zm*_2_), and the (*F:M*) *sex ratio* change and the fixed (but unknown) values of (***c***), (***d***), and (*C*) – *see Equations 5e and 6c (above); see also Equation 10c & Summary Equations (below)*.

#### B. Non-proportional Hazard

If the hazard functions for MS in *men* and *women* are not proportional, it is always possible that the “*actual*’ exposure level changes for *men* and *women* are each at their *“minimum”* values – i.e., 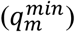 and 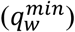 – such that: (***c*** = ***d*** = 1).

In this circumstance, the various observed and non-observed epidemiological parameter values still limit possible solutions. However, although (***c***) and (***d***) will still be constants, respectively, for *men* and *women*, no information can be learned about them or about their relationship to each other from changes in the (*F:M*) *sex ratio* and *P*(*MS*) over time. The observed changes in these parameter values over time could all simply be due to the different environmental circumstances of different times and different places. In this case, also, although *men* and *women* will still each have environmental thresholds, the parameter (*λ*) – which relates these thresholds to each other – is meaningless, and there is no hazard proportionality factor (*R*).

Nevertheless, even with non-proportional hazard, the ratio (*Zw*/*Zm*), during any *Time Period*, must still be proportional to the observed (*F:M*) *sex ratio* during that *Time Period* (*see Equation 1b*) and, if: ***c*** = ***d*** ≤ 1, then any observed disparity between (*Zw*) and (*Zm*), must be due entirely to a difference between *women* and *men* in the likelihood of their experiencing a sufficient environmental exposure (*E*) during that *Time Period* (*see Equation 7; above*).

#### C. Proportional Hazard

By contrast, if the hazards for *women* and *men* are proportional with the proportionality factor (*R*), the situation is altered. First, because (*R* > 0), those changes, which take place for the subsets *P*(*F, MS*) and *P*(*M, MS*) over time, must have the same directionality. Indeed, this circumstance is in accordance with our epidemiological observations where, over the past several decades, the prevalence of MS has been noted to be increasing for both *women* and *men* [6,22-30]. Second, including a possible difference in threshold between the genders, the proportionate hazard *Model* can be represented by those circumstance for which:

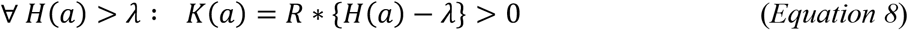

In this case, *Equations 6a & 6b*, which represent the failure probability in susceptible *women* given the exposure during the *1*^*st*^*& 2*^*nd*^ *Time Periods*, can be re-written as:

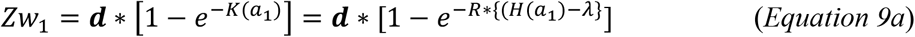

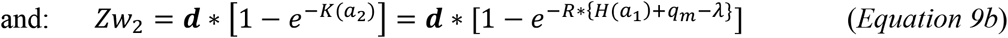

Rearranging *Equations 5a & 9a* for *<u>any</u> Time Period* yields:

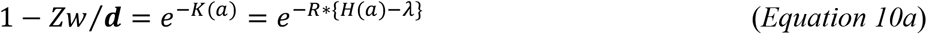

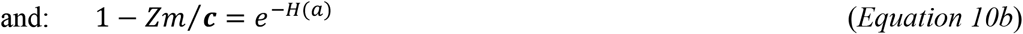

Division of *Equation 10a* by *10b*, and rearrangement, during any *Time Period*, yields:

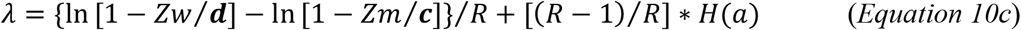

Consequently, for those circumstances in which (*R* = 1), *Equation 10c* becomes:

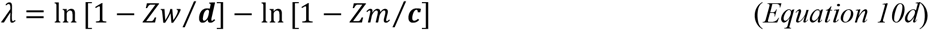

For any specific exposure level {*H*(*a*) > *λ*}, the quantities (*Zw*) and (*Zm*) are unknown.

However, considering any disease for which a proportionate hazard *Model* is appropriate, the parameters (***c, d***, *R*, & *λ*) are fixed (but unknown) constants, so that, from *Equations 10a & 10b*, the values of (*Zm*) and (*Zw*) are also fixed at any specific exposure level {*H*(*a*)}.

##### Defining an “Apparent” Proportionality Factor

We can also define a so-called “*apparent*” value of the hazard proportionality factor (*R*^*app*^) such that: 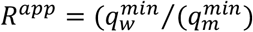. This “*apparent*” value incorporates, potentially, two fundamentally different processes. First, it may capture the increased level of “sufficient” exposure experienced by one group compared to the other. Indeed, from *Equation 7*, this is the only interpretation possible for circumstances in which: (***c*** = ***d*** ≤ 1). Second, however, if we admit the possibility that: (***c*** < ***d*** ≤ 1), then some of (*R*^*app*^) will be accounted for by the difference of (***c***) from unity. For example, using the *Summary Equations* (*below*), and using the proportionate hazard *Model*, (*q*_*m*_) – the *“actual”* exposure level change in *men* – when (***d*** = 1), has the limits:

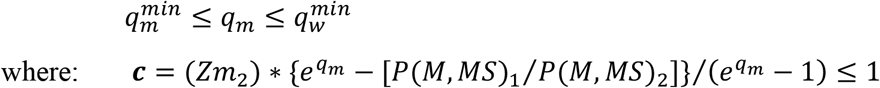

From this, we can define the *“actual”* hazard proportionality factor (*R*), at (***d*** = 1), such that:

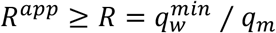

In this manner, if 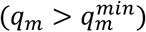, a portion of the “*apparent*” value (*R*^*app*^) will be accounted for by a reduction in value of (***c***) from unity, if such a reduction is possible. Moreover, if such a reduction is possible for susceptible *men*, then, clearly, it is also possible that the value of (***d***) is also reduced from unity in susceptible *women*, in which case: (***c*** < ***d*** < 1), where the *“actual”* exposure level in *women* (*q*_*w*_) would be greater than its minimum value 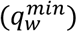 such that:

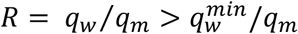

Consequently, in each of these circumstances, the *“actual”* value of (*R*) may be different from its *“apparent”* value (*R*^*app*^) although, from *Equations 7, 9a–b & 10a–b*, under the conditions of: (***c*** = ***d*** ≤ 1) then: (*R* = *R*^*app*^).

{*NB: Considering Equations 5a & 9a for any Time Period (above), the* (***c***) *and* (***d***) *constants, although defined differently (see above), are each the equivalent of a y-axis “scaling factor” for their respective exponential curves, considered separately. However, when* (***c*** = ***d***), *this “scaling factor” is the same for both men and women and, therefore, under those conditions where:* (***c*** = ***d*** ≤ 1), *all response curves for both men and women, which have the same* (*R*) *and* (*λ*), *will differ from each other only in that the y-axis scale is different. For example, suppose that we define a constant* (*α*) *such that:* ***c*** = ***d*** = *α* < 1. *In this case, the response curves for men and women, depicted after changing the y-scale by multiplying both sides of Equations 5a & 9a (for any Time Period) by the constant* (1/*α*), *are identical to those curves having the same* (*R*) *and* (*λ*), *but depicted for:* (***c*** = ***d*** = 1).

*This does not imply that these response curves are “actually” identical. Rather, the fact that the curves for those conditions under which:* (***c*** = ***d*** = *α* < 1) *can be scaled to be identical to those depicted for:* (***c*** = ***d*** = 1), *only indicates that the relationship <u>between</u> the response curve for men and that of women is the same regardless of the value of* (*α*) *– i*.*e*., *any changes in the* (*Zw*/*Zm*) *ratio with increasing exposure or, equivalently, any changes of the F:M sex ratio (see below, see also Equation 1b), will be the same for any value of* (*α*). *When* (***c*** < ***d***), *this kind of transformation is not possible*.}

##### *Implication that the* (*R*) *Value has for the Value of* (*λ*)

We define the ratios (*C*_*F*_ & *C*_*M*_) – *see Table 2* – and note that, because both *P*(*MS*) and the (*F:M*) *sex ratio* are both increasing with time [6,22-30], therefore:

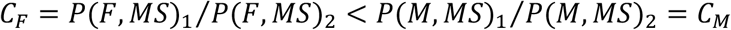

Notably, also, from *Equation 1b*, during any *Time Period*:

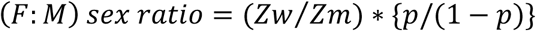

where (*p*) is independent of the environmental conditions of any *Time Period*. Therefore, for all solutions, the ratio (*Zw*/*Zm*) will mirror (*F:M*) *sex ratio* (i.e., changes in both ratios will have the same directionality).

##### *For Conditions where: (R* = 1)

Under those circumstances for which: (*R* = 1), it must be the case that:

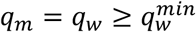

When: (*λ* = 0), from *Equation 10d*:

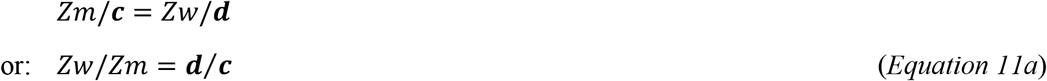

Therefore, in this circumstance, the *F:M sex ratio* will remain constant, regardless of the exposure level.

However, using the previously derived *Summary Equations* [3] presented at the end of this section:

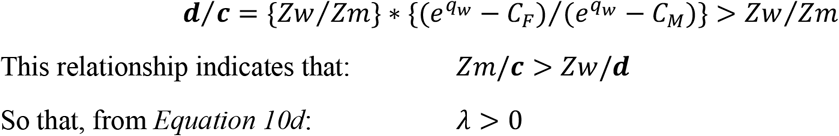

Thus, if (*R* = 1) and the *F:M sex ratio* is increasing, it must be the case that the threshold in susceptible *women* is greater than that in susceptible *men*.

##### *For Conditions where: (λ* ≤ 0) & (*R* > 1)

Because (*H*(*a*) ≥ 0), therefore, from *Equation 10c*, under these conditions, for any *Time Period*:

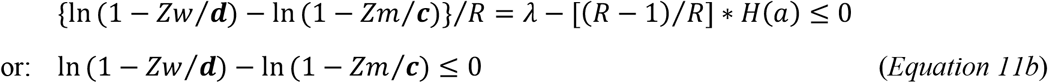

In turn, *Equation 11b*, in this circumstance, requires that:

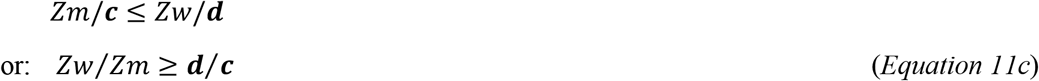

Moreover, regardless of the value of (*R*), from the definitions of (*E*), (***c***), and (***d***), and from *Equation 7* (*above*):

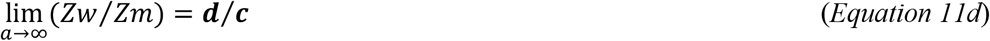

Therefore, because both (*Zw*) and (*Zm*) increase monotonically with increasing exposure (*see Methods #4A; above*), and because (*<u>R</u>* > 1), and because (*H*(*a*) ≥ 0), the condition that:

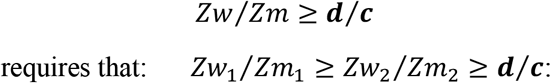

so that, in this circumstance, the ratio (*Zw*/*Zm*) decreases or remains constant with increasing exposure. Because the (*Zw*/*Zm*) ratio mirrors the *F:M sex ratio* (*see above*), therefore, under these conditions, the *F:M sex ratio* will also be decreasing or remaining constant (e.g., *Figures 1A–B*) – a conclusion, which is contrary to all the available evidence [6,22-30]. Consequently, the conditions of: (*R* > 1) & (*λ* ≤ 0) are not plausible, given the Canadian data [6].

Combining these two conclusions, therefore, it is clear that:

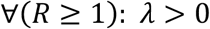

Thus, based solely on the observations of an increasing *P*(*MS*) and an increasing (*F:M*) *sex ratio* with time [6,22-30] – a circumstance which is true considering the “current” *Time Period #2* together with any of the reported previous 5-year epochs as *Time Period #1* [6] – we can conclude, based on purely theoretical grounds, that, if the hazards are proportional, and if: (*R* ≥ 1), susceptible *women* must have a higher threshold than susceptible *men*.

##### *For Conditions where*: (*λ* ≥ 0) & (*R* ≤ 1)

If: (*λ* ≥ 0) & (*R* ≤ 1) & (***c*** = ***d*** ≤ 1) ; then *men* would have as great (or a greater) failure probability than *women* (i.e., *Zm* ≥ *Zw*) at every exposure level (e.g., *Figure 1C*). However, as noted in *Methods #2C* (*abov*e), currently:

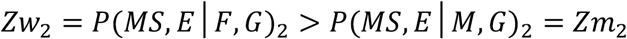

so that the posited conditions are not possible. Nevertheless, under the conditions of (*λ* ≥ 0) & (*R* ≤ 1), it is still possible that: (***c*** < ***d*** ≤ 1) – e.g., *Figure 1D*.

##### *For Conditions where: (λ* < 0) & (*R* ≤ 1)

In this circumstance, *Equation 11b* still holds so that; if: (***c*** = ***d*** ≤ 1), then: (*Zm* > *Zw*) so that, again, the posited conditions are not possible although it is still possible that: (***c*** < ***d*** ≤ 1).

Thus, based on these latter two conditions, it is clear that:

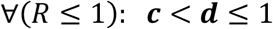

#### D. *Strictly Proportional Hazard*: (*λ* = 0)

If the hazards in *men* and *women* are “strictly” proportional to each other, then it must be the case that: (*λ* = 0). Therefore, when: (|*λ*| > 0), as it must be when (*R* ≥ 1), the hazards cannot be “strictly” proportional. Indeed, for those circumstances in which (*R* ≥ 1) and (*λ* = 0), the observed (*F:M*) *sex ratio*, as discussed *above*, either decreases or remains constant with increasing exposure (*see Method #4 Equations 11a–c*), regardless of the parameter values for (***c***) and (***d***) – e.g., *Figures 1A & 1B*. Consequently, the only “strictly” proportional circumstances, which are possible, are those in which *men* have a greater hazard than *women* – i.e., (*R* < 1). Moreover, if *men* have a greater hazard than *women*, then, as noted *above*, the conditions of: (***c*** = ***d*** ≤ 1) & (*λ* = 0) are also excluded.

Thus, the only possible “strictly” proportional circumstances are those, in which both (*R* < 1) and (***c*** < ***d*** ≤ 1) – e.g., *Figure 1D*.

{*NB: In these and subsequent Figures, all response curves exemplifying the conditions in which* (***c*** = ***d*** ≤ 1), *are depicted for the condition* (***c*** = ***d*** = 1). *Nevertheless, for all those conditions where* (***c*** = ***d*** < 1), *the response curves differ from the curves depicted in the Figures only in so far as the y-axis has a different scale. Therefore, the response curves, depicted at:* (***c*** = ***d*** = 1), *are representative of all curves for which* (***c*** = ***d***) *– see Methods #3C*.}

#### E. *Intermediate Proportional Hazard: (λ* < 0)

We can also consider another possible *Model*, which is intermediate between the “strictly” proportional and non-proportional hazard *Models* discussed *above*. In this intermediate *Model*, the hazards are still held to be proportional although the onset of the response curves are offset from each other by an amount (*λ* ≠ 0). As noted earlier: ∀ (*R* ≥ 1): *λ* > 0. Therefore, for those circumstances in which (*λ* < 0), the hazard in *men* must be greater than the hazard in *women*. Otherwise, the (*F:M*) *sex ratio* will decrease with increasing exposure, which is contrary to the evidence [6] – e.g., *Figure 2A*. Moreover, under those conditions, for which (***c*** = ***d*** ≤ 1) & (*R* < 1) & (*λ* < 0), the (*F:M*) *sex ratio* will initially decrease with increasing exposure to a level below {*p*/(1 − *p*)} and then, this ratio will steadily increase back to this ratio – e.g., *Figure 2B*. However, such a circumstance requires that (*Zm* > *Zw*) throughout the entire response curve until an (*F:M*) *sex ratio* of: {*p*/(1 − *p*)} is reached. This is also contrary to the evidence where currently (*Zm*_2_ < *Zw*_2_) and, thus, where: {(*F*: *M*) > *p*/(1 − *p*)} – *see Equation 1b; Methods #2C*. Thus, the only circumstances, in which (*λ* < 0) is possible, are those where: (***c*** < ***d***) – e.g., *Figure 2D* – but, even then, this is only possible in limited circumstances – e.g., *Figures 2C–D*.

**Figure 2.**
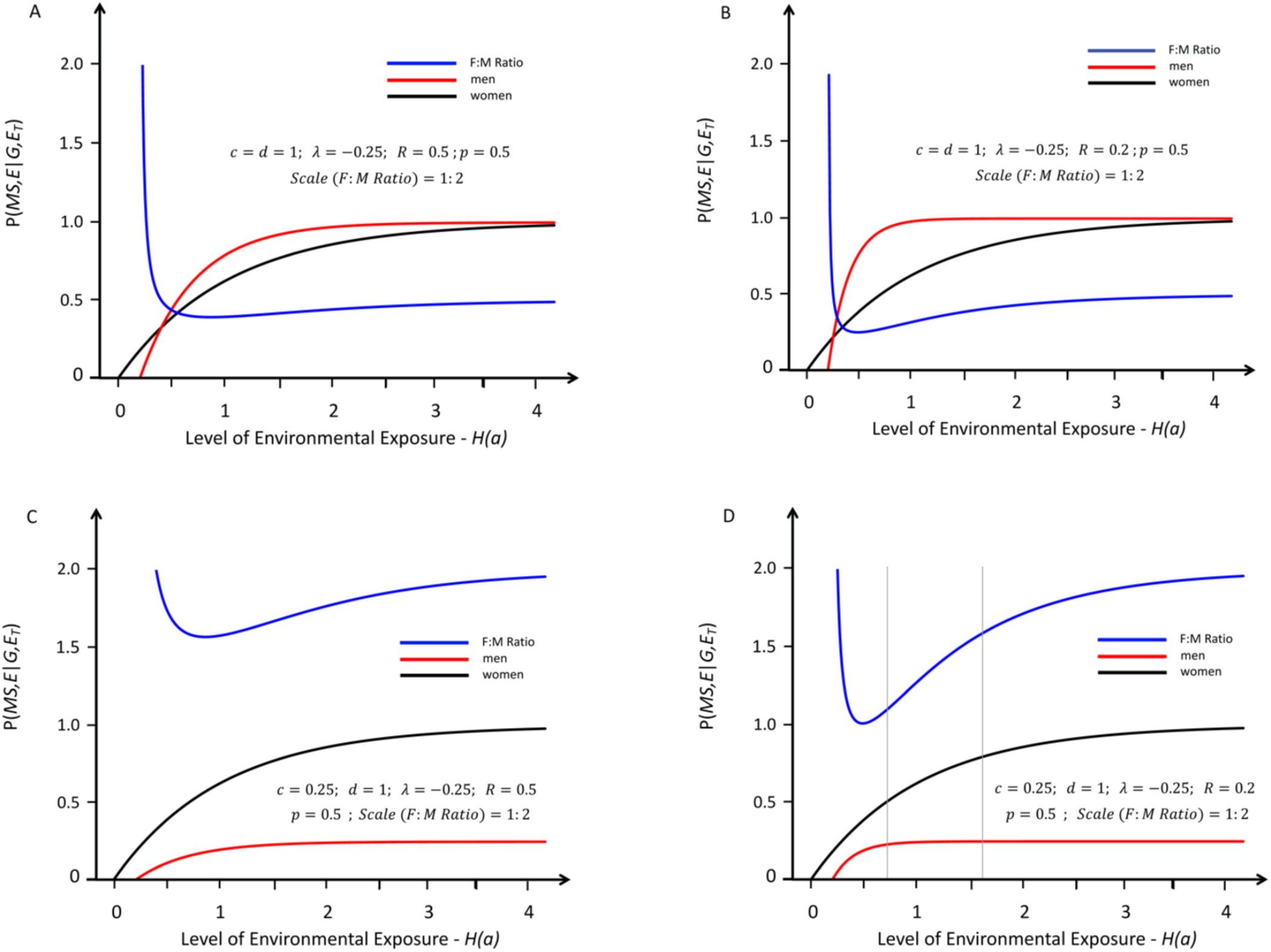
Response curves for the likelihood of developing MS in genetically susceptible *women* (black lines) and *men* (red lines) with an increasing probability of a “sufficient” environmental exposure – *see Methods #1B*. Like *Figure 1*, the curves depicted are also proportional although here the environmental threshold is greater for *men* than for *women* – i.e., under conditions in which: (*λ* < 0) – *see Text*. The blue lines represent the change in the (*F:M*) *sex ratio* (plotted at various scales) with increasing exposure. The thin grey vertical lines represent the portion of the response curve that covers the change in the (*F:M*) *sex ratio* from 2.2 to 3.2 (i.e., the actual change observed in Canada [6] between *Time Periods #1 & #2*). The grey lines are omitted under circumstances either where these observed (*F:M*) *sex ratios* are not possible or where both an increasing (*F:M*) *sex ratio* and (*Zw* > *Zm*) are not possible. Response curves *A* reflects conditions in which (***c*** = ***d*** = 1) & (*R* > 1); Response curves *B* reflects conditions in which (***c*** = ***d*** = 1) & (*R* < 1); curve *C* reflects conditions in which (***c*** < ***d*** = 1) and (*R* < 1) and curve *D* reflects those conditions in which (***c*** < ***d*** = 1) and (*R* < 0.5). To account for the observed increase in the (*F:M*) *sex ratio*, curve *D* (compared to curve *C*) requires a small enough value of (*R*) so that the (*F:M*) *sex ratio* curve dips below 2.2 and, also, a small enough value of (***c***) so that the curve rises above 3.2. For all points for curve *A* (*Zw*/*Zm* > ***d***/***c***). Curves *B & C* never even approach the (*F:M*) *sex ratio* of 2.2. By contrast, for curve *D*, an appropriate increase in the (*F:M*) *sex ratio* can be observed.

#### F. *Intermediate Proportional Hazard: (λ* > 0)

By contrast, when (*λ* > 0), although the hazard is still proportional, there are no absolute constraints on the hazard in *men* relative to that in *women*. Thus, the conditions of: (*R* < 1) & (*λ* ≥ 0) – e.g., *Figure 1D* – and: (*R* ≥ 1) & (*λ* > 0) – e.g., *Figure 3C* – can each lead to very similar conclusions. For this discussion, we define (*G*_*ia*_) to be the susceptibility genotype of the *i*^*th*^ susceptible individual that includes all (and only) those genetic factors, related to MS susceptibility, which are located on autosomal chromosomes. Also, the occurrence of (*G*_*ia*_) represents the event that an autosomal genotype, randomly selected from all such genotypes within (*Z*), is a member of the (*G*_*ia*_) subset. We define the family {*G*_*a*_} to include all the subsets (*G*_*ia*_) within the (*G*) subset. In a similar manner, the occurrence of {*G*_*a*_} represents the event that an autosomal genotype, randomly selected from all such genotypes within (*Z*), is a member of the {*G*_*a*_} family. The “susceptibility” genotype (*G*_*is*_), defined previously (*Methods #1A*), includes all (and only) those genetic factors, which are related to MS susceptibility (located on any chromosome) but does not include the entire genotype of the *i*^*th*^ individual. The event (*G*_*is*_) and the family {*G*_*s*_} have also been defined previously (*see Methods #1A*). Naturally, *women* and *men* may be members of the same (*G*_*ia*_) subset, but not be members of the same (*G*_*is*_) subset, either if there are factors on the *X-* chromosome related to susceptibility that differ between some susceptible *men* and *women*, or if there are factors on the *Y-*chromosome in some *men*, which are related to susceptibility.

**Figure 3.**
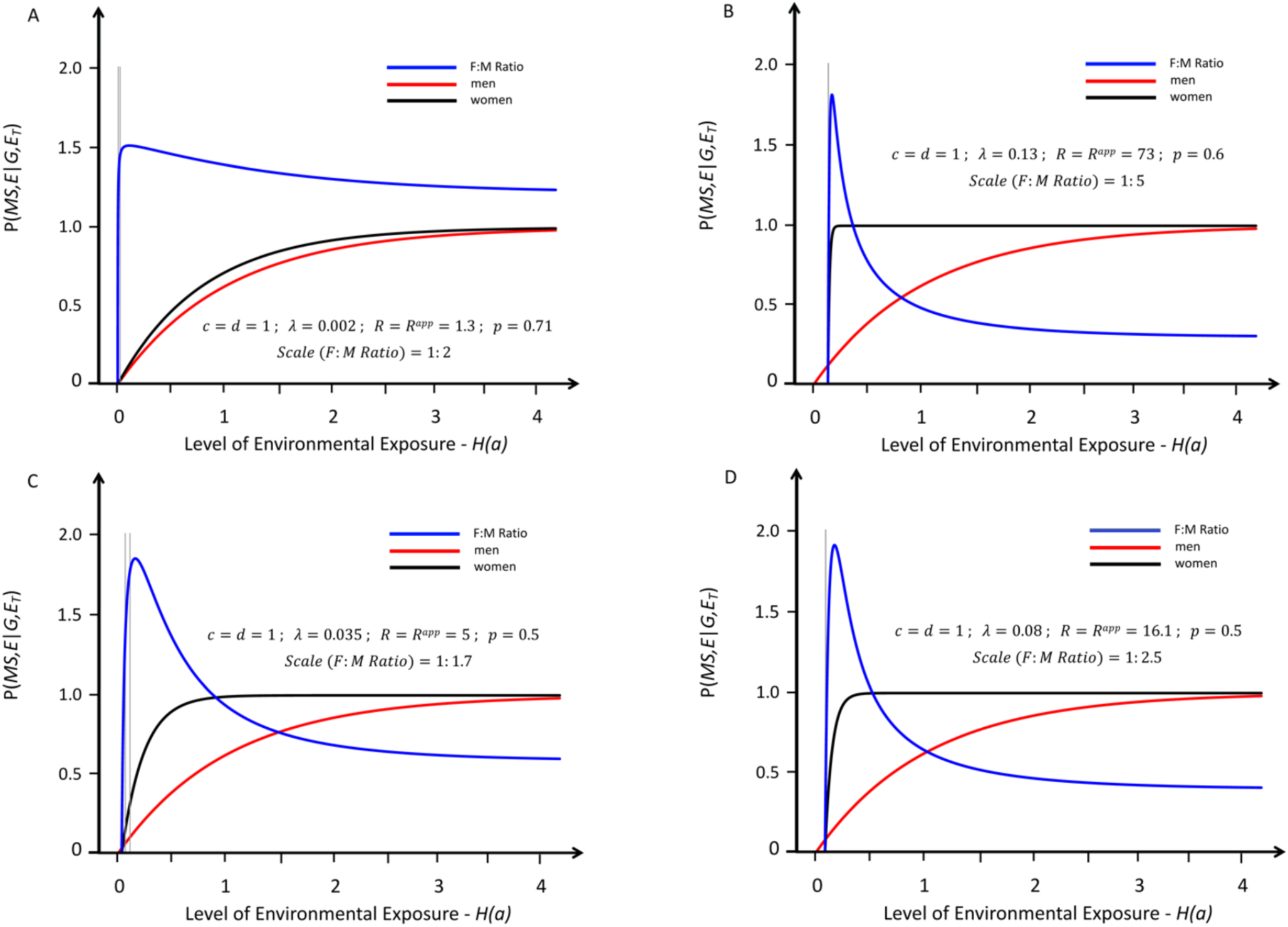
Response curves for the likelihood of developing MS in genetically susceptible *women* (black lines) and *men* (red lines) with an increasing probability of a “sufficient” environmental exposure – *see Methods #1B*. Like *Figure 1*, the curves depicted are also proportional (*R* = *R*^*app*^), but, for these, the environmental threshold in *women* is greater than that it is in *men* – i.e., these are conditions in which: (*λ* > 0). Also, all these response curves represent actual solutions and reflect conditions in which (***c*** = ***d*** = 1) and, as discussed in *Methods #4C*, are representative of all conditions in which ***c*** = ***d*** < 1). Moreover, with increasing values from (*R*^/00^ ≥ 1.3), which is the minimum value of (*R*^*app*^) for any solution – which is depicted in *Figure A*. The blue lines represent the change in the (*F:M*) *sex ratio* (plotted at various scales) with increasing exposure. The thin grey vertical lines represent the portion of the response curve (for the depicted solution), which represents the actual change in the (*F:M*) *sex ratio* that occurred between *Time Periods #1 & #2*). To account for the observed increase in the (*F:M*) *sex ratio*, these curves require the Canadian observations [6] to have been made over a very small portion the response curve – i.e., for most of these response curve, the (*F:M*) *sex ratio* is decreasing. Also, for each of these response curves, including the maximum difference in the environmental threshold (i.e., *λ* ≤ 0.13) under conditions of (***c*** = ***d*** = 1), which is depicted in *Figure B*, the ascending portion of the curve (which reflects and increasing *F:M sex ratio*) is very steep – a circumstance indicating that the portion of the response curve available for fitting the Canadian data [6] is quite narrow.

##### Considerations of Exposure “Intensity”

To evaluate the implications of circumstances in which (*λ* > 0), we also need to develop a notion of what we will call the “*intensity*” of exposure. For example, suppose that *<u>every</u>* set of sufficient exposures for *<u>every</u>* susceptible individual (of any *i-type*) includes both a deficiency of vitamin D and an Epstein Barr Viral (*EBV*) infection [3], each event needing to occur during some (but not necessarily the same) critical period during a person’s life [8]. Moreover, suppose further that, to cause MS, the vitamin D deficiency needs to be more severe or longer lasting in *women*, or that the critical age-window for the *EBV* infection is narrower in *women* than *men*, or both. In each of these circumstances, we can describe susceptible *women* as requiring a greater “*intensity*” of exposure than susceptible *men* and any threshold difference (*λ*) will reflect this increased “*intensity”* required by susceptible *women*.

This framing, however, does not imply that susceptible *wome*n require a more “*intense*” exposure to every environmental factor within a sufficient set. Nor does it imply that every observed change in environmental exposure will demonstrate this altered threshold. Thus, for example, suppose that (in the *above* example) susceptible *men* and *women* require the same “*intensity*” of *EBV* exposure but that *women* require a more “*intense*” vitamin D deficiency. In this circumstance, if the environmental change that occurs is due, in whole or in in part, to a change in population vitamin D levels, then the threshold difference will be apparent. By contrast, if the environmental change that occurs only involves only a change in *EBV* exposure, no threshold difference will be identified although, also, no change in the *F:M sex ratio* will be found (*see Methods #4C; Equations 11a-c*). Nevertheless, for those circumstances in which the hazards are proportional and: (*λ* > 0), we can be confident that the environmental change, which took place between the two *Time Periods*, include changes in a factor (or factors) for which *women* require a more *“intense*” exposure.

Also, if *men* require a less “*intense*” exposure to develop MS than *women*, presumably, in such a circumstance, both genders can still develop MS in response to the more “*intense*” exposures required by *women* (*see Methods #1B* for a discussion regarding environmental factors representing a range of exposures). In this case, we also need to expand our definition of the exposure family {*E*_*i*_}. Thus, because (*G*_*ia*_) genotypes are purely autosomal, we expect that:

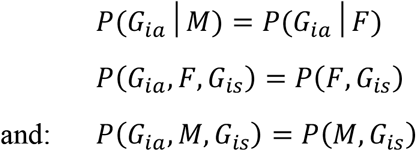

However, as noted *above*, these equivalences do not imply either that:

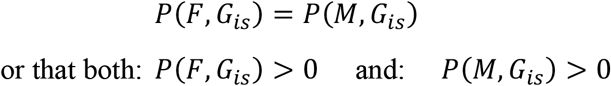

In this conceptualization, the environmental factors that comprise each set of sufficient exposures within the {*E*_*i*_} family (for *i-type* individuals) are the same regardless of whether the *i-type* individual happens to be a *man* or a *woman*. The difference, however, is that *women*, compared to *men*, require a more “*intense*” exposure to one or more of the factors that comprise these sets. We designate this subset of more “*intense*” exposures, within {*E*_*i*_}, as {*E*_*iw*_}. The subset of (*E*), which includes only the more “*intense*” environmental exposures required by *women* will be designated as (*E*_*w*_). By this definition, therefore: (*E*_*w*_) ⊂ (*E*) ; so that

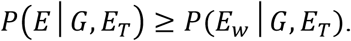

Following the arguments used in the development of *Equation 7* (*Methods #4A, above*) and, if *women* require a more *“intense”* exposure than *men* {i.e., for (*λ* > 0) *as envisioned above*}, then, during any *Time Period*:

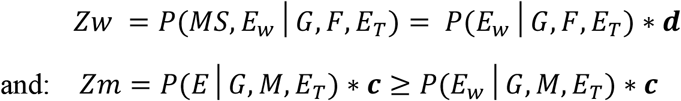

And, again, we can divide the *1*^*st*^ of these two *Equations* by the *2*^*nd*^, to yield:

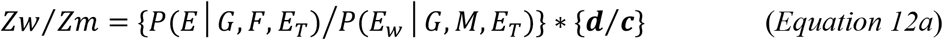

Consequently, for any circumstance in which: ***c*** = ***d*** ≤ 1:

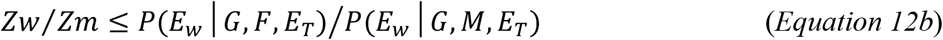

Therefore, by assuming that: (***c*** = ***d*** ≤ 1), we are also assuming that any difference in disease expression between susceptible *men* and *women* is due entirely to a difference between susceptible *men* and *women* in the likelihood of their experiencing a sufficient environmental exposure, despite the fact that, for every (*i*), the probability of an exposure to {*E*_*i*_} and {*E*_*iw*_} – i.e., *P*({*E*_*i*_}|*E*_*T*_) and *P*({*E*_*iw*_}|*E*_*T*_), respectively – are both *population-wide* and fixed during any specific *Time Period* (*E*_*T*_). Because such exposures are “*available*” to everyone, therefore, if the level of sufficient exposure differs between genders, any such difference will be due to a difference in *behavior* between susceptible *women* and *men* – i.e., to an increased exposure to, or avoidance of, susceptible environments by one or the other gender (perhaps consciously or unconsciously; or perhaps structurally, such as being due to differing gender-roles, differing occupations, differing recreational activities, etc.).

##### Exposure Considerations for i-Type Individuals

If both *men* and *women* are (or potentially could be) members of any specific *i-type* group, by definition, these *men* and *women* both have a non-zero probability of developing MS in response to every one of the (*v*_*i*_) sufficient sets of exposures within the {*E*_*i*_} family for this group. As discussed earlier, in these circumstances, these specific *i-types*, considered separately, will necessarily exhibit proportional hazards for the two genders (*see Methods #4A; above*). We have already defined the subset 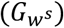– *see Methods #1A* – which includes all susceptible *women* 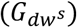 in (*Z*), where (*d* = 1,2, …, *mp*). If *<u>every</u> i-type* group includes (or, potentially, could include) both *men* and *women*, then, at every exposure level for a *man* {*H*(*a*) > *λ*}, we can define a proportionality constant (*R*_*i*_ > 0), such that the exposure level for any *i-type* susceptible *woman* {*K*_*i*_(*a*) > 0} can be expressed as:

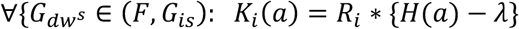

{*NB: Here, we don’t need to consider the i-type specific exposure for men, H*_*i*_(*a*), *because, by definition, it is always true that, if every i-type has the same threshold* (*λ* > 0) *then, for all* {*H*(*a*) > *λ*} *and for all* (*i*), *both* {*H*(*a*) − *λ* > 0]} *and* {*K*_*i*_(*a*) > 0}. *Therefore, in this circumstance, there will be some constant* (*R*_*i*_ > 0) *that permits this statement to be true for each* (*i*). *See below for a consideration of the impact of different i-types having different thresholds*.}

Also, because each 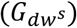 is a member of one or another of these posited *i-type* groups, we can define an exposure level 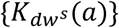 and a proportionality factor 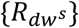 for each susceptible *woman* such that:

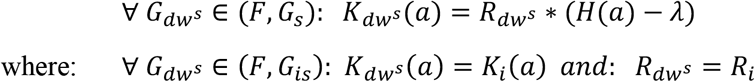

Thus, the expected exposure level for susceptible *women* can be expressed such that:

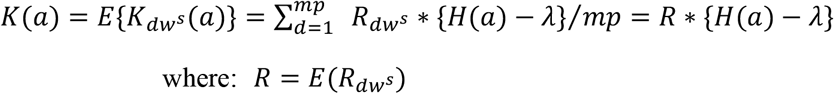

Consequently, if *women* and *men* can, potentially, be members of every *i-type* group, the hazards for *women* and *men* will always be proportional, although the hazard proportionality factor (*R*_*i*_) need not be the same for every *i-type* group. Alternatively, if *men* and *women* are of different *i-types*, but both are responding to the same environmental events, the hazards, again, will be proportional. Additionally, it is possible that the difference in threshold between *women* and *men* (*λ*_*i*_) may be different for individuals of different *i-types*. We consider, first, this possibility for those circumstance in which: (*λ* > 0). In this circumstance {*λ* = min(*λ*_*i*_) > 0} because, by definition, some *women* will begin to develop MS at this level of exposure. We can also define the threshold difference 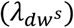 between each susceptible *woman* and that of susceptible *men*, in which case, because (*λ* > 0), by definition: (*λ*_*m*_ = 0) – *see Methods #4A*. Thus:

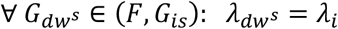

In this manner, the proportionality constants for each *i-type* (*R*_*i*_ > 0) and each *woman* 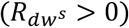 can be replaced by a *“adjusted”* proportionality constants 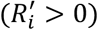 and 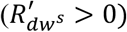 such that:

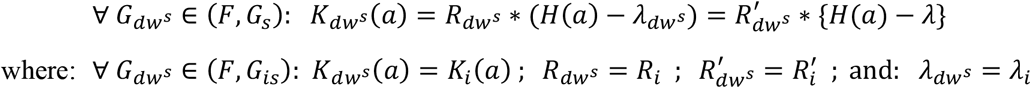

Thus, in this case, the expected exposure level for susceptible *woman* can be expressed such that:

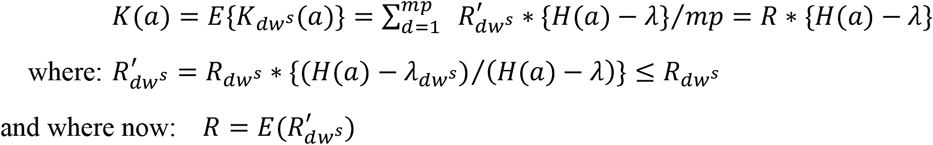

For the circumstance where: (*λ* < 0), the analysis is only changed in that: {*λ* = max(*λ*_*i*_)} and that, in this circumstance: (*λ*_*w*_ = 0). Thus, in either case, the hazards will still be proportional.

By contrast, if *men* and *women* are each responding to different environmental events, the hazards will not be proportional. In such a circumstance, *men* with MS would be envisioned as having a disease distinct from MS in *women*. Alternatively, perhaps, it could be that, for some autosomal genotypes – e.g., (*F, G*_*ia*_) and (*M, G*_*ia*_) – only *women* or only *men* could be in a the subset (*G*_*is*_) – i.e., if either: *P*(*F, G*_*ia*_, *G*_*is*_) = 0 or: *P*(*M, G*_*ia*_, *G*_*is*_) = 0 – whereas, for other autosomal genotypes, both *men* and *women* could be members of the same subset (*G*_*is*_). In such a circumstance, however, MS would then be envisioned as comprising three distinct diseases – one in *men* only, one in *women* only, and a third in both.

##### Contrasting the Possibilities that: (c = d) or (c < d)

When (*λ* > 0), to account for an increase in the (*F:M*) *sex ratio*, as shown in *Figure 3*, although it is possible for: (***c*** = ***d*** ≤ 1), this circumstance, nevertheless, seems unlikely. The ascending portion of the response curve is very steep, which indicates that any change in the (*F:M*) *sex ratio* is quite large in response to small changes in exposure. Thus, the window of possible changes in environmental exposure necessary to explain the Canadian data is quite narrow [6]. Moreover, following this narrow window, and for most of these response curves, the (*F:M*) *sex ratio* is declining – a circumstance, which is contrary to evidence [6,22-30]. Also, to achieve an *F:M sex ratio*, which reaches its current level (*i. e*., *p* ≈ 0.76), requires either that both the value of (*λ*) is small and the value of (*R*) is large (e.g. *Figures 3B–D*), or that the value of (*p*) is large (e.g. *Figures 3A*). Finally, the increase in failure rate (i.e., the increase in the penetrance of MS for the population), which was observed in Canada between the two *Time Periods*, was large (>32%) and especially prominent among women [3,6]. Each of these circumstances, although still compatible with the condition of: (***c*** = ***d*** = 1), still seems, nonetheless, to be somewhat at odds with the Canadian data, which demonstrates that the (*F:M*) *sex ratio* has been steadily, and gradually, increasing over many decades [6] and where, currently, the proportion of *women* among MS patients is quite high.

By contrast, the circumstances of *Figures 1D, 2D & 4A–D* (i.e., ***c*** < ***d*** ≤ 1), result in a continuously increasing (*F:M*) *sex ratio* with increasing exposure over most (or all) of the response curves, they easily account for the magnitudes of the observed (*F:M*) *sex ratios* and, in *Figures 1D & 4A–D*, they could also account for the observation that, at an earlier *Time Point* in the history of MS [40], in both Europe and the United States, the proportion of *men* among individuals with MS seemed to substantially exceed that of *women* such that:

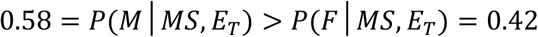

In conclusion, therefore, as indicated in *Methods #4C*, the condition of: (***c*** < ***d*** ≤ 1) is necessarily true for all circumstances, in which (*R* ≤ 1) and also, as discussed *above*, seems likely to be true for circumstances in which: (*R* > 1).

##### Summary Equations

For each of the two proportional hazard *Models*, we can use both the observed parameter values, the change in the (*F:M*) *sex-ratio*, and the change in *P*(*MS*) for Canada between any two *Time Periods* [6], and, thereby, construct each of these response curves in their entirety [3]. The values for: *Zw*_*2*_, *Zm*_*2*_, *Zw*_*1*_, *Zm*_*1*_, I, (***d***), *P*(*E*|*G,F*), *P*(*E*|*G,M*), and (*λ*) can then be determined [3] as:

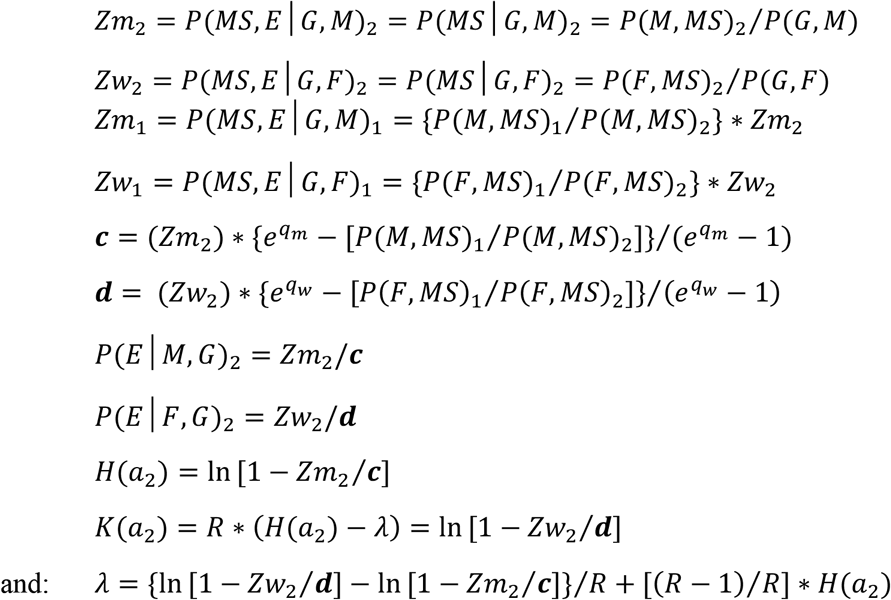

For the non-proportional *Model*, those parameters, which include (***c***) or (***d***), cannot be estimated from the observed changes in *P*(*MS*|*F*) and *P*(*MS*|*M*) over time. Notably, the values for *P*(*F*|*MS*)_1_, *P*(*F*|*MS*)_2_, and *P*(*MS*)_2_ have been directly or indirectly observed [3,6]. Also, the values of (*Zw*_1_), (*Zw*_2_), (*Zm*_1_), (*Zm*_2_), *P*(*E*|*G, F*)_2_, *P*(*E*|*G, M*)_2_, (***c***) and (***d***) are, not surprisingly, only related to the circumstances of either *men* and *women*, considered separately. Using a “substitution” analysis, we wrote a computer program, which incorporated the acceptable parameter ranges (*see Methods #2; above*) for the parameters {*P*(*G*) ; *P*(*MS*|*MZ*_*MS*_)_2_ ; *p* = *P*(*F*|*G*) ; *P*(*MS*)_2_ ; *P*(*MS*|*F, G*)_2_ ; *P*(*F*|*MS*)_1_ ; *r; s* ; *s*_*a*_ and *C*}, into the governing equations (*above*) and determined those combinations (i.e., solutions) that fit within the acceptable ranges for both the observed and non-observed parameters (*see Methods #2*). For this analysis, unlike for our *Cross-sectional Model*, we loosened the constraints on the values of (*r*) and (*s*) such that:

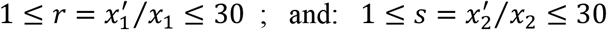

## Results

### 1. Cross-sectional Model

Assuming that the subset (*G*) conforms to the *<u>Upper Solution</u>* of the *Cross-sectional Model*, and using *Assertion C* (a*bove*) the range of values for the parameters *P*(*G*) and *P*(*MS*|*G*)_2_ were:

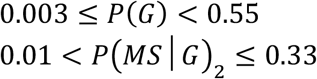

If we consider the more restricted range of (*s*_*a*_ < 1.9) for the impact of sharing the (*E*_*twn*_) environment with an *MZ-*twin, then:

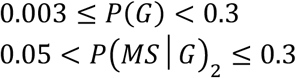

Assuming that the subset (*G*) does not conform to the *<u>Upper Solution</u>*, but that, considered separately, each of the subsets (*F,G*) and (*M,G*) do, and again using *Assertion C*, the range of values for the parameters *P*(*G*), *P*(*MS*|*G*)_2_, *P*(*MS*|*F, G*)_2_, *P*(*MS*|*M, G*)_2_, *P*(*G*|*F*), *P*(*G*|*M*); and the ratios (*x*_1_/*x*_2_) and 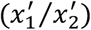 is:

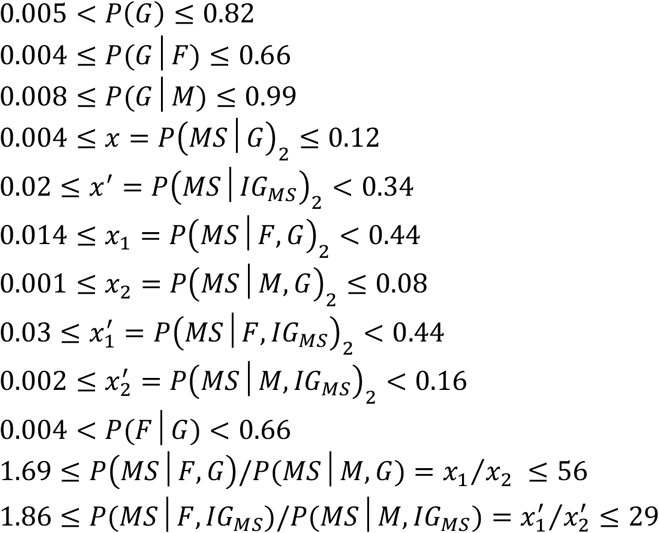

We previously concluded that it was possible (or even probable) that men might be disproportionately represented in the subset (*G*), although any marked disparity in this regard seemed implausible [3]. Therefore, if the restriction of: {*P*(*M*|*G*) ≤ 0.75} is included with the restrictions such that: (*s*_*a*_ < 1.9), (*r* ≤ 2) & (*s* ≤ 2), and also using the “current” *sex-ratio* data from Canada [6] such that: *P*(*F*|*MS*)_2_ = 0.74 − 0.78 ; (*see Methods #2*), then these estimates are unchanged except that:

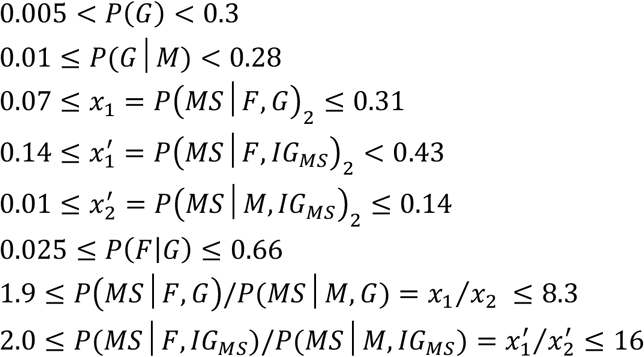

### 2. Longitudinal Model

Using the *Longitudinal Model*, assuming non-proportional hazards, the possible ranges for these various parameters were:

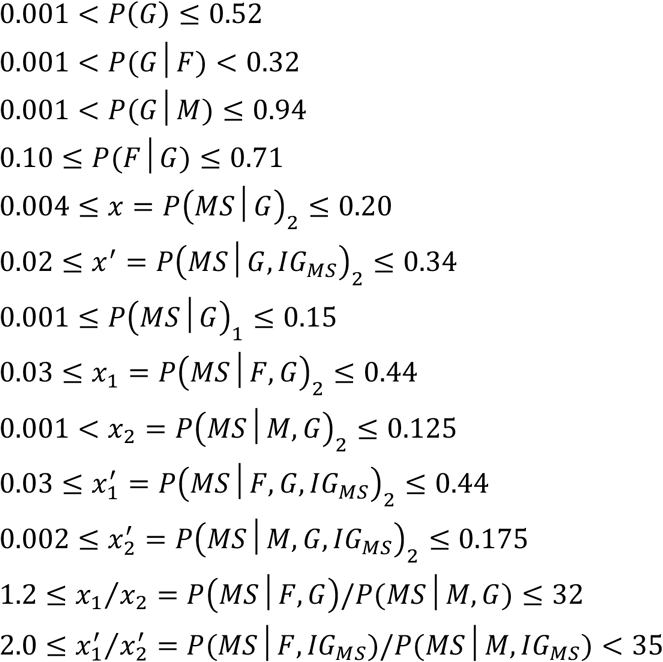

In addition, we found that the solution space for both (*r*) & (*s*) was restricted: (*r* < 20) and: (*s* < 30). Restricting the ranges such that: (*s*_*a*_ < 1.9), (*r* ≤ 2) & (*s* ≤ 2) changes the above estimations such that:

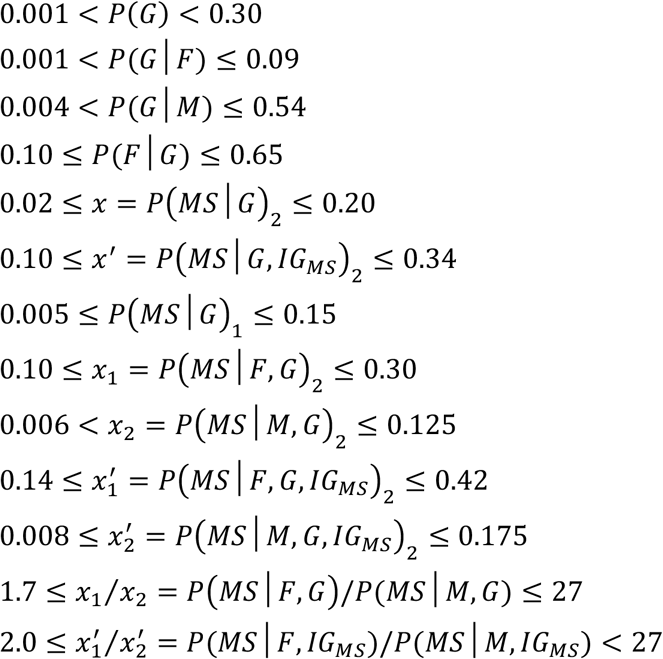

Using the *Longitudinal Model*, assuming (***c*** = ***d*** = 1) and, thus, with 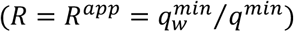, the possible ranges for these parameters are unchanged from the unrestricted non-proportional *Model* except for the additional estimations of:

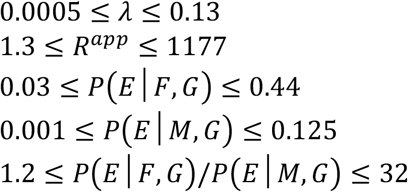

If the solution space were restricted: such that: (*s*_*a*_ < 1.9), (*r* ≤ 2) & (*s* ≤ 2), the above estimates are unchanged except that:

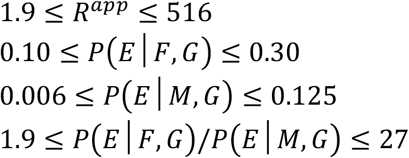

If (*R* = 1), these estimates are the unchanged from the non-restricted values above except:

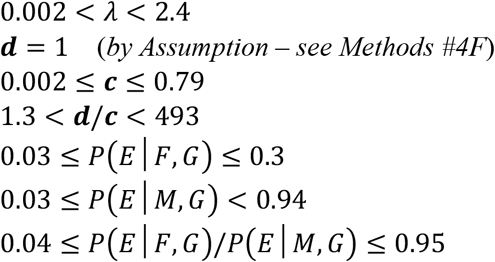

As in our analysis of the *Cross-sectional Model* (above), if the restriction of: {*P*(*M*|*G*) ≤ 0.75} is included with the above restrictions such that: (*s*_*a*_ < 1.9), (*r* ≤ 2) & (*s* ≤ 2) then, for (*R* = 1), these estimates become:

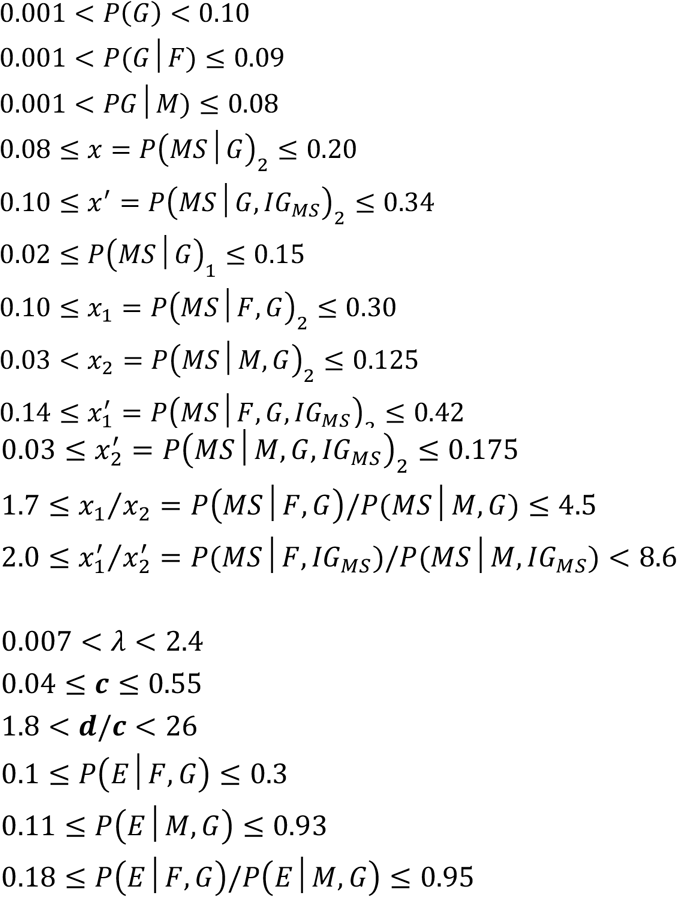

## Discussion

The present analysis provides considerable insight to the nature of susceptibility to MS. Thus, both of our *Models*, and the intersection of all our analyses, substantially support each other. For example, regardless of the whether the *Cross-sectional* or the *Longitudinal Model* was used, regardless of the whether the hazards are proportional and, if proportional, regardless of the proportional *Model* assumed, the consistently supported range for *P*(*G*) is:

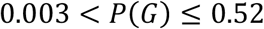

Thus, under any circumstance, a large percentage of the general population (≥ 48%), and likely the majority, must be impervious to getting MS, regardless of their environmental experiences. Consequently, if a person doesn’t have the appropriate genotype, they can’t get the disease. This conclusion is particularly evident for *women*, where:

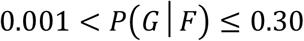

Thus, much of the population and most *women* lack this essential component of MS pathogenesis. In this sense, MS is fundamentally a genetic disorder.

Nevertheless, MS is also, fundamentally, an environmental disease. Thus, over the last several decades, the prevalence (and, thus, the penetrance) of MS has increased in many parts of the world [6,22-30]. Because genetic factors do not change this quickly, this fact implicates environmental factors as also critical to disease pathogenesis [3]. This conclusion is also supported by the observation that (*E*_*twn*_) environment seems to significantly impact the likelihood that an individual either has, or will subsequently develop, MS [7,31-37].

In considering the observations that have been made for *men* and *women* in Canada [6], one possible explanation is that the hazard functions for developing MS in the two genders are not proportional. In this view, each gender develops disease in response to distinct sets of environmental events (*see Methods #4A*) and, thus, MS in *women* represents a fundamentally different disease than MS in *men*. Moreover, in this non-proportional view, the environmental changes, which have taken place in Canada between the two *Time Periods* of 1941-1945 & 1975-1980 (whatever these are), would be interpreted as involving those events that impact MS development in susceptible *women* to a considerably greater extent than they do those events that impact MS development in susceptible *men*. However, even in this case, the limits derived for the parameters 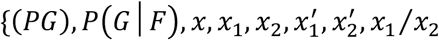 and 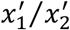 would still apply.

Nevertheless, the notion that MS in *men* and MS in *women* are fundamentally different diseases, involving distinct environmental events, seems implausible. Importantly, the view that hazards are proportional does not depend upon every individual (or *i-type*) having either the same proportionality constant or the same the threshold difference (*see Methods #4F*). Rather, it depends only upon the same environmental events having a non-zero probability of impacting the development of MS in both susceptible *women* and *men* (*see Methods #4A*). It is of note, therefore, that both genders seem to share very similar mechanisms of disease pathogenesis. Indeed, there have been several epidemiological observations that link MS, unequivocally, to environmental factors, to genetic factors, or to both and when these have been explored systematically, these factors seem to impact both *men* and *women* in a similar manner. For example, a *month-of-birth* effect has been reported in MS whereby, in the northern hemisphere, the risk of subsequently developing MS is greatest for babies born in May and least for babies born in November compared to other months during the year [41]. This *month-of-birth* effect was predicted to be inverted in the southern hemisphere [41] and, in fact, a subsequent *population-based* study from Australia found the peak risk to be for babies born in November-December and the nadir to be for babies born in May-June [42]. Although this *month-of-birth* effect is somewhat controversial [43], it has been widely (and reproducibly) reported by many authors and the effect is apparent in both *men* and *women* [41,42,44,45]. Thus, MS-risk seems to cycle throughout the year and this observation, if correct, clearly, implicates an environmental factor (or factors) – affecting both *men* and *women* alike – which is (are) linked to the solar cycle and occur(s) during the intrauterine or early post-natal period [8]. Second, the recurrence risk of MS is generally found to be greater in a co-twin of a *DZ-*twin proband with MS compared to a non-twin co-sibling of a sibling proband with MS [7,31-37]. This effect also implicates an environmental factor that occurs in proximity to the birth and is apparent in both *men* and *women* [3,8]. Third, it is widely reported that MS becomes increasingly prevalent in those geographic regions, which lie farther (either north or south) from the equator [8,46,47]. This observation could implicate either environmental or genetic factors but, regardless, this gradient is apparent in both *women* and *men* [46,47]. Fourth, evidence of a prior *EBV* infection is found in essentially all MS patients [8,48]. If this infection is actually present in 100% of MS patients, then this indicates that an *EBV* infection is a necessary factor in the causal pathway leading to MS for all susceptible *women* and *men* [8]. Fifth, vitamin D deficiency has been implicated as being an environmental factor in MS pathogenesis [8,49-53] and this factor is related to MS in both *men* and *women* [49-52]. And lastly, smoking tobacco has been implicated as being environmental factor associated with MS pathogenesis [8,54] and, again, this factor is associated with MS in both *women* and *men* [54].

Also, the genetic basis of MS seems to be very similar in both *women* and *men*. Thus, the strongest genetic associations with MS are for certain haplotypes within the *HLA*-region on the short arm of Chromosome 6 [55-59] and, in the predominantly Caucasian population of the Wellcome Trust Case Control Consortium (*WTCCC*) dataset [60,61], the most strongly MS-associated haplotypes in this region are similarly associated with MS in both *women* and *men* (*Tables 3 & 4*).

**Table 3.** MS Associations for Class I and Class II *HLA-*Haplotypes in *Men* and *Women*^*\**^

| *Haplotype* | *OR*<sup>†</sup><br>(*Women*) | *p* | *OR*<sup>†</sup><br>(*Men*) | *p* |
| --- | --- | --- | --- | --- |
| *DRB1* *15:01~ *DQB1* *06:02 – 1 copy<sup>‡</sup> | 3.1 | < E-182 | 2.7 | < E-82 |
| *DRB1* *15:01~ *DQB1* *06:02 – 2 copies<sup>‡</sup> | 6.5 | < E-110 | 6.2 | < E-70 |
| *DRB1* *03:01~ *DQB1* *02:01 – 1 copy<sup>‡</sup> | 1.1 | < 0.05 | 1.2 | < 0.01 |
| *DRB1* *03:01~ *DQB1* *02:01 – 2 copies<sup>‡</sup> | 2.9 | < E-21 | 2.2 | < E-7 |
| *A* *02:01~ *C* *05:01~ *B* *44:02 – 1 copy | 0.6 | < E-7 | 0.6 | < E-3 |

<sup>\*</sup> Class I or Class II haplotypes in the WTCCC within the human leukocyte antigen (*HLA*) region on the short arm of Chromosome 6 [60,61]. These haplotypes include either *HLA* Class I (*A*, *C* & *B*) or *HLA* Class II (*DRB1* & *DQB1*) alleles.
<sup>†</sup> Odds ratio (*OR*) for MS in *men* and *women* having either 1 or 2 copies of each haplotype except for the *A*\*02:01~*C*\*05:01~*B*\*44:02 haplotype, which had too few observations of the 2-copy data. Each was compared to individuals having no copies of the other 2 *haplotypes* (95% *CI* range in parenthesis). The *p-values* are expressed in scientific notation as powers of 10 (E).
<sup>‡</sup> The difference in *OR* between possessing 1 or 2 copies of the *DRB1*\*15:01~*DQB1*\*06:02 01 haplotype was significant for both *women* ( $p < 10^{-13}$ ) and *men* ( $p < 10^{-10}$ ). Similarly, the difference in *OR* between possessing 1 or 2 copies of the *DRB1*\*03:01~*DQB1*\*02:01 haplotype was significant for both *women* ( $p < 10^{-14}$ ) and *men* ( $p < 0.001$ ).

**Table 4.**
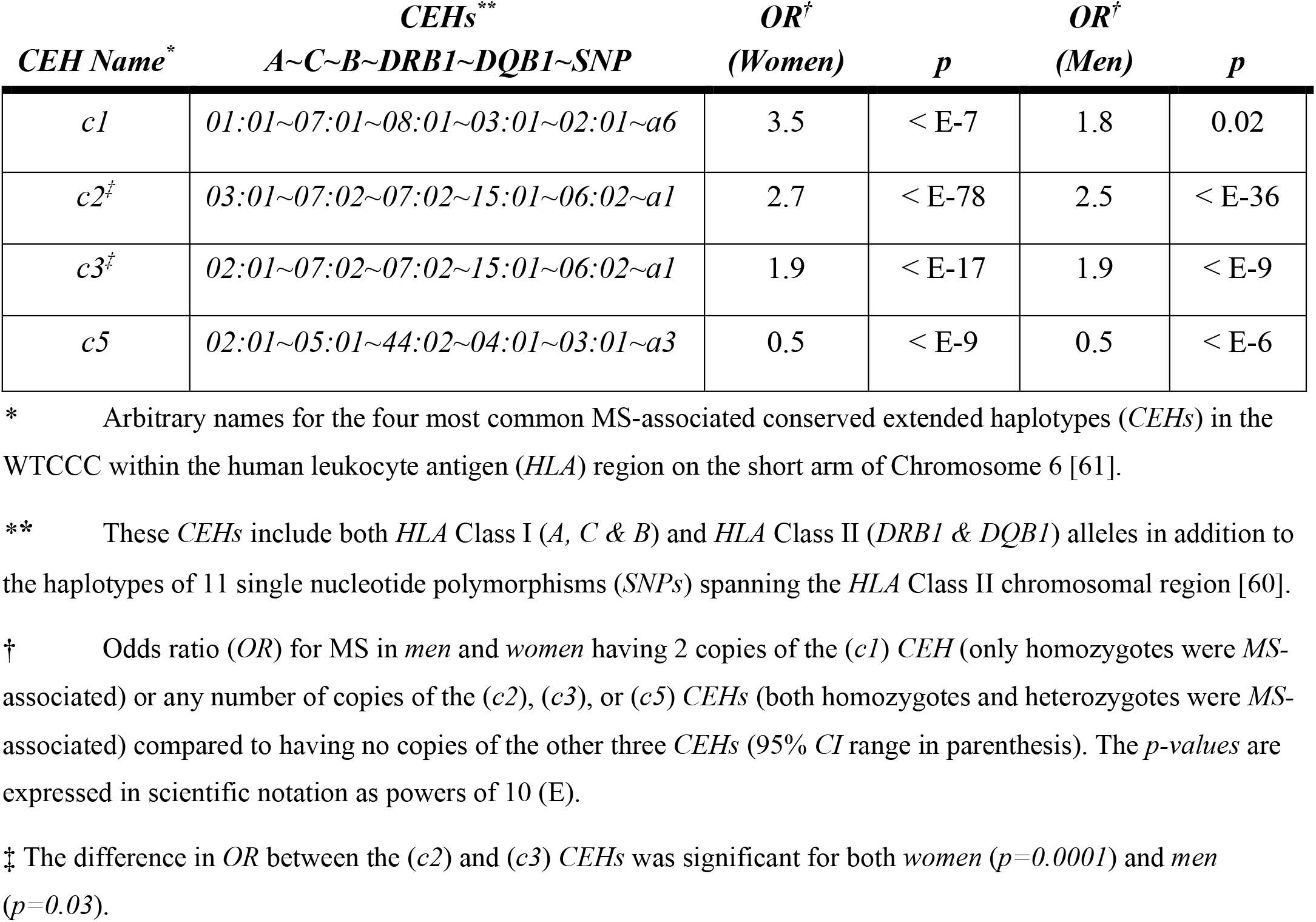
MS Associations for Extended *HLA-*Haplotypes Region in *Men* and *Women*

In addition, it is useful to further consider the notion that, to explain the threshold difference, susceptible *women* might require a more *“intense”* environmental exposure than susceptible *men*. For example, in *Methods #4F*, we entertained the notion that different *i-types* might potentially require different families of sufficient environmental exposure {*E*_*i*_}, each with a potentially different threshold difference (*λ*_*i*_), and each with a potentially different proportionality constant (*R*_*i*_) and, potentially, with many different sufficient sets within each family. It seems rather surprising, if such marked variability truly existed, that this could possibly lead to a circumstance in which *<u>all</u>* susceptible *women* required a more “*intense*” exposure compared to *<u>some</u>* susceptible *men*. By contrast, if all *i-type* individuals required the same set (or sets) of environmental exposure, a consistent difference in the *“intensity”* of the required exposures might be easier to rationalize (see our earlier discussion regarding the possible role that *EBV* and Vitamin D deficiency might have in MS pathogenesis – *Methods #4F*). Also, if the different *i-types* require the same set (or sets) of environmental exposure, this also makes it easier to rationalize the fact that those environmental factors, which have been consistently identified as MS-associated, have been linked to MS, generally, but not to any particular subgroup [8,41,42,44-54].

As noted earlier (*see Methods #4A & 4F; Equations 7 & 12b*), by assuming that: (***c*** = ***d*** ≤ 1), we are also assuming that the difference in disease expression between *men* and *women* is due entirely to a difference in the likelihood of their experiencing a sufficient environmental exposure. Therefore, because currently (*Zw*_2_ > *Zm*_2_) – *see Methods #2C* – those conditions in which (***c*** = ***d*** ≤ 1) would necessarily lead to the conclusion that, currently, susceptible *women* are more likely to experience a sufficient environment compared to susceptible *men* despite the fact that the probabilities of exposure to each family, *P*({*E*_*i*_}|*E*_*T*_) and *P*({*E*_*iw*_}|*E*_*T*_), are fixed constants during any (*E*_*T*_).

Moreover, there are also several additional lines of evidence, which, taken together, strongly suggest that the circumstance of (***c*** = ***d*** ≤ 1) is unlikely. First, for all (*R* ≤ 1), it must be the case that: (***c*** < ***d*** ≤ 1) – *see Methods #4D & 4E*. Second, as discussed earlier (*see Methods #4F*), the response curves required for conditions where: (***c*** = ***d*** ≤ 1) & (*R* > 1) have very steep ascending portions and, thus, present only a narrow window of opportunity to explain the Canadian data [6] regarding the changes in the (*F:M*) *sex ratio* and its magnitude over time (*see Methods #4F; see also Figure 3*). Also, for these response curves, following this narrow window, and contrary to evidence [6], the (*F:M*) *sex ratio* decreases with increasing exposure (*Figure 3*). By contrast, these Canadian data suggest that there has been a gradual and sustained increase in the (*F:M*) *sex ratio* over the past several decades [6]. Third, as noted in *Methods #1D*, there seems to be little impact of the (*E*_*sib*_) environment on the development of MS. However, when (***c*** = ***d***) the only explanation for (*R* > 1) is a disproportionate exposure to sufficient environments experienced by *women* (*see above*). Thus, proband siblings and their non-twin co-siblings (both *men* and *women*), despite sharing common genes and a common childhood environment, still depend upon (and differ in) only their (*E*_*pop*_) exposures to develop their MS. Fourth, if (*λ* > 0), as it must be for all for (*R* ≥ 1), there is an inherent tension between the fact that *men* are responding to a broader range of environmental exposures (i.e., to both the less and the more *intense* exposures) compared to *women* (who respond to only the more *intense* exposures) and, yet, that *women* are more likely to experience a sufficient environmental exposure compared to *men*.

Fifth, and most importantly, for each of the known (or suspected) environmental factors related to MS pathogenesis, there is no evidence to suggest that *women* are disproportionately experiencing them. Thus, the *month-of-birth* effect is equally evident for *men* and *women* [41,42,44,45]; the latitude gradient is the same for both genders [8,46,47]; the impact of the (*E*_*twn*_) environment is the same for *men* and *women* [3]; By young adulthood (i.e., 20-25 years), the likelihood of an *EBV* infection (a factor probably in the causal chain leading to MS), is about equal (∼95%) for both genders, although infection likely occurs earlier among *women* [8,62,63]; vitamin D levels are the same in both genders [49-53]; and smoking tobacco is actually more common among *men* [8,54]. Taken together, these epidemiological observations suggest that *women* and *men* are currently experiencing the same relevant environmental events in an equivalent manner. Therefore, each of these observations suggests that:

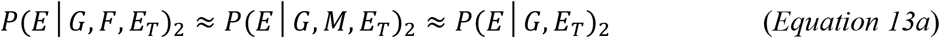

In this context, the possibility that (***c*** = ***d*** ≤ 1) & (*R* > 1) – which are noted in the *Results* and are depicted in *Figure 3* – seems remote, especially given the facts that the relevant exposures are *population-wide* and that the difference between (*Zw*) and (*Zm*) can only be explained by a disproportionate exposure to sufficient environments by susceptible *women* (*see Equation 10a*) – a circumstance for which there is decidedly no evidence (*see above; see also, Equation 12b*). Therefore, if *Equation 13a* is correct, then also:

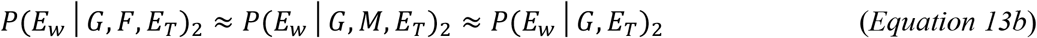

Moreover, because, the population experiences the same level of exposure (*u* = *a*) during any (*E*_*T*_), therefore, from *Equations 6* and *9*, these approximate equivalences (if correct) would indicate both that: (***c*** < ***d*** ≤ 1) and that: (*R* ≈ 1) – *see Methods #4F*. Such a configuration easily explains an increasing (*F:M*) *sex ratio* and its magnitude throughout most (or all) of the response curves (*see Figures 4A–D*), it accounts for a time in MS history where the disease may have been more prevalent in *men* [40] − e.g., *Figures 4C & 4D* – and it does not assume that, even though susceptible *men* and *women* have the same *population-wide* exposures, {*P*({*E*_*i*_}|*E*_*T*_)} and {*P*({*E*_*iw*_}|*E*_*T*_)}, there is, nonetheless, a marked and systematic increase in the exposure to (*E*_*w*_) for susceptible *women* compared to *men*. However, any condition, for which (***c*** < ***d***), does require that some susceptible *men* will never develop MS, even when the correct genetic background occurs together with an environmental exposure sufficient to cause MS in those individuals. Indeed, if, as suggested: (*R* = 1), then, necessarily, (***c*** < ***d***) – *see Methods #4F, above* − and, indeed, such *men* will comprise 22-98% of the susceptible *male* subset (*M, G*). Naturally, in such a circumstance, it seems likely that the proportion of *women* who ultimately develop MS, given the same conditions, will also be less than unity (e.g., *Figures 4B & 4D*). However, because we needed to assume that: (***d*** = 1) for the purposes of our analysis, this possibility cannot be addressed using the Canadian data.

**Figure 4.**
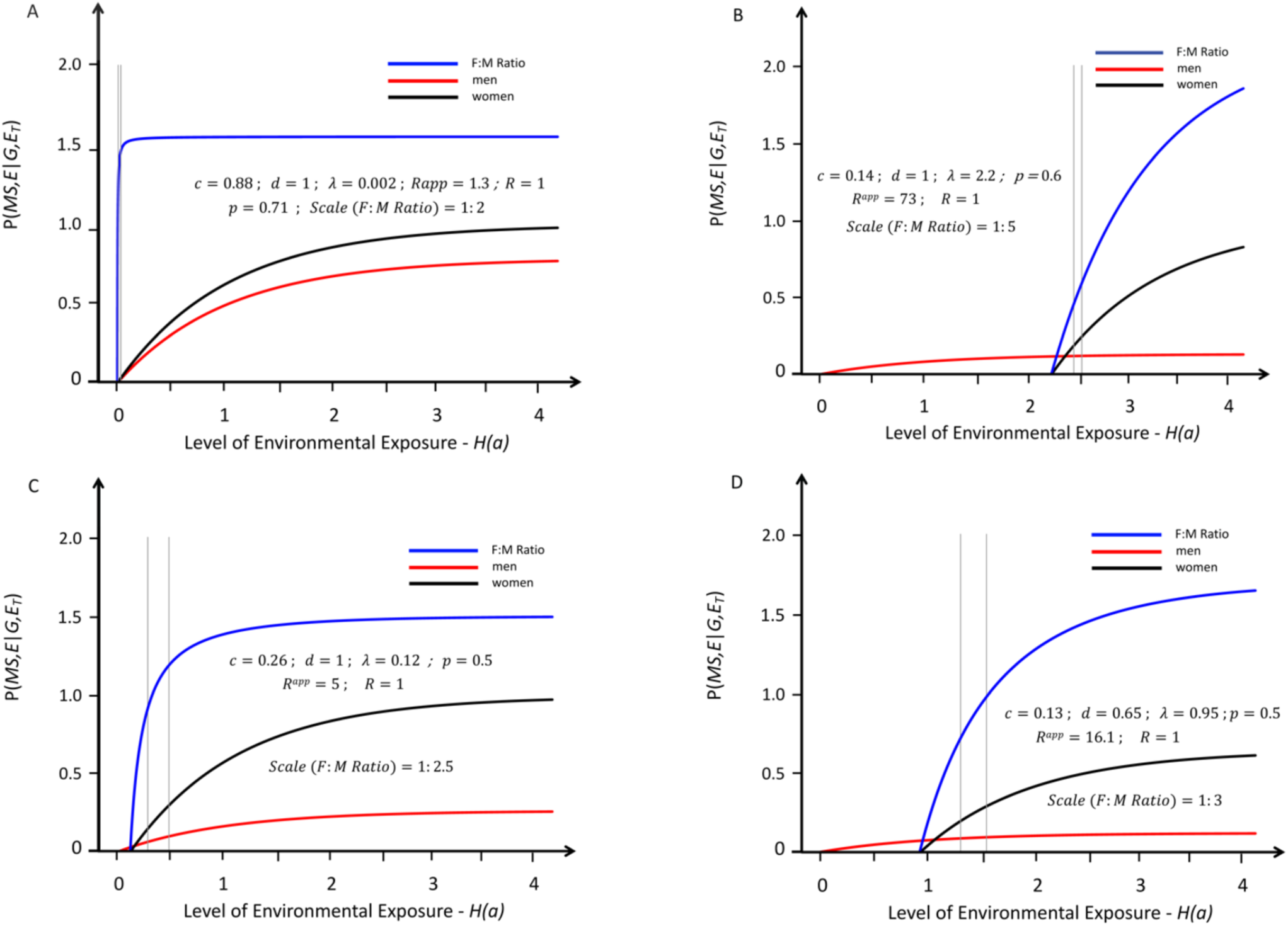
Response curves for the likelihood of developing MS in genetically susceptible *women* (black lines) and *men* (red lines) with an increasing probability of a “sufficient” environmental exposure – *see Methods #1B*. Like *Figure 1*, the curves depicted are also proportional (*R* = 1), but, for these, the environmental threshold in *women* is greater than that it is in *men* – i.e., these are conditions in which: (*λ* > 0). Also, these curves represent the same solutions as those depicted in *Figure 3* except that these are for conditions in which (***c*** < ***d*** ≤ 1). The blue lines represent the change in the (*F:M*) *sex ratio* (plotted at various scales) with increasing exposure. The thin grey vertical lines represent the portion of the response curve (for the depicted solution), which represents the actual change in the (*F:M*) *sex ratio* that occurred between *Time Periods #1 & #2*). Unlike the curves presented in *Figure 3*, however, an increase in the (*F:M*) *sex ratio* with increasing exposure is observed for any two-point interval along the entire response curves and, except for *Figure A*, the grey lines are clearly separated.

Some of the individuals who don’t seem to develop MS despite having an environmental exposure sufficient to cause MS, no doubt, will have subclinical disease. Indeed, as suggested by several autopsy studies, the prevalence of “asymptomatic” MS in the population (*Z*) may be as high as ∼0.1% [64-67]. Moreover, such a figure is generally supported by several magnetic resonance imaging (*MRI*) studies of asymptomatic individuals [68,69]. Nevertheless, although these considerations suggest that some proportion of MS can be asymptomatic, this fact seems unlikely to account for any difference either (***c***) from (***d***), or of (***c***) from the expected 100% occurrence of MS in *men* who are both genetically susceptible and, in addition, experience an environment sufficient to cause MS given their specific genotype. Thus, if asymptomatic disease did account for (***c***) being less than (***d***), then *men* should account for a disproportionately large percentage of these asymptomatic individuals. However, this is decidedly not the case. Rather, *men* account for only 16% of the asymptomatic individuals detected by *MRI* [68,69] −a percentage well below their proportion of symptomatic cases [3]. Consequently, if (***c*** < ***d*** ≤ 1), as the Canadian data [6] seems to indicate, then chance must play a role in disease pathogenesis.

Alternatively, however, perhaps our previous definition of exposure “*intensity*” does not account properly for certain other potential aspects of exposure “*intensity*”, which might play an important role in disease pathogenesis. As a concrete example of this notion, suppose, as before, that one (or more) of the sufficient sets of exposures for the *i*^*th*^ susceptible individual includes both a deficiency of vitamin D and a prior *EBV* infection [3], each occurring during some critical period of the person’s life (not necessarily during the same period). Furthermore, suppose that, with all other necessary factors being equal in the *i*^*th*^ susceptible individual, a mild vitamin D deficiency for a short period during the critical time, together with an asymptomatic *EBV* infection at age 10, causes MS to develop 10% of the time, whereas a more prolonged, and more marked, vitamin D deficiency during the critical period, together with a symptomatic *EBV* infection (mononucleosis) at age 15, causes MS to develop 75% of the time. Notably, each of these posited conditions is sufficient, by itself, to cause MS; the only difference is in the likelihood of this outcome, given the different levels (i.e., “*intensity*”) of exposure. Although this notion of “*intensity*” differs from our previous definition and can’t be easily quantified, presumably, there will be a positive correlation between an increasing “*intensity*” of this exposure (whatever this means operationally) and an increasing risk of MS for each susceptible individual. Moreover, each susceptible individual must reach a *maximum* likelihood of developing MS as the “*intensity*” of their exposure increases. This maximum may be at 100% or it may be at something less than this but, whatever it is, there must be a maximum for each person. In addition, unlike our previous definition of *P*{*E*_*i*_}, where only one sufficient set of exposures was necessary, here, an individual for whom two or more of their sufficient sets of exposure occur, may experience a greater “*intensity*” of exposure than if only one set occurs. Nevertheless, none of these circumstances alters the fact that each person will still have their “*maximum*” likelihood of developing MS under optimal environmental conditions. We can then define the “*intensity*” of exposure – *P*(*E*|*G, E*_*T*_) – as the average (or expected) “*intensity*” of exposure (however this is measured) experienced by members of the (*G*) subset, given the environmental conditions of the time (*E*_*T*_). When no sufficient exposure occurs for any member of (*G*): *P*(*E*|*G, E*_*T*_) = 0. When the “*intensity*” has increased to the point where every member of (*G*) has reached their maximum likelihood of developing MS then: *P*(*E*|*G, E*_*T*_) = 1. And, again, we can define (*u*), as the odds of exposure:

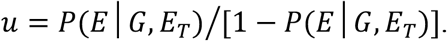

Although, clearly, this conceptualization of exposure *intensity* is different (and perhaps more realistic) than the “*sufficient*” exposures considered earlier, two of its features are particularly noteworthy. First, randomness is integral to this notion of exposure “*intensity*”. Thus, disease expression at low “*intensity*” exposures, by definition, incorporates an element of chance because the likelihood of developing MS under these conditions must be less than the *maximum*, for at least some susceptible individuals. If not, then this “*intensity*” of exposure would have no impact on anyone, and this *Model* becomes equivalent to the “*sufficient*” exposures *Model* considered earlier. Second, despite exposure being measured differently, and despite the hazard functions likely being different, all the equations and transformations presented in *Methods #4A–F* (*above*) as well as the calculated response curves are unchanged by measuring exposure as *“intensity”* in this manner rather than as *“sufficiency”*. Indeed, this conclusion applies to any measure of exposure, which incorporates the notion of “sufficient sets” of environmental exposure defined earlier (*see Methods #1B*).

Thus, by any measure, the Canadian data [6] seem to indicate that there is a stochastic factor (i.e., an element of chance) in MS pathogenesis, at least for *men*, which determines, in part, who gets the disease and who does not. Such a conclusion might be viewed as surprising because, in the universe envisioned by many physicists, events are (or seem to be) deterministic. For example, imagine a rock thrown at a window. If the rock has a mass, a velocity, and an angle of impact sufficient to break the window, given the physical state of the window at the moment of impact, then we expect the window to break 100% of the time. If the window only breaks some of the time, likely, we would conclude that we hadn’t adequately specified the sufficient (i.e., initial) conditions. If the *population-based* observations in over 29,000 Canadian MS patients are to be believed, however, this is not so for the development of MS. Even for an individual with a susceptible genotype and an environmental experience sufficient to cause disease given their specific genotype, they still may or may not develop the illness. This result cannot be ascribed to contributions from other, unidentified, environmental factors because each set of environmental circumstances considered here is defined to be sufficient, by itself, to cause MS in that specific susceptible individual. If other environmental conditions were needed to cause MS reliably in that individual, these conditions would already be necessary components of these sufficient environments. Even altering the definition of exposure to include the importance of different *intensities* of exposure doesn’t alter this conclusion. Certainly, the invocation of stochastic processes in disease pathogenesis requires replication, both in MS and in other disease states, before being accepted as fact. Nevertheless, if replicated, such a result would imply that there is a fundamental randomness to the behavior of some complex physical systems (e.g., organisms), in contrast to the apparent determinism of physical laws [70].

It is of note that some authorities have argued (albeit controversially) from fundamental physical principles that true randomness (i.e., thermodynamic equilibrium or maximum entropy) was a primordial property of our universe in the earliest tiny fraction of a second of the big bang and that this inherent randomness is reflected by a currently observable randomness for both microscopic (i.e., quantum uncertainty) and macroscopic descriptions of the universe [71]. By contrast, the deterministic hypothesis envisions that earliest state of the universe was one of minimum entropy and asserts that, when we perceive certain macroscopic events as being due to chance, this perception is illusory and merely a reflection of our ignorance regarding the relevant initial conditions [70,71]. One author has stated this deterministic point of view succinctly by noting that, while “the quantum equations lay out many possible futures, … they deterministically chisel the likelihood of each in mathematical stone” [70]. Obviously, the question of which, if either, of these alternate views of the universe represents reality has far-reaching implications [70,71]. Perhaps the best contemporary evidence for macroscopic randomness, cited by proponents of the non-deterministic worldview, is the case of biological evolution by means of natural selection [71]. Thus, natural selection is envisioned to be a non-sentient process, which depends upon the occurrence of apparently random events and, using these events, permits living species to respond both continuously and adaptively to the varying environmental conditions of different times, or different places, or both. Moreover, the direction, in which any new species evolves, is seemingly not predictable but, rather, depends upon the nature of the specific random events, which take place.

Placed into a broader context, this biological evolution, which has been so clearly documented on Earth, is probably best viewed as a part of (or as a continuation of) the process of chemical evolution – a process that began only a few minutes after the onset of the big bang, and at a time when the universe was composed of 75% hydrogen, 25% helium, and a small admixture of lithium [70,72]. The chemistry of this early universe was extremely rudimentary. Helium is a nobel gas and reacts with essentially nothing. Hydrogen and lithium combine to form only a few simple chemical compounds such as lithium hydride (*LiH*) and molecular hydrogen (*H*_2_). A more complex chemistry (and, in particular, the chemistry necessary to create and sustain life and, also, to permit biological evolution) only evolved later with synthesis of the heavier atomic elements – a synthesis that, following these first few minutes of the big bang, only occurred with the collapse and/or explosion of massive stars at the end of their life cycle [70,72]. This synthesis, and the subsequent build-up of heavier elements in the universe, was gradual and took time. Also, this process of chemical evolution continues to this day, not only with the ongoing synthesis of heavier elements inside contemporary stars and the interactions of these elements with each other throughout the universe, but also with the synthesis of a multitude of novel chemical compounds, created by living organisms (including humans). Moreover, each step of this evolutionary sequence seems to require the occurrence of random events – i.e., which nuclei happen to collide, whether they fuse, whether (and when) they decay, which star becomes a supernova, where and when these supernovas occur, which life-forms evolve, and under what circumstances, with what chemistries, and in what places, etc.

In this broader context, then, it is hard to imagine that the past and future course of this evolutionary sequence was, and is, a pre-determined outcome and yet, in the case of biological evolution on Earth, for the macroscopic process that produces it, both to be so exquisitely adaptive to contemporary external events and yet, to be so completely dependent upon apparently random occurrences. Nevertheless, it is extremely difficult to prove that any macroscopic process (including this evolutionary sequence) is truly random. Despite this difficulty, however, the hypothesis of determinism is quite fragile in the sense that, if the true randomness of even one macroscopic process or event could be established, the hypotheses of determinism would be undermined. Consequently, if replicated, any demonstrated randomness in MS disease expression or in the expression of any other disease {i.e., any circumstance, in which either: (***c*** < ***d*** = 1) or: (***c*** ≤ ***d*** < 1)}, would provide strong empiric evidence in support for the non-deterministic worldview. By contrast, the deterministic worldview requires the condition that: (***c*** = ***d*** = 1).

There are two features of the response curves in *men* and *women* that merit further comment. First, the plateaus for these curves (if, in fact, ***c*** < ***d*** ≤ 1) – e.g., *Figures 1D, 2D, 4A–D* – reflect this inherent randomness in the process of disease development. Indeed, in this circumstance, it would be this randomness, rather than the genetic and environmental determinants, which lies at the heart of the difference in disease expression between *men* and *women*. Thus, genetically susceptible *women*, who experience an environment sufficient to cause MS given their genotype, are more likely to develop disease compared to susceptible *men* in similar circumstances. Consequently, if (***c*** < ***d***), there must be something about “*female-ness*” that favors disease development in *women* over *men* although, whatever this is, it is not part of any causal chain of events leading to disease (i.e., in the sense that, if a coin-flip determines, in part, an outcome, then this random event is not part of any *<u>causal</u>* chain). As noted above, if either (***c*** < ***d*** = 1) or: (***c*** ≤ ***d*** < 1), then disease development in the setting of a susceptible individual experiencing a sufficient exposure must include a truly random event (at least for *men*). Moreover, if: (***c*** < ***d***), the fact that this random process favors disease development in *women* does not make it any less random. For example, the flip of a biased coin is no less random than the flip of a fair coin. The only difference is that, in the former circumstance, the two possible outcomes are not equally likely. In the context of MS, “*female-ness*” would then be envisioned to bias the coin differently than does “*male-ness*” (whatever these terms mean).

Second, the thresholds reflect the minimum exposure at which disease expression begins and the response curves, with increasing exposure, that follow this onset [3], need to account for both the increasing prevalence of MS and the steadily increasing proportion of *wome*n among MS patients [6,22-30,40]. If the hazards in *men* and *women* are not proportional, as discussed *above*, little accounting is necessary. By contrast, if the hazards are proportional and if both: (***c*** < ***d*** ≤ 1) & (*λ* > 0), then this could account for all of the epidemiological observations – i.e., the increasing prevalence of MS [22-30], the continuously increasing proportion of *women* among MS patients [6,22-30,40], the magnitudes of the observed (*F:M*) *sex ratios* [3,6], and a 1922 study [3,40], in which MS prevalence in both the United States and Europe was reported to be substantially higher in *men* than in *women* (e.g., *Figures 1D, 4A–D*).

During the development of our *Longitudinal Model*, we observed that when the prevailing environmental conditions of a time (*E*_*T*_) were such that: {*P*(*E*|*E*_*T*_) = 0}, no member of (*G*) could develop MS. Previously, we considered the possibility that such an environment might not be possible to achieve because some susceptible individuals might be able to develop MS under *<u>any</u>* environmental conditions – i.e., if these cases were “purely genetic” [3]. Upon further reflection, however, such a possibility seems remote. For example, the first clinical description of MS was published in 1868 by Charcot although earlier pathological descriptions predated this clinical description by ∼30 years [73,74]. Perhaps, the earliest described case of MS was that of Saint Lidwina of Schiedam (c. 1396) although the argument that Augustus d’Este (c. 1822) suffered from MS seems more unequivocal [73]. Even though many human afflictions were initially described during the advent of modern medicine in the 19^th^ century, MS is a rather distinctive disorder, and it seems likely that, if MS existed, case descriptions (familiar to us) would have appeared in earlier eras. Moreover, with the onset of the industrial revolution in the late 18^th^ or early 19^th^ century, the environmental conditions began to change substantially (especially for humans). Therefore, both MS as a disease and permissive environmental conditions seem likely to be of relatively recent onset. More importantly, ever since its original description, MS seems to be changing in character – a fact that underscores the critical importance of environmental factors in MS pathogenesis. For example, although considered uncommon initially, ever since Charcot’s initial characterization, MS has become increasingly recognized as a common neurological condition [74-76]. Also, in the 19^th^ century Charcot’s triad of limb ataxia, nystagmus (internuclear ophthalmoplegia), and scanning (cerebellar) speech was considered typical whereas, today, while this triad still occurs, such a syndrome is unusual [74-76]. Moreover, in the late 19^th^ and early 20^th^ centuries, the disease was thought to be more (or equally) prevalent in *men* compared to *women* [40,74,75], whereas, today, *women* account for 66–76% of the cases [3,76]. Also, in many parts of the world, MS is increasing in frequency, particularly among *women* [6,22-30]. Indeed, in Canada, *P*(*MS*) has increased by an estimated minimum of 32% over a span of 35–40 years [3] – a circumstance which has led to a 10% increase in the proportion of *women* among MS patients (*p* < 10^−6^) over the same time-interval [6].

By contrast, those genetic markers, which are associated with MS, seem to have been present for far greater periods of time. For example, the best established (and strongest) genetic associations with MS are for certain haplotypes within the *HLA* region on the short arm of chromosome 6 (e.g., *Table 3*), including haplotypes such as *DRB1*15:01*∼*DQB1*06:02; DRB1*03:01*∼ *DQB1*02:01*; and *A*02:01*∼*C*05:01*∼*B*44:02* [55-59]. Each of these haplotypes, as well as each of the conserved extended haplotypes (*CEHs*) – *see Table 4* – is well represented in diverse human populations around the globe [77,78] and, thus, these haplotypes must be of ancient origin. Presumably, therefore, the absence of MS prior to the late 14^th^ (and possibly the early 19^th^) century, together with the markedly changing nature of MS over the past 200 years, points to a change in environmental conditions as the basis for the recent occurrence of MS as a clinical entity and for the changes in MS epidemiology, which have taken place over the past two centuries. Consequently, it seems that {*P*(*E*|*G, E*_*T*_) = 0} is possible under those environmental conditions that existed prior to the late 14^th^ century and, thus, that “purely genetic” MS does not exist.

In summary, the development of MS (in an individual) requires both that they have an appropriate genotype (which is uncommon in the population) and that they have an environmental exposure sufficient to cause MS given their individual genotype. Nevertheless, even when the necessary genetic and environmental factors, required for MS pathogenesis, co-occur for an individual, this still seems to be insufficient for that person to develop MS. Thus, disease pathogenesis, even in this circumstance, seems not to be deterministic but, rather, to involve an important element of chance.

## Data Availability

This manuscript uses data that is already published and is publicly available

## Acknowledgement

We are especially indebted to John Petkau, PhD, Professor Emeritus, Department of Statistics, University of British Columbia, Canada. Dr. Petkau helped immeasurably with this project; devoting numerous hours of his time to critically reviewing early drafts of this manuscript and providing an invaluable contribution both to the clarity and to the logical development of the mathematical and statistical arguments presented herein.

